# A multiplexed Cas13-based assay with point-of-care attributes for simultaneous COVID-19 diagnosis and variant surveillance

**DOI:** 10.1101/2022.03.17.22272589

**Authors:** Maturada Patchsung, Aimorn Homchan, Kanokpol Aphicho, Surased Suraritdechachai, Thanyapat Wanitchanon, Archiraya Pattama, Khomkrit Sappakhaw, Piyachat Meesawat, Thanakrit Wongsatit, Artittaya Athipanyasilp, Krittapas Jantarug, Niracha Athipanyasilp, Juthamas Buahom, Supapat Visanpattanasin, Nootaree Niljianskul, Pimchai Chaiyen, Ruchanok Tinikul, Nuanjun Wichukchinda, Surakameth Mahasirimongkol, Rujipas Sirijatuphat, Nasikarn Angkasekwinai, Michael A. Crone, Paul S. Freemont, Julia Joung, Alim Ladha, Omar Abudayyeh, Jonathan Gootenberg, Feng Zhang, Claire Chewapreecha, Sittinan Chanarat, Navin Horthongkham, Danaya Pakotiprapha, Chayasith Uttamapinant

## Abstract

Point-of-care (POC) nucleic acid detection technologies are poised to aid gold-standard technologies in controlling the COVID-19 pandemic, yet shortcomings in the capability to perform critically needed complex detection—such as multiplexed detection for viral variant surveillance—may limit their widespread adoption. Herein, we developed a robust multiplexed CRISPR-based detection using LwaCas13a and PsmCas13b to simultaneously diagnose SARS-CoV-2 infection and pinpoint the causative SARS-CoV-2 variant of concern (VOC)— including globally dominant VOCs Delta (B.1.617.2) and Omicron (B.1.1.529)—all while maintaining high levels of accuracy upon the detection of multiple SARS-CoV-2 gene targets. The platform has several attributes suitable for POC use: premixed, freeze-dried reagents for easy use and storage; convenient direct-to-eye or smartphone-based readouts; and a one-pot variant of the multiplexed detection. To reduce reliance on proprietary reagents and enable sustainable use of such a technology in low- and middle-income countries, we locally produced and formulated our own recombinase polymerase amplification reaction and demonstrated its equivalent efficiency to commercial counterparts. Our tool—CRISPR-based detection for simultaneous COVID-19 diagnosis and variant surveillance which can be locally manufactured—may enable sustainable use of CRISPR diagnostics technologies for COVID- 19 and other diseases in POC settings.

## Introduction

The recurrent emergence of new severe acute respiratory syndrome coronavirus 2 (SARS-CoV-2) variants emphasizes the need of broad genomic and variant surveillance to monitor SARS-CoV-2 characteristics and evolution. Genomic and variant surveillance is critical to the understanding of the public health risk posed by new variants, as well as to the re-design and continual improvements of vaccines, therapeutics, and diagnostic technologies. Low- and middle-income countries (LMICs) are hotspots where several SARS-CoV-2 variants of concern (VOCs) are postulated to emerge from (Bi et al., 2022), yet LMICs are at a distinct disadvantage in ramping up variant surveillance capacity, due to resources being more limited, mismanaged, or both. The majority (77%) of LMICs sequenced a minuscule fraction (<0.5%) of their COVID-19 cases; 20 LMICs do not have any sequencing activity reported in public databases (Brito et al., 2021). Together, this calls for a robust and affordable genomic surveillance for LMICs.

Beyond genomic/variant surveillance activity, LMICs have limited access to the newest crisis management technologies; lack the capacity and infrastructure to manufacture and distribute crisis-critical products; and rely heavily on imports of these products which are subjected to constraints in the supply chain. Efforts to strengthen genomic/variant surveillance activity of LMICs in a sustainable manner—through the development of simple-to-use surveillance tools that can be locally manufactured–should be a global priority. Ideally, diagnostics and genomic/variant surveillance should be combined in the same technological platform to maximize disease detection and variant surveillance capacity while optimizing resource use.

Genetic surveillance of SARS-CoV-2 can either target the whole viral genome or specific genetic variations such as single nucleotide polymorphisms and indels through the use of next-generation sequencing or quantitative real-time PCR with reverse transcription (RT-qPCR) (European Centre for Disease et al., 2021). However, both sequencing- and PCR-based investigations could be slow in returning results, thereby delaying an effort to contain the outbreak especially for VOCs with greater transmissibility such as Delta (B.1.617.2; R_0_ of 5-7 (Burki, 2021; Liu & Rocklöv, 2021) and Omicron (B.1.1.529; R_0_ estimated to be as high as 10 (Burki, 2022)). Most importantly, current surveillance technologies require complicated, expensive instruments and expertise in handling and data analysis, resulting in the limited use of these technologies in LMICs (Brito et al., 2021) where disease detection and variant surveillance are critically needed.

The collateral activity of clustered regularly interspaced short palindromic repeats (CRISPR)- associated 12/13 (Cas12/13) nucleases (Chen et al., 2018; Gootenberg et al., 2018; Gootenberg et al., 2017; Harrington et al., 2018) has recently been used as a basis for nucleic acid detection, including that of SARS-CoV-2 (Broughton et al., 2020; Fozouni et al., 2020; Joung et al., 2020; Patchsung et al., 2020). CRISPR-based detection has many attributes suitable for use in LMICs especially in point-of-care settings: the assay does not need complicated equipment and provides fast-to-return results, while maintaining high levels of diagnostic accuracy and complex testing capabilities characteristic of laboratory-based diagnostic technologies. CRISPR-based detection is particularly amenable to multiplexing, due to orthogonal cleavage preferences of bystander nucleic acid probes by different Cas enzymes—particularly the Cas13 family—each of which can be programmed for sequence-specific detection by its corresponding crRNA (Gootenberg et al., 2018). Due to its high sequence specificity (when carefully designed and/or optimized), CRISPR-based detection has been explored as a tool to map key SARS-CoV-2 mutations. A few reported technologies use equipment-heavy PCR (He et al., 2022) and Fluidigm microfluidics (Welch et al., 2022) to amplify genetic materials and increase the assay throughput, respectively. While such technologies drastically enhance the ability to profile multiple mutations of SARS-CoV-2 and other viruses, the reliance on extensive equipment may exclude them from utility in resource-poor settings. Equipment-light CRISPR-based detection is considered highly practical for several reasons: these platforms often combine an isothermal amplification step—compatible with a simple heating apparatus like a water bath and obviating the need of a PCR thermocycler—with a CRISPR-Cas-based detection step for maximal sensitivity and specificity. While discrimination of SARS-CoV-2 VOCs can be achieved with POC CRISPR-based detection, current POC CRISPR-based assays have diminished sensitivity (a trade-off upon using a simplified protocol (Arizti-Sanz et al., 2021) and lack multiplexed detection capability (Arizti-Sanz et al., 2021; Fasching et al., 2022).

Here we develop multiplexed CRISPR-based detection with point-of-care characteristics to clinically diagnose SARS-CoV-2 infection and inform the causative VOCs in the same reaction, while maintaining high sensitivity of detection (Figure 1). To do so, we extensively optimized components and reaction conditions of the reverse-transcription recombinase polymerase amplification (RT-RPA) to enable multiplexed amplification of two SARS-CoV- 2 genes, *n* and *s* (Figure 1A, B). We explored several Cas13 enzymes including an engineered variant for multiplexed, orthogonal CRISPR-Cas13-based detection, and further screened different parameters for improved detection sensitivity. We lyophilized premixed amplification and CRISPR reactions and stored them in freeze-dried forms, which can be reconstituted simply through addition of buffered nucleic acid analytes. The freeze-dried reagent format enables convenient field deployment as well as longer reagent shelf-life. We further developed a highly sensitive one-pot reaction which combines multiplexed RT-RPA and Cas13-based detection in the same tube and demonstrated a simple visualization setup for multiplexed gene detection. Finally, our system was successfully validated in suspected SARS-CoV-2 samples from the real clinical setting; an indicative of both feasibility and robustness of our multiplexed Cas13-based assay.

**Figure 1.**
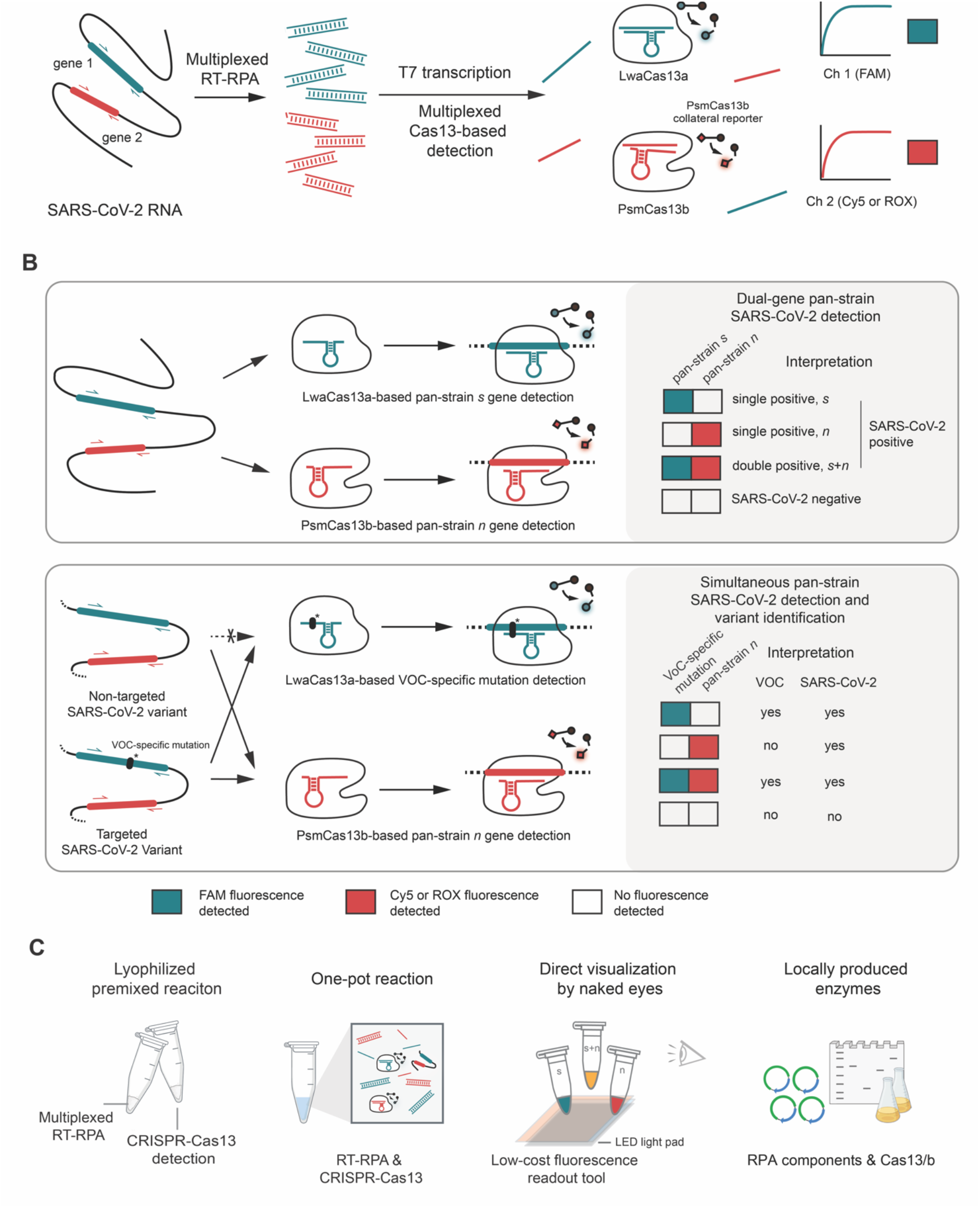
A multiplexed Cas13-based assay for simultaneous SARS-CoV-2 detection and variant identification. (A) Regions of interest in SARS-CoV-2 RNA, such as the *s* and *n* genes, are isothermally amplified by multiplexed RT-RPA. T7 transcription then converts and amplifies DNA amplicons to RNAs, which are recognized by cognate Cas13a-crRNA and Cas13b-crRNA complexes with orthogonal collateral cleavage preferences. Cleavage of orthogonal RNA reporters by target-activated Cas enzymes elicits multicolored fluorescence, which can be monitored with standard fluorescence detection instruments. (B) Top, dual-gene detection via CRISPR-Cas13a/b for COVID-19 diagnosis. Bottom, dual-gene detection for simultaneous COVID-19 diagnosis and SARS-CoV-2 variant identification. LwaCas13a is used to detect a universal, pan-strain region (top) or a variant- specific region (bottom) of the SARS-CoV-2 *s* gene. PsmCas13b is used to detect a pan- strain region of the *n* gene. The amplified regions of the S and N genes are shown in red and green respectively. (C) Developing simple-to-use SARS-CoV-2 variant surveillance tools which can be locally manufactured. Our multiplexed CRISPR-based detection reactions can be premixed and lyophilized for convenient use and storage; multiplexed RPA and Cas13-based reactions can be combined in one tube; result readouts can be directly observed by eye with low-cost light sources; and RPA and Cas13a/b can be locally produced and formulated.

After assessing cross-reactivity and the limit of detection (LoD), we clinically validated the multiplexed CRISPR-based detection of SARS-CoV-2 *n* and *s* genes (Figure 1B, top) on 136 clinical samples containing a full range of threshold cycle values (13-39). We found the multiplexed CRISPR-based detection of SARS-CoV-2 obtained from nasopharyngeal and throat swabs of infected patients to be 100% specific and 95-97% sensitive compared to RT-qPCR. Within the characterized LoD (Ct ∼37) the method is 100% specific and 100% sensitive. To allow for broader implementation of the technology during the pandemic, we submitted the most sensitive version of our freeze-dried, multiplexed CRISPR-based detection of SARS-CoV-2 RNA for technological evaluation with the Food and Drug Administration (FDA) of Thailand, and received full approval on September 22, 2021.

With emerging VOCs, we re-designed our multiplexed detection platform—with minimal adjustments to our FDA-approved protocol beyond new primers and crRNAs—to concurrently diagnose SARS-CoV-2 infection—via pan-variant detection of the *n* gene—and discriminate the highly transmissible Delta (B.1.617.2) and Omicron (B.1.1.529) via variant-specific detection of the *s* gene (Figure 1B, bottom). Our best design for multiplexed Delta variant detection showed excellent sensitivity (LoD at C_t_ ∼37) and specificity, with no cross-reactivity toward wild-type, Alpha (B.1.1.7), and Omicron SARS-CoV-2. Our Omicron variant detection—which detected the HV69-70 deletion also found in the now extinct Alpha strain— was slightly less sensitive (LoD at C_t_ ∼34) and exhibited minimal cross-reactivity with wild-type and Delta SARS-CoV-2.

To sustain SARS-CoV-2 detection and variant surveillance capability in LMICs, we also report here an RT-RPA reaction in which all protein components of RPA were locally produced in Thailand, and demonstrated similar amplification efficiency, including for multiplexed detection, of the locally produced RPA to commercial RPA. Our overall setup (Figure 1C)— an equipment-light, field-deployable, and easy-to-use multiplexed CRISPR-based assay capable of simultaneous detection of SARS-CoV-2 and VOC identification, while maintaining high specificity and sensitivity—can now be prepared with locally manufactured components, providing affordable access of critical reagents for diagnostic and variant surveillance assays to the most vulnerable populations.

## Results

### Developing multiplexed reverse-transcription RPA for SARS-CoV-2 RNA detection

Standard SARS-CoV-2 laboratory diagnostics include the detection of at least two loci, often in a multiplexed manner, as a precaution against escape mutations (Tang et al., 2020); we wished to first confer this multiplexed detection capability to our platform. We focused our optimization of multiplexed isothermal amplification on the RPA technique, as we and others (Bao et al., 2020) found another popular isothermal amplification technique, LAMP, to be highly susceptible to carryover contamination, likely due to its generation of large concatemer amplicons that are highly amplificative and resistant to degradation. RPA can also be performed at lower, near ambient temperature, making it more easily deployed for point-of-care or field applications. Our previous work identified four sets of RPA primer pairs which enable amplification of the spike (*s*), nucleocapsid (*n*), and two regions within the replicase polyprotein 1ab (*orf1ab*) genes of SARS-CoV-2 (Patchsung et al., 2020) (Figure 1‒figure supplement 1A). Their amplification rates were variable, with the primer pair detecting the *s* gene being the most sensitive (Patchsung et al., 2020). We found that simply combining two of these primer pairs did not result in amplification of both targets, as the fast-amplifying product of one gene would dominate the reaction (Figure 1‒figure supplement 1). In order to achieve simultaneous amplification of two targets, the amplification rates of both targets should match (Jauset-Rubio et al., 2018), and be as high as possible for maximal sensitivity. We decided to focus on improving the amplification of the *n* gene to match that of the *s* gene. The *n* gene has relatively fewer mutations over time (Dutta et al., 2020) and consistently provides the most sensitive detection in many nucleic acid-based detection assays due to its presence in many subgenomic RNA forms (Alexandersen et al., 2020). Screening of 12 RPA primer pair combinations identified a new *n* primer pair (F4-R1) with similar sensitivity of amplification to our *s* gene primers (Figure 1‒figure supplement 1B-C). Combining the optimized *n* and *s* primers in the same reverse-transcription RPA (RT-RPA) reaction containing SARS-CoV-2 RNA as a template allowed us to detect both *n* and *s* amplicons with similar efficiency (Figure 1‒figure supplement 1D). Beyond primer design, we optimized the primer concentration and fine-tuned the *n*:*s* primer pair ratio by increasing the *n* primer pair amount 1.25-1.5-fold, and found the *n* gene detection at these slightly skewed primer pair ratios to improve and match that of the *s* gene at all RNA concentrations tested (Figure 1‒figure supplement 2).

Consistent with previous reports (Patchsung et al., 2020; Qian et al., 2020), we found that RNase H improves RT-RPA efficiency, but reducing RNase H amount in RT-RPA 4-8 fold compared to what we initially reported (Patchsung et al., 2020) further enhanced the reaction (Figure 1‒figure supplement 2C). We reasoned that while RNase H can remove RNA from the RNA:DNA duplex product of RT and accelerate subsequent RPA, excess RNase H may degrade the SARS-CoV-2 RNA template in the starting RNA:primer complex and dampen the enzyme’s beneficial effect.

### Optimizing multiplexed CRISPR-based detection for SARS-CoV-2 RNA

We established a readout platform for multiplexed RNA amplification, via a multiplexed CRISPR-Cas reaction. Cas13 enzymes with orthogonal collateral cleavage preferences are well characterized; among the variants, Cas13a from *Leptotrichia wadei* (LwaCas13a) and Cas13b from *Prevotella* sp. MA2016 (PsmCas13b) have already been demonstrated to work well in tandem, and have been used in combination with multiplexed RPA to detect two synthetic DNA targets (Gootenberg et al., 2018). We previously set up LwaCas13a-based detection of SARS-CoV-2 RNA, primarily for the *s* gene, utilizing a FAM-labeled polyU reporter (Patchsung et al., 2020). To extend the detection to two genes, we expressed and purified PsmCas13b (Figure 1‒figure supplement 3), designed its CRISPR RNA (crRNA) to target the *n* gene of SARS-CoV-2, and prepared a Cy5-labeled polyA reporter to match with adenine cleavage preference of PsmCas13b.

We confirmed that PsmCas13b exhibits collateral cleavage of the polyA reporter, thereby eliciting Cy5 fluorescence when its *n* gene target is present. We empirically optimized the enzyme:crRNA amount used in the detection reaction, as well as the polyA reporter concentration (Figure 1‒figure supplement 4). Generally, more enzyme, crRNA, and the reporter are needed for the PsmCas13b-based detection compared to conditions used for LwaCas13a, reflecting poorer collateral cleavage kinetics of PsmCas13b, which was previously documented (Gootenberg et al., 2018) and which we also confirmed by performing side-by-side detection reactions with PsmCas13b vs LwaCas13a on the same RPA product (Figure 1‒figure supplement 5). Despite the slower kinetics of PsmCas13b, we demonstrated that PsmCas13b and LwaCas13a can function orthogonally in a multiplexed reaction, to detect *n* and *s* genes of SARS-CoV-2 respectively (Figure 1‒figure supplement 2D).

We also explored Cas13d from *Ruminococcus flavefaciens* (RfxCas13d), which is the most catalytically active of Cas13 orthologs in mammalian applications (Konermann et al., 2018), as well as its engineered variant, RfxCas13d-RBD, which has an RNA binding domain (RBD) from human protein kinase R fused to RfxCas13d to improve RNA binding (Yin et al., 2020). However, both RfxCas13d variants exhibited the polyU cleavage efficiency that was lower than that of LwaCas13a (Figure 1‒figure supplement 6). Therefore, we decided to proceed with the current best enzyme pair, LwaCas13a and PsmCas13b, and determine the system’s clinical performance in the multiplexed gene detection of SARS-CoV-2 (Figure 1‒figure supplement 6).

We screened solubility/stability-enhancing additives in the reactions to improve the efficiency of the multiplexed RT-RPA and CRISPR reaction. Triglycine boosted the multiplexed RPA reaction the most, consistently increasing the positive rates (Figure 1‒figure supplement 7A) and betaine monohydrate consistently increased the fluorescence signal generated from both Cas13a- and Cas13b-mediated cleavage by around 2-fold (Figure 1‒figure supplement 7B and Figure 1‒figure supplement 8). The beneficial effect of betaine in the Cas detection step can be combined with that of triglycine in the multiplexed RPA step, and we obtained the best signal-to-noise upon multiplexed detection when both additives were used in their respective reactions. (Figure 1‒figure supplement 7C). Empirical adjustments to the Cas:crRNA ratio and total amount, and reaction buffers further improved the multiplexed Cas reactions (Figure 1‒ figure supplement 9).

### Lyophilized RPA and CRISPR-Cas reaction pellets for portable SARS-CoV-2 RNA detection

We simplified liquid handling steps involved in the amplification and detection reactions and increased the shelf-lives of protein/RNA components of the reactions by storing them in lyophilized forms. All components required for either multiplexed RT-RPA or the CRISPR- LwaCas13a-based detection, aside fr6om potassium acetate and magnesium acetate for RT-RPA, and magnesium chloride for CRISPR-Cas reaction, can be lyophilized together in the presence of cryoprotective trehalose with minimal loss of activity upon reconstitution (Figure 1‒figure supplement 10 and Figure 1‒figure supplement 11). Trehalose may help the lyophilized RPA and Cas reactions through two mechanisms: it is known to help stabilize proteins for long-term storage and, from our additive screens, also helps boost the efficiency of both RPA and CRISPR-Cas13 reactions (Figure 1‒figure supplement 7A, B).

Freeze-dried RT-RPA pellet can simply be rehydrated by direct addition of the RNA extract from clinical samples, along with KOAc and Mg(OAc)_2_. The use of all-in-one freeze-dried RT-RPA pellets has an added benefit in that at least twice the volume of RNA can be used as input, resulting in even higher detection sensitivity compared to our conventional protocol where all reaction components are supplied in liquid form and the RNA input volume is restricted (Figure 1‒figure supplement 11). In addition, increasing the quantity of components in RPA pellets by adding another RPA pellet (RT-2xRPA) boosted the analytical sensitivity of detection in samples with C_t_ as high as 37 (Figure 1‒figure supplement 2E)

### Analysis of clinical specificity, analytical sensitivity (the limit of detection) and clinical sensitivity of the multiplexed detection of SARS-CoV-2 RNA

We performed clinical validation with a lyophilized premixed RT-2xRPA formulation, as it had the highest detection sensitivity and suitable features for POC use. We found that the multiplexed RPA and Cas13a/b-based detection are specific for the *s* and *n* genes of SARS-CoV-2, and exhibited no cross-reactivity upon using RNA input from other common human coronaviruses, including human coronavirus OC43 (hCoV-OC43), hCoV-NL63, and hCoV- 229E (Figure 2A). Using serially diluted RNA extracts from cultured SARS-CoV-2 in Vero cells (clinical isolate hCoV-19/Thailand/Siriraj_5/2020; GISAID accession ID: EPI_ISL_447908), we determined the limit of detection (LoD) of the optimized multiplexed detection of the SARS-CoV-2 *s* and *n* genes to be at C_t_ ∼37 (Figure 2B).

**Figure 2.**
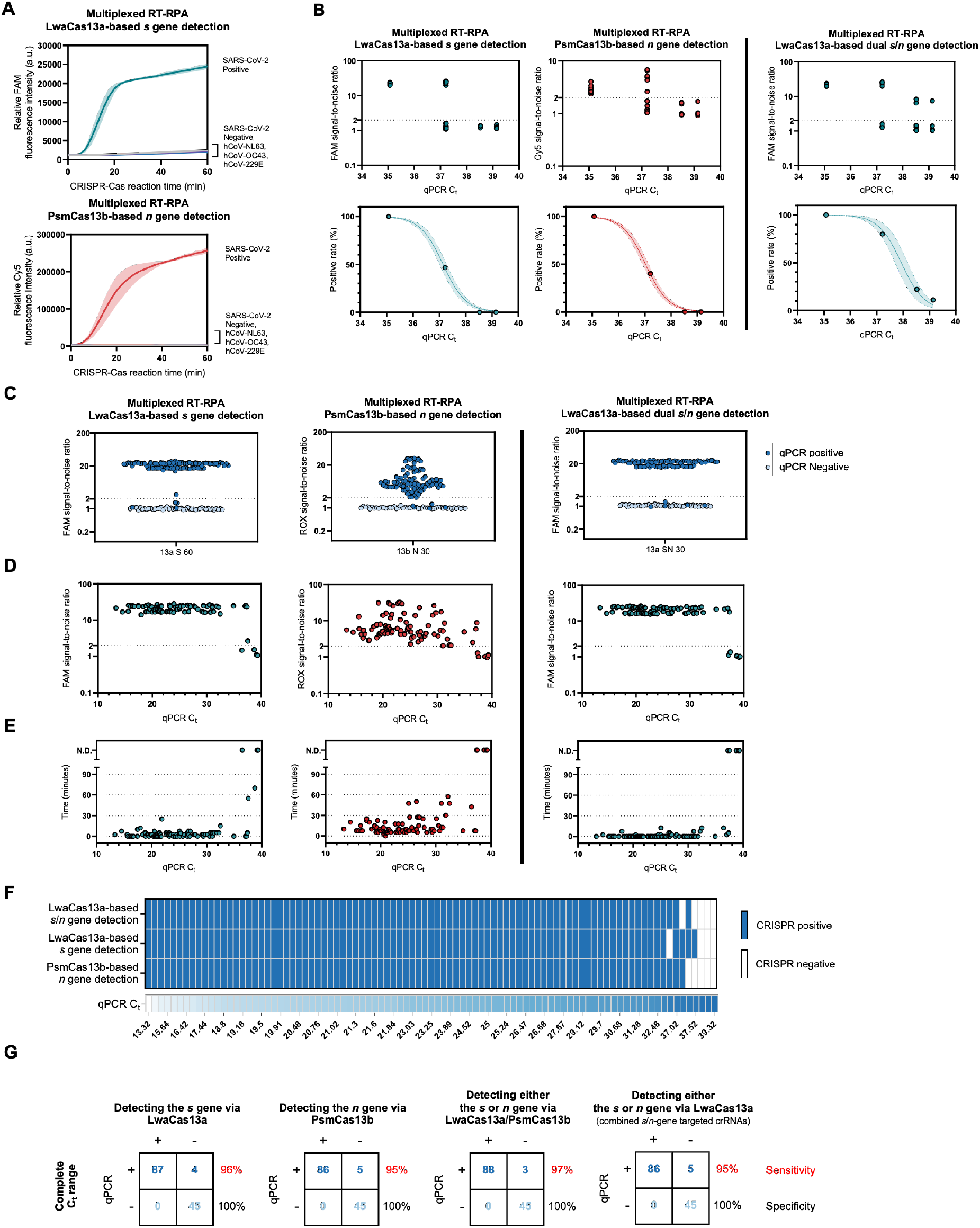
Specificity and sensitivity of the multiplexed CRISPR-based detection of SARS-CoV-2 RNA. **(A)** Clinical specificity. Kinetics of FAM (top) and Cy5 (bottom) fluorescence signal generation from the SARS-CoV-2 *s* (top) and *n* gene (bottom) detection via a multiplexed CRISPR-Cas reaction, using RNA extracts from clinical samples verified to be positive with different coronaviruses via RT-qPCR. The SARS-CoV-2 negative sample is an extract from a clinical sample confirmed to be SARS-CoV-2-negative by RT-qPCR. Data are mean ± s.d. from 3 replicates. **(B)** The limit of detection. Multiplexed RT-RPA followed by multiplexed Cas-based detection were performed with serially diluted SARS-CoV-2 RNA, whose Ct values were determined using a Luna one-step RT-qPCR assay targeting the *n* gene of SARS- CoV-2. At least three independent replicates of the amplification/detection reactions were performed for each SARS-CoV-2 RNA dilution. Top: signal-to-noises (S/N) of FAM (from LwaCas13a-mediated *s* gene detection) and Cy5 (from PsmCas13b-mediated *n* gene detection) fluorescence intensities are shown. The S/N threshold for a positive result was set at 2. Noise is defined as the fluorescence intensity generated from a negative sample with water as input performed in parallel. Bottom: positive rates of the detection of the *s* and *n* genes from multiplexed CRISPR-based detection at different SARS-CoV-2 RNA input concentrations. **(C)-(G)**, clinical sensitivity. **(C)** Summary of S/N obtained from multiplexed CRISPR-based detection results on 136 clinical samples (91 are COVID-19-positive by RT- qPCR; 45 are negative). **(D)** Relation of S/N obtained from multiplexed CRISPR-based detection to C_t_ values of COVID-19-positive samples. **(E)** Time to positive CRISPR detection results vs C_t_ values of COVID-19-positive samples. **(F)** Results of multiplexed CRISPR-based detection for 91 COVID-19-positive samples ordered by C_t_ values from RT- qPCR. **(G)** Concordance between RT-qPCR and multiplexed CRISPR-based detection of SARS-CoV-2 RNA. On the right of each concordance box, sensitivity values (%) for the detection of the *s* and *n* genes under a given scheme are shown in red; specificity values (%) are shown in black.

The clinical performance of the multiplexed detection platform on RNA extracts from 136 nasopharyngeal and throat swab clinical samples, 91 of which are SARS-CoV-2 positive by RT-qPCR (C_t_ of the *n* gene ranging from ∼13-39), matched well with the characterized specificity and LoD (Figure 2C-G). We were able to identify all 45 SARS-CoV-2-negative samples (100% specificity, Figure 2C). Among 91 RT-qPCR-positive samples, we were able to detect both *s* and *n* genes in 85 samples and at least either the *s* or the *n* gene in three other one sample, within 60 min of the multiplexed CRISPR reaction (Figure 2D, E). We were able to detect both genes in samples with C_t_ as high as 37.5 (Figure 2F). The clinical sensitivity of our multiplexed detection within the determined LoD (C_t_ ∼37) is 100% for the detection of either the *s* or *n* gene of SARS-CoV-2, and 95% for the detection of both genes (Figure 2G). For the full range of C_t_’s we encountered in clinical samples, the clinical sensitivity is 95-97% for at least one gene detected. Due to extensive optimizations, the limit of detection of our multiplexed detection scheme with lyophilized reagents are even higher than those of freshly prepared singleplex RPA and CRISPR reactions for SARS-CoV-2 *s* gene we previously reported (Patchsung et al., 2020) (LoD C_t_ ∼33.5).

In addition to performing multiplexed detection with LwaCas13a and PsmCas13b, we explored the use of LwaCas13a and crRNA combinations to detect both *s*- and *n*-gene RPA amplicons (Figure 2B-G, LwaCas13a-based dual S/N gene detection). In this scheme, the use of multiple primer pairs targeting different regions of the template could increase the probability of successful amplification by at least one primer pair, especially at very low template concentrations. Specific amplicons generated can all be detected by LwaCas13a—the Cas enzyme with highest collateral activity known to date—programmed with a mixture of crRNAs targeting all desired amplicons. Indeed, the clinical sensitivity for this combined output approach is better than that of the conventional multiplexed detection, reaching 95% for the full C_t_ range in our clinical samples, allowing for highly sensitive detection of SARS-CoV-2 RNA, while maintaining 100% specificity. The high level of detection accuracy was maintained upon testing with different SARS-CoV-2 variants (Figure 2‒figure supplement 1). This lyophilized, LwaCas13-based dual-gene detection of SARS-CoV-2 RNA served as a basis for our submission for technological evaluation with the Food and Drug Administration (FDA) of Thailand, who specified precise specificity (no cross-reactivity with Influenza A and B; MERS-CoV; and respiratory syncytial virus) and sensitivity (4,000 SARS-CoV-2 viral copies/mL) requirements for non-PCR molecular tests for SARS-CoV-2. Our test received full approval from the Thai FDA on September 22, 2021.

### A multiplexed CRISPR-based assay for simultaneous universal SARS-CoV-2 detection and SARS-CoV-2 variant differentiation

With suitable RPA primers and crRNA designs incorporating PAM requirements and synthetic mismatches, CRISPR-based detection should have the ability to differentiate single-nucleotide differences (Chen et al., 2021; Gootenberg et al., 2018; Gootenberg et al., 2017; Myhrvold et al., 2018; Ooi et al., 2021; Shinoda et al., 2021; Teng et al., 2019; Zhou et al., 2020) found in pathogen subtypes, including SARS-CoV-2 variants. In practice, cross-detection of highly similar sequences could not be eliminated; this reduced specificity necessitates the use of combinatorial detection of multiple signature mutations to provide high-confidence variant identification (Welch et al., 2022). Here, we sought to be judicious in our target sequence selection and crRNA design such that one CRISPR-Cas reaction is sufficient to identify a SARS-CoV-2 variant. A multiplexed CRISPR-based detection combining a pan-strain detection reaction with a variant differentiation one would be a cost-effective way to simultaneously diagnose COVID-19, and track the emergence and spread of a known VOC in a population.

In selecting target sequences for variant differentiation, we prioritized indel mutations (range 3-9 base-pairs per mutation) over substitution mutations (a single base-pair per mutation), as the number of bases affected by indel mutations likely result in greater number of mismatches which improves specificity for crRNA discrimination. However, the optimal indel length mismatch between a crRNA and its target is unclear; 1- to 5-nucleotide indels were linked to off-target activation of Cas9 (Lin et al., 2014), while a Cas13-based detection has been shown to tolerate up to 15-nucleotide deletion in the target sequence (Gootenberg et al., 2018). While sequence context likely plays a key role, short mismatches may still permit non-cognate crRNA: target recognition, and longer mismatches may allow formation of RNA bulges which minimally interfere with crRNA:target base pairing, also resulting in non-cognate target recognition and Cas activation. We thus decided to empirically test three different indel lengths when we selected SARS-CoV-2 variant-specific mutations: a three-nucleotide deletion (S: Y144Δ, specific for Alpha); two six-nucleotide deletions (*s*:HV69/70Δ, specific for Alpha and Omicron BA.1 and BA.3; and *s*:EF156/157ΔR158G; specific for Delta); and a nine-nucleotide deletion (*orf1ab*:SGF3675-3677Δ; specific for Alpha) (Figure 3A and Figure 3‒figure supplement 1). We also tested the detection of one single-nucleotide substitution (S:T22917G, resulting in L452R mutation; specific for Delta) and incorporated a synthetic mismatch in our crRNA design (Figure 3‒figure supplement 2).

**Figure 3.**
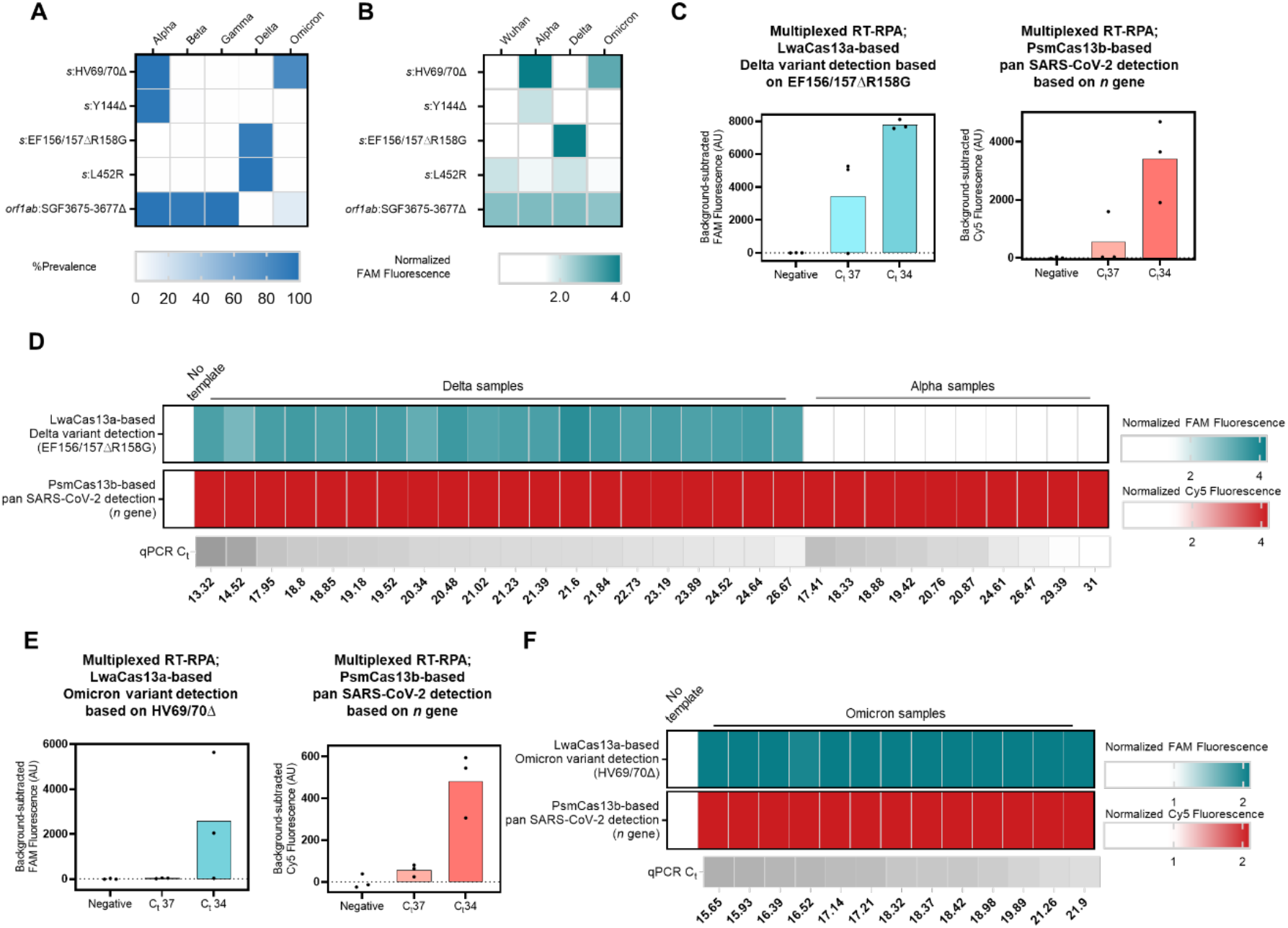
Multiplexed CRISPR-based detection for SARS-CoV-2 variant surveillance. (**A**) Prevalence of the target mutations in five major SARS-CoV-2 variants of concern (obtained from https://outbreak.info on, 15 March 2022, Alpha (n = 1,155,468), Beta (n = 41,428), Gamma (n = 120,775), Delta (n = 4,239,814), and Omicron (n = 2,150,574)) (**B**) Selectivity of singleplexed CRISPR-based SARS-CoV-2 variant detection targeting different mutations of interest. FAM fluorescence intensities at 60 minutes were normalized against intensities obtained from the no template control. Data are mean of two replicates. Raw kinetic traces are shown in Figure 3‒figure supplement 4 **(C)** Multiplexed CRISPR-based detection for the Delta variant. The Delta *s* gene was detected via its EF156/157ΔR158G mutation and LwaCas13a. Pan-strain SARS-CoV-2 detection was performed via PsmCas13b-based detection of a conserved region in the *n* gene. Relative FAM (from LwaCas13a-mediated *s* gene detection) and Cy5 (from PsmCas13b-mediated *n* gene detection) fluorescence intensities are shown. Data are mean ± s.d. from 3 replicates. Raw kinetic traces are shown in Figure 3‒figure supplement 7A **(D)** Clinical performance of the multiplexed CRISPR-based detection for the Delta variant. Each cell represents a COVID-19-positive clinical sample from Delta (left, n = 20) or Alpha (right, n = 10) lineage. FAM and Cy5 fluorescence intensities at 60 minutes were normalized against intensities obtained from the no template control. **(E)** Multiplexed CRISPR-based detection for the Omicron variant. The Omicron *s* gene was detected via its HV69/70Δ mutation and LwaCas13a. Pan-strain SARS-CoV-2 detection was performed via PsmCas13b-based detection of the *n* gene. Relative FAM (from LwaCas13a-mediated *s* gene detection) and Cy5 (from PsmCas13b-mediated *n* gene detection) fluorescence intensities are shown. Data are mean ± s.d. from 3 replicates. Raw kinetic traces are shown in Figure 3‒figure supplement 7B **(F)** Clinical performance of the multiplexed CRISPR-based detection for Omicron. FAM and Cy5 fluorescence intensities at 30 minutes were normalized against intensities obtained from the no template control.

We designed corresponding RPA primers and LwaCas13a crRNAs for all five mutation-specific CRISPR-based detection (Figure 3‒figure supplement 2 and Figure 3‒figure supplement 3), and first tested their specificity (Figure 3B, Figure 3‒figure supplement 4). The five SARS-CoV-2 variant-specific mutation detection reactions showed SARS-CoV-2 RNA-dependent signal but exhibited different levels of selectivity for their target variants. The two six-nucleotide deletion detections performed the best: EF156/157ΔR158G was highly specific for Delta, with no cross-reactivity with Wuhan, Alpha, or Omicron strains; HV69/70Δ was also highly specific for Alpha and Omicron as designed, but showed low-level cross-reactivity with Wuhan and Delta. Y144Δ, SGF3675-3677Δ and L425R showed clear detection preference for Alpha, Alpha, and Delta respectively, but background signals from other strains were significant. We thus proceeded with the two best-performing variant-specific detection reactions—EF156/157ΔR158G and HV69/70Δ—for subsequent experiments.

We characterized the limit of detection of the singleplex EF156/157ΔR158G-based detection using a dilution series of RNA extracted from cultured SARS-CoV-2 Delta strain, and found the detection to have an excellent LoD, capable of detecting Delta RNA with C_t_ of up to ∼38 (Figure 3‒figure supplement 5A). Further evaluation revealed robust detection in clinical samples (C_t_ 15-37) (Figure 3‒figure supplement 5B). We then combined the LwaCas13a-based Delta-specific detection of the *s* gene with the PsmCas13b-based pan-SARS-CoV-2-strain detection of the *n* gene in a multiplexed format (Figure 1B). Using the premixed, lyophilized and multiplexed formulation as previously described, we confirmed that the excellent LoD of Delta-specific reaction was maintained in the multiplexed format (Figure 3C and Figure 3‒ figure supplement 6). Furthermore, the LwaCas13a-based Delta-specific multiplexed detection was able to correctly identify all of twenty Delta patient specimens (C_t_ 13-27), with no cross-reactivity toward ten Alpha clinical specimens (C_t_ 17-31) (Figure 3D). The PsmCas13b-based pan-strain detection was fully functional, detecting the presence of SARS-CoV-2 in this same set of thirty clinical samples.

With the recent emergence of the Omicron variant, we explored the use of the HV69/70Δ- based detection for Omicron identification. We also performed additional cross-reactivity evaluation with higher RNA titer at C_t_ ∼25 since we observed sporadic cross-reactivity in the previous experiment (Figure 3B and Figure 3‒figure supplement 7). We found that the HV69/70Δ-based detection was able to distinguish Alpha and Omicron variants from other variants even at higher RNA titer. Next, we demonstrated that the singleplex HV69/70Δ-based detection was sensitive to RNA with C_t_ up to ∼34 upon testing with a dilution series of Omicron variant RNA (Figure 3‒figure supplement 5C) and was able to detect all Omicron variant clinical samples (Figure 3‒figure supplement 5D). Using serially diluted RNA from an Omicron clinical extract, we demonstrated sensitive detection of the Omicron variant at up to Ct ∼34 in a multiplexed format (LwaCas13-based detection of Omicron HV69/70Δ *s* signature, and PsmCas13b-based pan-strain *n* gene detection) (Figure 3E, Figure 3‒figure supplement 6). In addition, we were able to detect the HV69/70Δ *s* mutation as well as the SARS-CoV-2 *n* gene in fifteen Omicron clinical specimens (C_t_ 16-22), using the HV69/70Δ-based multiplexed detection approach (Figure 3F).

### Combining multiplexed RPA and Cas13-based detection in a single step

To simplify usage and minimize the risk of carryover contamination, we formulated a one-pot protocol which combined components of multiplexed RPA and Cas13-based detection in a single reaction tube. We first tested a previously reported one-pot SHINE protocol (Arizti-Sanz et al., 2020) for the detection of SARS-CoV-2 S gene, but saw that its analytical sensitivity is poorer than our conventional two-step amplification and detection in separate pots by ∼10-100- fold. Realizing that concentrations of RPA components in SHINE are diluted by 2-fold compared to the two-step variant, potentially hampering performance of RPA in the one-pot reaction, we re-formulated a one-pot recipe in which final concentrations of all components are as close to those in our optimized two-step variant as possible. We also switched the reaction temperature to be 39 °C, found to be optimal for combined RPA and Cas13-based detection (Figure 4‒figure supplement 1). We tested the optimized one-pot multiplexed RPA amplification of the *s* and *n* genes and concurrent detection by LwaCas13a programmed with *s*- and *n*-targeted crRNAs, and found its analytical sensitivity to be on par with the two-step protocol (Figure 4A).

**Figure 4.**
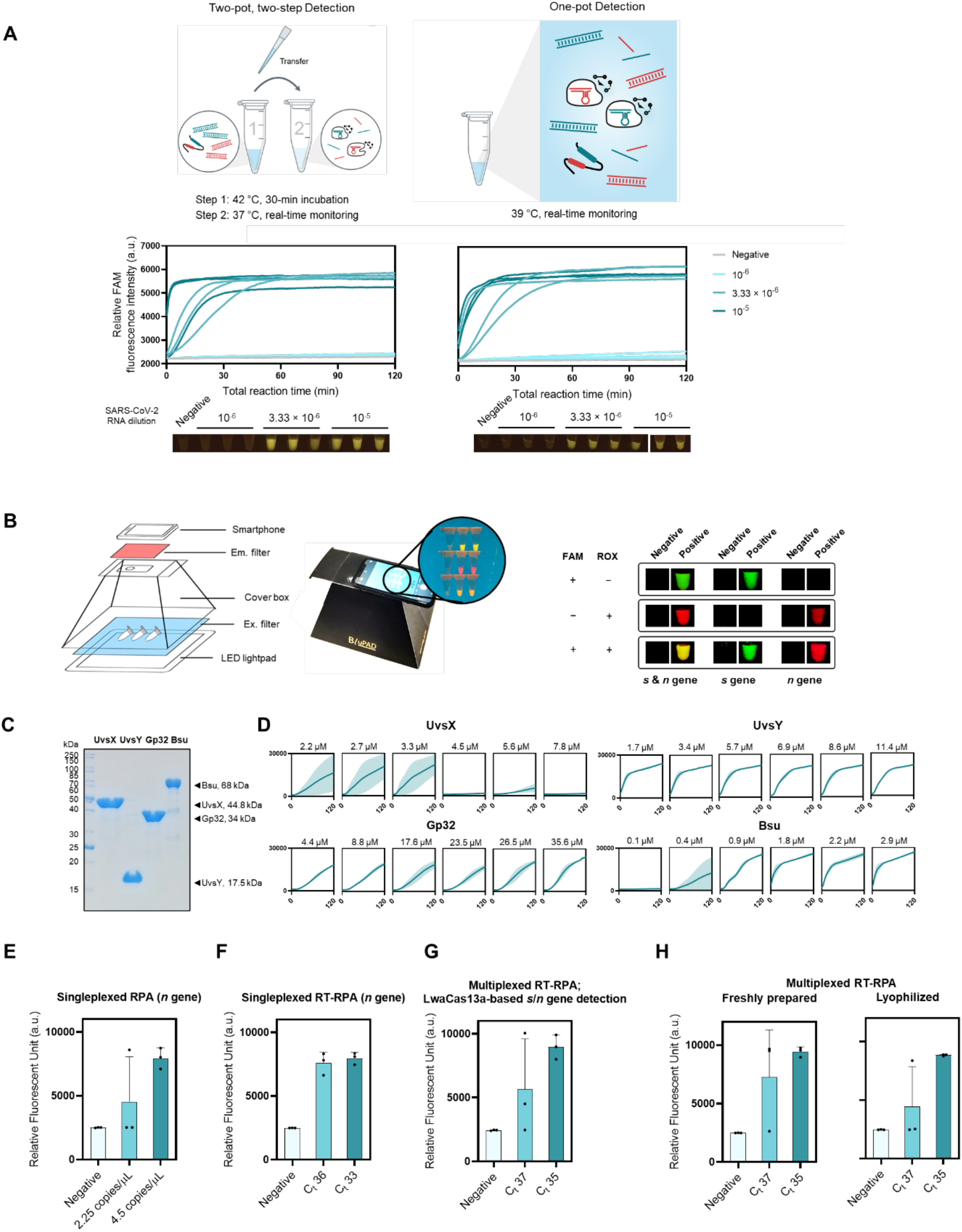
One-pot formulation, easy visualization, and local production of components for RPA and multiplexed CRISPR-based detection. (**A**) The one-pot reaction scheme and its analytical sensitivity compared to the standard two-pot setup. For all, *s*- and *n*-gene amplicons generated from multiplexed RPA were detected with LwaCas13a-based reaction programmed with *s*- and *n*-targeted crRNAs. Data from 3 independent replicates at each RNA input dilution are shown. (**B**) Easy visualization of multiplexed Cas13-based detection. A sample equipment setup with a LED transilluminator, appropriate lighting gels for visualization of fluorescein (FAM) and rhodamine X (ROX), and a smartphone for image capturing is shown. Smartphone-captured images of FAM and ROX signal generated from multiplexed Cas13a/b-based detection of the *s* and *n* genes of SARS-CoV-2. (**C**) SDS-PAGE analysis of locally produced and purified RPA components: UvsX, UvsY, Gp32, and Bsu polymerase large fragment (Bsu LF). (**D**) Optimizations of protein concentrations in in-house RPA. RPA reactions with varying concentrations of UvsX, UvsY, Gp32, and Bsu LF were performed with pUC57-2019-nCoV-N plasmid as a DNA template (at 10,000 copies/µl). Thereafter, the RPA products were used in a LwaCas13a-based detection. FAM fluorescence generated over 120 min for each RPA condition is shown. (**E, F**) Optimally formulated in-house RPA in amplification of N gene of SARS-CoV-2 from pUC57-2019-nCoV-N plasmid (**E**) and serially diluted SARS-CoV-2 RNA (**F**). LwaCas13a-based detection was then used to detect *n*-gene amplicons, with FAM fluorescence intensity generated after 90 min at 37°C shown. (**G**) Optimized in-house RT-RPA for multiplexed detection. Multiplexed RT-RPA to amplify the *s* and *n* genes of SARS-CoV-2 was performed using serially diluted SARS-CoV-2 RNA as a template. Detection of *s* and *n* amplicons was monitored through FAM fluorescence signal generated by Cas13a- mediated *s* and *n* gene detection. The reaction was measured after 90 min at 37°C. (**H**) Lyophilized multiplexed in-house RT-RPA has similar sensitivity as freshly prepared reactions. Amplification and detection of the SARS-CoV-2 *s* and *n* gene are as in (**G**). Data are mean ± s.d. from 3 replicates. RNase-free water was used as input of all negative control reactions.

### Direct visualization of multiplexed Cas13-based detection

To facilitate the use of a multiplexed CRISPR reaction in point-of-care settings, we switched the fluorophore labels of the PsmCas13b-polyA reporter, from Cy5 to rhodamine X (ROX). The ROX-based polyA reporter generates slightly higher levels of signal-to-noise upon PsmCas13b-mediated cleavage compared to the Cy5-based reporter (Figure 4‒figure supplement 2). Moreover, ROX and fluorescein (FAM, on the LwaCas13a-polyU reporter) can be efficiently excited using a single blue LED light source(Ball et al., 2016), under which Cy5 gives no signal. ROX and FAM emissions can then be simultaneously monitored using different combinations of colored plastic filters (Figure 4B, Figure 4‒figure supplement 3 and Figure 4‒figure supplement 4). The emissions are easily readable by eye and ultimately, can be coupled to a portable smartphone-based detection (Priye et al., 2018; Priye et al., 2017; Samacoits et al., 2021) for quantitative analysis.

### Establishing a fully configurable RPA reaction

Given the bottleneck experienced by many labs in procuring and affording commercial RPA, we sought to produce protein components and formulate efficient RPA in house. We expressed the four main protein components of the RPA reaction—bacteriophage T4 recombinase UvsX (UvsX), bacteriophage T4 recombinase loading factor UvsY (UvsY), bacteriophage T4 single-stranded binding protein Gp32 (Gp32), and *Bacillus subtilis* DNA Polymerase I, large fragment (Bsu LF)—in *Escherichia coli* and purified them to about 95% purity (Figure 4C). The activity of our laboratory-made RPA, formulated using a previously reported condition (Piepenburg et al., 2006), was tested via the amplification of a synthetic DNA representing the *s* gene of SARS-CoV-2, which showed clear amplification across different input concentrations under this standard condition (Figure 4‒figure supplement 5). We next titrated the concentrations of the four main protein components of the RPA reaction in order to improve the amplification and found optimal concentrations for each component as follows: 2–3 µM UvsX; 2–11 µM UvsY, 26–35 µM Gp32, and 2-3 µM Bsu LF (Figure 4D). We combined optimized protein concentrations with the triglycine additive to successfully amplify the *n* gene of SARS-CoV-2 from either DNA or RNA samples for subsequent CRISPR-Cas13a detection (Figure 4E-4F). The optimized in-house RT-RPA can be used in a multiplexed amplification reaction of the *s* and *n* genes, coupled to LwaCas13a-based detection with *s*- and *n*-targeted crRNAs (G) and can be lyophilized without loss in detection sensitivity (Figure 4H).

## Discussion and outlook

Many advances in point-of-care nucleic acid detection technologies for SARS-CoV-2 RNA— from crude lysate preparation (Arizti-Sanz et al., 2020; Joung et al., 2020; Qian et al., 2020), nucleic acid enrichment post-lysis to enhance sensitivity (Joung et al., 2020), multiple options of isothermal amplification (Subsoontorn et al., 2020), amplification-free detection (Fozouni et al., 2020), multiple modes of detection (Chaibun et al., 2021; Patchsung et al., 2020) to mobile, automated readouts of results (Fozouni et al., 2020)—have collectively pushed toward large-scale practical uses of these technologies for the surveillance of the ongoing pandemic. Here, we extend the utility of CRISPR-based detection—already shown to be among the most sensitive and specific isothermal nucleic acid tests—toward robust multiplexed detection of SARS-CoV-2 RNA. Multi-parameter optimizations of multiplexed RPA and CRISPR-Cas reaction resulted in a highly specific and sensitive detection platform for the *s* and *n* genes of SARS-CoV-2. We critically evaluated its performance on a large set of clinical samples and demonstrated the high sensitivity (95-97%) and specificity (100%) of the multiplexed CRISPR-based detection of SARS-CoV-2 RNA. Our enhanced multiplexed CRISPR-based reaction can be re-designed to simultaneously provide COVID-19 diagnosis and identify the causative SARS-CoV-2 variant of concern, including Delta and Omicron, with high specificity and sensitivity. Beyond variant surveillance, our platform can be used for other multiplexed combinations, including the detection of one SARS-CoV-2 gene and a human gene/RNA control, and the detection of SARS-CoV-2 with another virus, such as influenza, whose infection can co-occur with COVID-19 and present similar symptoms (Mayuramart et al., 2021).

Beyond complex detection, we also improve the usability and portability of the enhanced multiplexed CRISPR-based detection platform through freeze-dried reaction mixtures and a one-pot configuration. While the current sensitivity of our enhanced multiplexed platform is already high, we think the key to further improve the test accuracy while maintaining the ease of use of the detection technology lies in the ability to tinker with the RPA reaction itself. The ability to manufacture this key reagent at high quality in the lab has enabled us to make progress toward this goal, first by improving the ease of use through tailored lyophilization of reaction components. In addition to increasing sensitivity, engineering RPA such that the reaction can function directly and robustly in diverse biological fluids could make RPA and CRISPR-based detection viable testing platforms in very limited-resource settings—even at home—for currently circulating or newly emerging SARS-CoV-2 and other infectious agents. Beyond improving technology characteristics, local manufacturing of test kit components creates opportunities for economic and research development, which could lead to sustainable use of these technologies in disease monitoring and diagnosis especially in developing countries.

Both RPA and CRISPR-Cas reactions contain primarily enzyme components; in this study, we started to explore the use of an engineered Cas13 enzyme to improve the reactions. Continual efforts to engineer these enzyme components and their substrates as recently demonstrated with Cas12a (Nguyen, Rananaware, et al., 2020) and its crRNA (Nguyen, Smith, et al., 2020), along with integration with automation platforms for high-throughput assay development, could pave way for substantially improved detection platforms.

## Materials and Methods

### Cloning methods for RPA expression plasmids

Nucleic acid sequences codon optimized for *E. coli* expression for T4 UvsX, T4 UvsY, T4 gp32, and Bsu DNA polymerase large fragment (Bsu LF) were ordered as plasmids from GeneArt (ThermoFisher Scientific). Bsu LF and *uvsY* were cloned into the pET28a backbone using Circular Polymerase Extension Cloning (CPEC) (Quan & Tian, 2011). *Gp32* and *uvsX* were cloned into the pET28a backbone using Type IIS assembly (Engler et al., 2008). Plasmids were sequence verified (Eurofins Genomics) and are available on Addgene as follows: pET28a-MH6-Bsu LF (Plasmid #163911); pET28a-gp32-H6 (Plasmid #163912); pET28a- uvsX-H6 (Plasmid #163913); and pET28a-MH6-uvsY (Plasmid #163914).

### Expression and purification of protein components of RPA

We used near-identical expression and cell lysis protocols for UvsX, UvsY, gp32, and Bsu LF, but the purification step and final storage buffer conditions were different for each enzyme, as given below.

An expression plasmid for UvsX, UvsY, gp32, or Bsu LF was transformed into *E. coli* BL21(DE3) cells. The cells were grown in LB medium at 37°C until OD_600_ reached 0.7–0.8. Protein expression was induced using the following concentrations of isopropyl-β-D-thiogalactopyranoside (IPTG): 1 mM for UvsX and UvsY; 0.2 mM for gp32; and 0.5 mM for Bsu LF. The cells were grown for additional 16 hours at 16°C and harvested by centrifugation. The cell pellet was resuspended in lysis buffer (50 mM sodium phosphate pH 8.0, 500 mM sodium chloride, 10 mM imidazole). Phenylmethylsulfonyl fluoride (PMSF) was added into resuspended solution at 1 mM final concentration followed by sonication for a total burst time of 3 minutes (3 sec on for short burst and 9 sec off for cooling). The cell lysate was clarified by centrifugation at 27,000×g for 30 minutes at 4 °C. Nickel-nitrilotriacetic acid (Ni-NTA) agarose beads (Qiagen) was washed with two column volumes of water and equilibrated with lysis buffer. The clarified cell lysate was incubated with the Ni-NTA agarose beads (Qiagen) at 4 °C for 40 minutes. The column was washed with ten column volumes (CVs) of washing buffer (50 mM sodium phosphate pH 8.0, 500 mM sodium chloride, and 20 mM imidazole), and the bound proteins were eluted with elution buffer (50 mM sodium phosphate pH 8.0, 500 mM sodium chloride, 250 mM imidazole).

For UvsX, the protein was further purified by Heparin Sepharose Fast Flow (GE Healthcare Life Sciences) with a linear gradient of 0.1–1 M NaCl in Buffer A (20 mM Tris-HCl pH 8, 5 mM β-mercaptoethanol (βME)). Proteins were concentrated and stored in 20 mM Tris-HCl pH 8.0, 500 mM NaCl, and 40% (v/v) glycerol at −20 °C.

For UvsY, the protein was further purified by Heparin Sepharose Fast Flow (GE Healthcare Life Sciences) with a linear gradient of 0.1–1 M NaCl in Buffer A (20 mM Tris-HCl pH 8, 5 mM βME). Afterward, proteins were dialyzed against 20 mM Tris-HCl pH 7.5, 400 mM NaCl, 20% (v/v) glycerol and 5 mM βME, and the glycerol concentration adjusted to be 50% (v/v) with the addition of UvsY freezing buffer (20 mM Tris-HCl pH 7.5, 400 mM NaCl, and 80% (v/v) glycerol). Protein aliquots were stored in 20 mM Tris-HCl pH 7.5, 400 mM NaCl, and 50% (v/v) glycerol at −20 °C.

For gp32, eluted protein fractions were dialyzed against 20 mM Tris-HCl pH 7.5, 400 mM NaCl, and 5 mM βME, and the glycerol concentration adjusted to be 50% (v/v). Protein aliquots were stored in 10 mM Tris-HCl pH 7.5, 200 mM NaCl, and 50% (v/v) glycerol at −20 °C.

For Bsu LF, eluted protein fractions were concentrated and dialyzed against 50 mM Tris-HCl pH 7.5, 50 mM KCl, 1 mM DTT, 0.1 mM EDTA, and 20% (v/v) glycerol. Glycerol concentration was adjusted to be 50% (v/v) with the addition of Bsu LF freezing buffer (50 mM Tris-HCl pH 7.5, 50 mM KCl, 1 mM DTT, 0.1 mM EDTA, and 90% (v/v) glycerol). Protein aliquots were stored in 50 mM Tris-HCl pH 7.5, 50 mM KCl, 1 mM DTT, 0.1 mM EDTA, and 50% (v/v) glycerol at −20 °C.

### Expression and purification of Cas enzymes

#### LwaCas13a expression and purification

LwaCas13a was expressed from pC013-His_6_-Twinstrep-SUMO-LwaCas13a (Addgene: #90097) and purified as previously described (Patchsung et al., 2020).

#### PsmCas13b expression and purification

*Escherichia coli* BL21 (DE3) transformed with pC0061-His_6_-Twinstrep-SUMO-PsmCas13b (Addgene: #115211) were grown in LB media containing 25 µg/mL ampicillin, and the protein expression induced by the addition of 500 µM IPTG at 16 °C for overnight. Cells were collected by centrifugation at 8,000 rpm for 20 min at 4 °C and the supernatant was discarded. The cell pellet was resuspended in extraction buffer (50 mM Tris-HCl, 500 mM NaCl, 1 mM DTT, 1x protease inhibitor cocktail, 0.25 mg/mL lysozyme, 5 mM imidazole, pH 7.5) then lysed by sonication (Sonics Vibracell VCX750) using 40-60% pulse amplitude (on 10 s and off 20 s until completely lysed). The lysate was centrifuged at 15,000 rpm for 45 min at 4 °C, and the soluble fraction filtered through a 0.45 µm polyethersulfone membrane and loaded into Chelating Sepharose^TM^ Fast Flow column which was pre-loaded with 0.2 M NiSO_4_ and pre-equilibrated with binding buffer (50 Tris-HCl, 500 mM NaCl, 5 mM imidazole pH 7.5). The column was washed with 5 column volumes of binding buffer, 6 column volumes of washing buffer (50 Tris-HCl, 500 mM NaCl, 50 mM imidazole pH 7.5) and eluted with 6 column volumes of elution buffer (50 Tris-HCl, 500 mM NaCl, 500 mM imidazole pH 7.5). The fractions containing His-Twinstrep-SUMO-PsmCas13b were pooled and exchanged with SUMO cleavage buffer (20 mM Tris-HCl, 150 mM NaCl, pH 8.0) at 4 °C overnight.

The SUMO tag was cleaved with Ulp1 SUMO protease (Addgene #64697) using 10:1 of SUMO substrate by incubation at room temperature for 5 h. The reaction was adjusted to pH 6.0 and filtered with 0.45 µm polyethersulfone membrane before loading into HiTrap SP HP column which was equilibrated with binding buffer (50 mM phosphate pH 6.0, 200 mM NaCl). The protein was washed and eluted with 50 mM phosphate, 200 mM NaCl with pH ranging from 6.0, 6.5, 7.0, 7.5, and 8.0, followed by 50 mM Tris pH 8.0, 1 M NaCl. The PsmCas13b fractions were pooled, concentrated, exchanged buffer using centricon 30k, diluted to a concentration of 1.2 mg/mL, and stored at -80 °C in storage buffer (50 mM Tris-HCl, 600 mM NaCl, 5 % glycerol, 2 mM DTT).

#### RfxCas13d and RfxCas13d-RBD expression and purification

*E. coli* BL21 (*DE3*) cells transformed with CMS1371 (for His_6_-MBP-RfxCas13d expression) or CMS1372 (for His_6_-MBP-RfxCas13d-dsRBD) plasmid were grown in LB media containing 100 µg/mL ampicillin, and the recombinant gene expression induced by the addition of 500 µM IPTG at 16 °C for overnight. Cells were collected and lysed as for PsmCas13b. Following lysate clarification, the soluble fraction was filtered through 0.2 µm polyethersulfone membrane and purified by HisTrap FF column connected to FPLC system. The column was pre-equilibrated with binding buffer (50 mM Tris-HCl, 500 mM NaCl, 5 mM imidazole; pH 7.5). The soluble fraction was loaded at 2 mL/min, washed with 5 column volumes of binding buffer. The recombinant protein was eluted in linear gradient of elution buffer (50 mM Tris-HCl, 0.5 M NaCl, and 0.5 M imidazole; pH 7.5). The eluted fractions were pooled and dialyzed with 50 mM Tris-HCl, 0.5 mM EDTA, 1 mM DTT and 200 mM NaCl; pH 7.5 at 4 °C for 3 hours, before proceeding to MBP tag removal.

The MBP tag was cleaved with ultraTEV protease using 20:1 substrate: protease ratio in 50 mM Tris-HCl, 0.5 mM EDTA, 1 mM DTT and 200 mM NaCl; pH 7.5. The reaction mixture was incubated at 4 °C for overnight with gentle shaking before applied onto HisTrap column, which was pre-equilibrated with (50 mM Tris-HCl, 500 mM NaCl, 5 mM imidazole; pH 7.5). The flow-through and unbound fractions containing RfxCas13d and RfxCas13d-dsRBD protein were then collected while retained MBP and other contaminants were later washed out with high concentration of imidazole. The purified protein was exchanged against 40 mM Tris, 400 mM NaCl; pH 7.5 in DEPC-treated water and concentrated using centricon 30k, and diluted to a concentration of 1.22 mg/mL (for RfxCas13d) and 1.89 mg/mL (for RfxCas13d- RBD).

### RPA primers, crRNAs, and RNA reporters

The oligonucleotides used are listed in Table S2. The crRNAs used for variant detection were synthesized via *in vitro* transcription. The T7-3G oligonucleotide (at the final concentration of 0.5 µM) and the ssDNA crRNA template (at the final concentration of 0.5 µM) were subjected to 34 cycles of 50 μL Q5^®^ High-Fidelity DNA Polymerase reaction (New England Biolabs). PCR products were purified using DNA Clean & Concentrator-5 kits (Zymo Research) and eluted in 20 μL of nuclease-free water. Four picomoles of the purified dsDNA crRNA template were used in *in vitro* transcription reactions using RiboMAX Large Scale RNA Production System–T7 (ProMega) or MEGAshortscript™ T7 Transcription Kit (Invitrogen). The reactions were performed at 37 °C overnight, treated with DNase I, and purified using phenol-chloroform extraction and alcohol precipitation. 15% acrylamide/7.5 M urea PAGE with GelRed^®^ (Biotium) staining was used to assess the size and purity of the transcribed product. Gels were imaged on ImageQuant™ LAS 4000 (GE Healthcare); quantifications of produced crRNAs were performed by measuring band densitometries using Fiji ImageJ software (Schindelin et al., 2012) and comparing to RNA standards.

### Design of RPA primers and crRNAs

#### General RPA primers and crRNAs design

RPA primers and crRNA were designed according to a published protocol (Kellner et al., 2019).

#### Design of RPA primers and crRNA targeting specific SARS-CoV-2 mutations

Representative genome sequences of Alpha (n = 1,100), Beta (n = 126), Gamma (n = 729), and Delta (n = 1,329) were retrieved from NCBI SARS-CoV-2 data packages using PANGO designations (Rambaut et al., 2020) as queries (dates of data retrieval, 27 April and 5 May 2021). Retrieved sequences were aligned using MAFFT (Katoh & Standley, 2013). The alignments were visualized and used to create consensus sequence for each variant using Jalview (Waterhouse et al., 2009). The four consensus sequences along with the SARS-CoV- 2 isolate Wuhan-Hu-1 NCBI reference genome (Accession ID NC_045512.2) were re-aligned using MAFFT and visualized in SnapGene software (Insightful Science). Using visualized multiple sequence alignment, RPA primers and crRNAs were manually designed to cover regions spanning the target mutation while avoiding regions with genetic variations among SARS-CoV-2 variants. The design and chosen regions are shown in Figure 3‒figure supplement 2 and Figure 3‒figure supplement 3.

#### Exclusivity evaluation of designed crRNAs

crRNA spacer sequences and their complementary sequences were searched against a BLAST database of Betacoronavirus nucleotide sequences (Betacoronavirus BLAST, https://blast.ncbi.nlm.nih.gov/Blast.cgi?PAGE_TYPE=BlastSearch&BLAST_SPEC=Betacoronavirus), with the maximal number of target sequences set to 5000. The results were filtered to include complete genomes (>29,000 nt) with 90-100% query coverage and 90-100% identity (date of data retrieval 1 March 2022). The clade assignment of genomes was done using NextClade (https://clades.nextstrain.org) (Aksamentov et al., 2021). The genomes with bad and mediocre NextClade overall quality control status were excluded. The analysis was summarized in Figure 3‒figure supplement 8. The associated dataset was available in Figure 3–source data 1.

#### Inclusivity evaluation of designed primers and crRNAs

GenBank accession lists of SARS-CoV-2 variant genomes sampling by the NextStrain COVID-19 global analysis open-data build were acquired by applying filter data by clade before downloading author metadata (date of data retrieval, 3 March 2022); Alpha (n = 110), Beta (n = 30), Gamma (n = 31), Delta (n = 1,044), and Omicron (n = 785)). FASTA files comprising listed genomes were downloaded from GenBank and aligned using MAFFT. The resulted alignments were visualized and inspected using JalView (Aksamentov et al., 2021). The prevalence of the sequences aligned with primers and crRNAs in different variants were calculated as a percentage of genomes with the complete matched sequence observed in the alignments and represented by the frequency of the sequence among the variant isolates (%F). Consensus sequences of each variant along with their %F values are provided in Figure 3‒ figure supplement 3 for primers and Figure 3‒figure supplement 2 for crRNAs. The associated dataset was available in Figure 3–source data 2.

The NextStrain Clades or PANGO lineages were designated to each WHO classified variant of concern as described in Table S1

### RPA Primer and primer combination screening with colorimetric lateral-flow Cas13a- based readout (for Figure 1‒figure supplement 1)

RT-RPA was set up as previously described (Patchsung et al., 2020) using TwistAmp Basic Kit (TwistDx) and EpiScript reverse transcriptase (Lucigen). Sequences of RPA primers used were given in Table S2. LwaCas13a-based detection reactions were also set up exactly as previously described (Patchsung et al., 2020), then visualized with HybriDetect lateral-flow strips (Milenia Biotec). Sequences of LwaCas13a-crRNAs for each amplicon were given in Table S2.

### Optimized multiplexed RT-RPA

The optimized reaction condition for multiplexed RT-RPA for the amplification of *s* and *n* genes of SARS-CoV-2 with TwistAmp Basic RPA is as follows. The protocol is for the preparation of 5 multiplexed RPA reactions at a time from one lyophilized RPA pellet (TwistAmp Basic kit, TwistDx). Working multiplexed RPA contained 2.5 μM *s*-gene forward RPA primer, 2.5 μM *s*-gene reverse RPA primer, 3.1 μM *n*-gene F4 forward RPA primer, and 3.1 μM *n*-gene R1 forward RPA primer.

One RPA pellet was resuspended with 29.5 μL of the rehydration buffer from the kit. 1 μL EpiScript reverse transcriptase (200 U/μL stock; Lucigen), 0.36 μL RNase H (5 U/μL stock; NEB), 5 μL triglycine (570 mM stock, Sigma), and 5 μL multiplexed RPA primer mix were added to the RPA resuspension. A portion of 8.2 μL of the RPA-primer-enzyme master-mix was aliquoted into five precooled 1.5 ml Eppendorf tubes. 5.3 μL RNA extract from nasopharyngeal and throat swab samples was then added to each aliquot. Lastly, 0.7 μL magnesium acetate (280 mM stock) was added to initiate the amplification. The reactions were incubated at 42 °C for 25 min, then placed on ice before proceeding to the Cas-based detection step.

### Optimized multiplexed Cas13-based detection with fluorescence readout

Each multiplexed Cas13-based detection reaction can be assembled as follows, and making a mastermix by multiplying the amount given is possible: 2 μL HEPES (200 mM stock, pH 6.8), 0.8 μL ribonucleoside triphosphate mix (rNTPs, 25 mM stock, NEB), 0.6 μL NxGen T7 RNA polymerase (50 U/μL stock, Lucigen), 2.5 μL FAM-polyU-IABkFQ reporter (2 μM stock, IDT), 2.5 μL Cy5-polyA-IABkRQ or ROX-polyA-IABkRQ reporter (4 μM stock, IDT), 1 μL LwaCas13a-crRNA for the *s* gene (10 ng/μL stock, Synthego), 1 μL PsmCas13b-crRNA for the *n* gene (30 ng/μL stock, Synthego), 1 μL LwaCas13a enzyme (126 μg/mL; 900 μM working stock in enzyme storage buffer), 1 μL PsmCas13b enzyme (420 μg/mL; 2700 μM working stock in enzyme storage buffer), 3 μL betaine monohydrate (5 M stock, Sigma), 1 μL magnesium chloride (480 mM stock), and 1.3 μL DEPC-treated water.

Final concentrations of each component were: 20 mM HEPES pH 6.8, 1 mM of each rNTP, 1.5 U/μL T7 RNA polymerase, 250 nM polyU-IABkFQ, 500 nM polyA-IABkRQ, 22.5 nM LwaCas13a crRNA, 67.5 nM PsmCas13b crRNA, 45 nM LwaCas13a enzyme, 135 nM PsmCas13b enzyme, 750 mM betaine monohydrate, and 24 mM magnesium chloride.

After mixing, 18 μL of the multiplexed Cas reaction mixture was transferred to a 384-well-plate well, and 2 μL of the multiplexed RPA reaction was added. Fluorescent signals of FAM, ROX, and Cy5 were monitored using a fluorescence microplate reader (Infinite M Plex, Tecan) at 37 °C. Alternatively, FAM and ROX fluorescence can be visualized using an LED transilluminator (BluPAD Dual LED Blue/White light transilluminator) equipped with plastic lighting gels as excitation and emission filters (LEE Filters, filter no.101 Yellow, 106 Primary red, 119 Dark blue, 139 Primary green, 158 Deep orange).

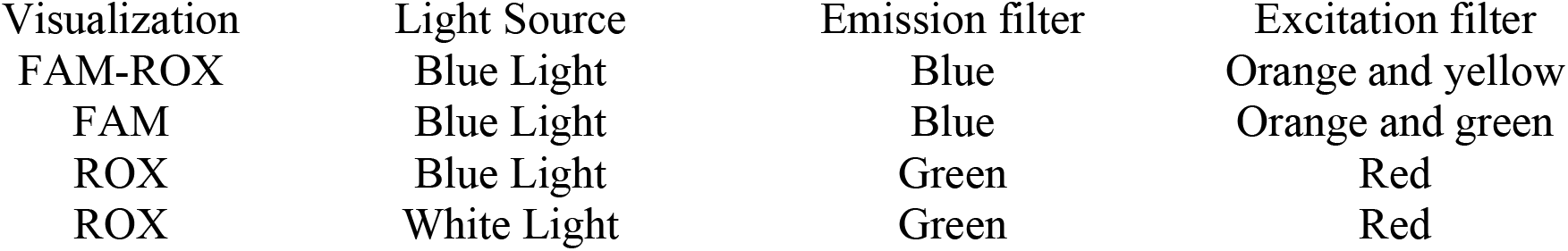

| Visualization | Light Source | Emission filter | Excitation filter |
| --- | --- | --- | --- |
| FAM-ROX | Blue Light | Blue | Orange and yellow |
| FAM | Blue Light | Blue | Orange and green |
| ROX | Blue Light | Green | Red |
| ROX | White Light | Green | Red |

**Specific RPA and Cas-based detection methods** for relevant figures, see Table S3 and Table S4.

### Lyophilized RT-RPA for multiplexed gene detection

An RT-RPA reaction buffer A was prepared from 500 μL 50% (w/v) PEG20000, 250 μL of 1 M Tris pH 7.4, and 100 μL of 100 mM DTT. The multiplexed RT-RPA reaction mixture was prepared by resuspending one RPA pellet (TwistAmp Basic kit, TwistDx) with 14.2 μL of RT-RPA reaction buffer A. 15 μL nuclease-free water, 1 μL EpiScript reverse transcriptase (200 U/μL stock; Lucigen), 0.36 μL RNase H (5 U/μL stock; NEB), 5 μL triglycine (570 mM stock, Sigma), 5 μL of multiplexed RPA primers mix were added to the RPA resuspension. 8.2 μL of the RPA-primer-enzyme master-mix was aliquoted into precooled 1.5 mL Eppendorf tubes; the recipe given was enough to make 5 aliquots. The mixture was snap frozen with liquid nitrogen and lyophilized using a freeze dryer (Alpha 2-4 LDplus, Martin Christ) overnight.

To reconstitute and initiate the RT-RPA reaction, the lyophilized RT-RPA pellet was resuspended with 12.7 μL of the RNA sample. Lastly, 1 µL of a solution composed of 196 mM magnesium acetate and 1.5 M potassium acetate was added to each reaction. The reactions were incubated at 42 °C for 25 min, then placed on ice before proceeding to the Cas-based detection using Cas13a-based detection step containing crRNAs for both S and N genes as previously described. The relative FAM fluorescence intensity was evaluated using a real-time thermal cycler (CFX Connect Real-Time PCR System, Bio-Rad).

### One-pot, multiplexed RT-RPA/CRISPR-Cas13a detection

We prepare the one-pot, monophasic detection reactions by preparing the multiplex RT-RPA and CRISPR-Cas13a reaction mixes separately, before mixing them together, as follows.

RT-RPA reaction buffer and multiplexed RPA primers mix were prepared as described above. crRNA-reporter solution was prepared from 24 μL DEPC-treated water, 4 μL of 100 ng/μL LwaCas13a crRNA for *s* gene, 4 μL of 100 ng/μL LwaCas13a crRNA for *n* gene, and 2 μL of 100 μM FAM-PolyU reporter. Then, CRISPR-Cas13a reaction mix was prepared by combining 2.8 µL rNTPs mix (25 mM each), 2 µL T7 RNA polymerase (50 U/µL), 7 µL LwaCas13a (63 μg/mL), and 3 µL of the crRNA-reporter solution.

The multiplexed RT-RPA reaction mixture was prepared by resuspending one RPA pellet (TwistAmp Basic kit, TwistDx) with 14.2 μL of RT-RPA reaction buffer A. 1 μL EpiScript reverse transcriptase (200 U/μL stock; Lucigen), 0.36 μL RNase H (5 U/μL stock; NEB), 5 μL triglycine (570 mM stock, Sigma), 5 μL of multiplexed RPA primers mix, and 15 μL of CRISPR-Cas13a reaction mix were then added to the RPA resuspension. 8.2 μL of the RPA- primer-enzyme master-mix was aliquoted into precooled 0.1 mL PCR strip tubes; the recipe given was enough to make 5 aliquots.

To initiate amplification and detection, 5.3 μL of the RNA sample was added to each aliquot, followed by 0.7 μL magnesium acetate (280 mM). The reactions were incubated at 39 °C and generated FAM fluorescence was monitored using a real-time thermal cycler (CFX Connect Real-Time PCR System - Bio-Rad).

### Standard (unoptimized) in-house RPA reaction

Standard RPA reactions were set up as previously reported (Piepenburg et al., 2006) and contained the following components: 50 mM Tris (pH 7.5), 100 mM potassium acetate, 14 mM magnesium acetate, 2 mM DTT, 5% (w/v) PEG20000, 200 µM dNTPs, 3 mM ATP, 50 mM phosphocreatine, 100 µg/mL creatine kinase, 120 µg/mL UvsX, 30 µg/mL UvsY, 900 µg/mL Gp32, 30 µg/mL Bsu LF, 450 nM primers, and DNA templates. Reactions were performed in 20 µL volumes at 37 °C for 60 min, followed by inactivation at 65 °C for 10 min. Ten microliters of each reaction were mixed with 2% (w/v) final concentration of sodium dodecyl sulfate (SDS) and Novel Juice DNA staining reagent (BIO-HELIX), separated on 2% (w/v) agarose gel and visualized under blue light (BluPAD, BIO-HELIX).

## Optimizing RPA through titration of protein components

Concentrations of the main protein components of RPA (UvsX, UvsY, Gp32, and Bsu LF) were varied as indicated in . We also switched to using RPA primers for the *n* gene amplification (the F4/R1 pair) and pUC57-2019-nCoV-N plasmid (MolecularCloud cat no. #MC_0101085) at 10,000 copies/µL as the DNA template. The RPA reaction conditions were otherwise identical to the standard RPA reaction (see previous section), and were allowed to proceed at 42°C for 60 min. Thereafter, 2 µL of the RPA products were mixed with 18 µL of the Cas13a-based detection reaction, which contained 20 mM Tris-HCl pH 7.4, 60 mM NaCl, 6 mM MgCl_2_, 1 mM of each rNTPs, 1.5 U/μL NxGen T7 RNA polymerase, 6.3 µg/mL LwaCas13a, 0.5 ng/µL LwaCas13a crRNA, and 0.3 µM FAM-PolyU reporter. The generated FAM fluorescence was monitored at 37 °C over 90 min using a fluorescence microplate reader (Varioskan, Thermo Scientific or Infinite M Plex, Tecan).

### Optimized in-house RPA for single-gene detection

The single-plexed RPA of the *n* gene was set up with the following components (20 µl total reaction volume, consisting of 19 µL reagent mastermix and 1 µL RNA input): 50 mM Tris (pH 7.5), 100 mM potassium acetate, 14 mM magnesium acetate, 2 mM DTT, 5% (w/v) PEG20000, 200 µM dNTPs, 12 mM ATP, 50 mM phosphocreatine, 100 µg/mL creatine kinase, 150 µg/mL (3.3 µM) UvsX, 30 µg/mL (1.7 µM) UvsY, 900 µg/mL (26.5 µM) Gp32, 120 µg/mL (1.8 µM) Bsu LF, 40 mM triglycine (Sigma), and 700 nM *n* gene primers. One µL of pUC57-2019-nCoV-N plasmid (MolecularCloud cat no. #MC_0101085) was used as a template, and the RPA reactions were allowed to proceed at 42°C for 60 min. Thereafter, 2 µL of the RPA products were mixed with 18 µL of the Cas13a-based detection reaction, which contained 40 mM Tris-HCl pH 7.4, 60 mM NaCl, 6 mM MgCl_2_, 1 mM of each rNTPs, 1.5 U/μL NxGen T7 RNA polymerase, 6.3 µg/mL LwaCas13a, 1 ng/µL LwaCas13a crRNA for *n* gene, and 0.3 µM FAM-PolyU reporter. The generated FAM fluorescence was monitored at 37 °C over 90 min using a real-time thermal cycler (CFX Connect Real-Time PCR System, Bio-Rad).

### Optimized in-house RT-RPA for single-gene and multiplexed Cas13-based detection

The single-plexed RT-RPA of the *n* gene was set up with the following components (20 µL total reaction volume, consisting of 14.5 µL reagent mastermix and 5.5 µL RNA input): 50 mM Tris (pH 7.5), 100 mM potassium acetate, 14 mM magnesium acetate, 2 mM DTT, 5% (w/v) PEG20000, 200 µM dNTPs, 12 mM ATP, 50 mM phosphocreatine, 100 µg/mL creatine kinase, 150 µg/mL (3.3 µM) UvsX, 30 µg/mL (1.7 µM) UvsY, 900 µg/mL (26.5 µM) Gp32, 120 µg/mL (1.8 µM) Bsu LF, 26.6 U/mL RNase H, 2.8 U/μL EpiScript Reverse Transcriptase, 40 mM triglycine (Sigma), and 700 nM *n* gene primers. 5.5 µL of serially diluted SARS-CoV- 2 RNA was used as a template, and the RT-RPA reactions were allowed to proceed at 42°C for 60 min.

The multiplexed RT-RPA to detect the *s* and *n* genes of SARS-CoV-2 was set up in a similar condition as the single-plexed amplification, except for the concentration of specific primers, and the amount of RNA template used. Here, we used the *n* gene and *s* gene RPA primers at 222 nM and 177 nM, respectively, and add 7 µL of serially diluted SARS-CoV-2 RNA as input. RT-RPA reactions were performed at 42 °C for 60 min, and the RPA products monitored through multiplexed Cas13a/b-based detection as described in the optimized multiplexed Cas13-based detection methods section.

Thereafter, 2 µL of the RPA products were mixed with 18 µL of the Cas13a-based detection reaction, which contained 40 mM Tris-HCl pH 7.4, 60 mM NaCl, 6 mM MgCl_2_, 1 mM of each rNTPs, 1.5 U/μL NxGen T7 RNA polymerase, 6.3 µg/mL LwaCas13a, 1 ng/µL LwaCas13a crRNA (1 ng/µL LwaCas13a crRNA for single gene detection and 0.5 ng/µL LwaCas13a crRNA for *s* gene, 0.5 ng/µL LwaCas13a crRNA for *n* gene for multiplexed detection), and 0.3 µM FAM-PolyU reporter. The generated FAM fluorescence was monitored at 37 °C over 90 min using a real-time thermal cycler (CFX Connect Real-Time PCR System, Bio-Rad).

### Lyophilized in-house RT-RPA for multiplexed gene detection

The multiplexed RT-RPA reaction mixture was prepared by assembling all components as described in the optimized in-house RT-RPA methods section, except that magnesium acetate and potassium acetate were not included, and the mixture was further supplemented with 6% (w/v) trehalose. The mixture was lyophilized using a freeze dryer (Alpha 2-4 LDplus, Martin Christ).

To reconstitute and initiate the RT-RPA reaction, the lyophilized RT-RPA pellet was resuspended in a solution consisting of 1 μL of 280 mM magnesium acetate, 0.4 μL of 5 M potassium acetate, and 17.6 μL of SARS-CoV-2 RNA. The RT-RPA reaction was incubated at 42 °C for 60 min. A portion of the reaction was then transferred to the Cas13a-based detection step containing crRNAs for both *s* and *n* genes as previously described. The generated FAM fluorescence was monitored using a real-time thermal cycler (CFX Connect Real-Time PCR System, Bio-Rad).

### Sample collection, RNA preparation, and ethical approval

As previously described (Chaimayo et al., 2020), respiratory samples were collected from patients with suspected SARS-CoV-2 infection at Siriraj Hospital and processed at the Diagnostic Molecular Laboratory, Department of Microbiology, Faculty of Medicine, Siriraj Hospital. Nasopharyngeal and throat swabs were collected in viral transport media (VTM, Gibco). RNA was extracted from 200 µL of the respiratory swabs in VTM using magLEAD® 12gC automated extraction platform (Precision System Science, Japan), eluted in 100 µL buffer, and either used directly for clinical laboratory diagnosis of SARS-CoV-2 via RT-qPCR or stored at -70 °C for later use.

Excess RNA extracts from these samples were then used without any personally identifiable information being collected for our studies. 91 samples with known positive results from RT-PCR for SARS-CoV-2, 45 samples with known negative results from RT-PCR for SARS-CoV- 2, 20 Delta clinical samples, 10 Alpha clinical samples, 15 Omicron clinical samples, and 3 samples with positive results for human coronavirus OC43, NL63, and 229E were included for the method validation. Samples were randomized upon giving to study staff, who were kept blinded to the SARS-CoV-2 RT-PCR diagnostics results of the samples when they performed validation experiments of CRISPR diagnostics.

Ethical approval of the study was given by the Siriraj Institutional Review Board (COA: Si 339/2020 and Si 424/2020).

### Cultured SARS-CoV-2 viral RNA extracts and serial dilutions

SARS-CoV-2 Wuhan variant (clinical isolate hCoV-19/Thailand/Siriraj_5/2020; GISAID accession ID: EPI_ISL_447908), Alpha variant (clinical isolate hCoV- 19/Thailand/Bangkok_R098/2021; GISAID accession ID: EPI_ISL_11000098), and Delta variant (clinical isolate hCoV-19/Thailand/Bangkok_SEQ4389/2021; GISAID accession ID: EPI_ISL_3038904) were propagated in Vero E6 cells cultured in MEM-E supplemented with 10% fetal bovine serum. The culture media were collected and extracted as described for the clinical sample. The RNA extracts at given dilution were prepared by dilution with nuclease-free water and stored at -70 °C for later use.

### RT-qPCR

The RT-qPCR was performed according to a published protocol (Vogels et al., 2020). In brief, a 5 µL of the RNA sample was added to a 20-µL reaction prepared with Luna® Universal Probe One-Step RT-qPCR Kit (New England Biolabs) and nCoV_N1 primer-probe set (Integrated DNA Technologies). The reactions were monitored using CFX Connect Real-Time PCR System (Bio-Rad).

Allplex™ SARS-CoV-2 Variants I Assay (Seegene, Korea), which targets *RdRp* and *S*; delH69/V70, E484K and N501Y of SARS-CoV-2, was used for SARS-CoV-2 Alpha and Omicron variant screening. Allplex™ SARS-CoV-2 Variants II Assay (Seegene, Korea), which targets *RdRp* and *S*; K417N, K417T, L452R and W152C, was used for Delta variant screening. All identified variant samples were confirmed with full length spike sequencing.

### Droplet digital RT-PCR (RT-ddPCR)

The RT-ddPCR was performed using One-Step RT-ddPCR Advanced Kit for Probes (Bio-Rad) according to the manufacturer’s protocol and in line with RT-qPCR protocol described above. A 20-µL reaction consisted of 5 µL of RNA input, 5 µL of Supermix, 2 µL of reverse transcriptase, 1 µL of 300 mM DTT, 4 µL nuclease-free water, and each 1 µL of following component from nCoV_N1 primer-probe set (Integrated DNA Technologies): 10 µM forward primer, 10 µM reverse primer, and 10 µM probe. The reaction droplets were generated and read using a QX100™ Droplet Digital™ PCR system (Bio-Rad) connected to a T100™ Thermal Cycler (Bio-Rad). Thermocycling conditions were reverse transcription at 50 °C for 60 minutes, enzyme activation at 95 °C for 10 minutes, followed by 40 cycles of 95 °C for 15 seconds, 55 °C for 30 seconds, 58 °C for 10 seconds. The enzyme deactivation was done at 98°C for 10 minutes. The quantification was carried out using QuantaSoft™ software (Bio-rad).

**Standard curve of qPCR Ct versus ddPCR-derived copies/uL RNA input** see Appendix 1

### Data analysis and visualization

Statistical analyses were performed, and graphical representations created with GraphPad Prism 9, unless indicated otherwise. Schematic diagrams were created with Biorender.com and Adobe Illustrator CC 2017.

## Data Availability

The main data supporting the results in this study are available within the paper and its supplementary information. Raw datasets generated and analysed during the study are available from the corresponding authors upon reasonable request.

## Acknowledgments

We thank Prof Alice Ting (Stanford) for Cas13d and ultraTEVp plasmids, Dr Panvika Pannopard, Nujaree Promjin, and Veeraya Sachue (VISTEC) for management and logistical support, Prof Wipa Suginta and Prof Daniel Crespy (VISTEC) for equipment use and advice, Dr Charlie Gilbert (Cambridge) for plasmids and helpful advice, and Kamnoetvidya Science Academy for providing accommodation and equipment. Several equipment used for the project was generously provided on loan by Bang Trading, Eppendorf Thailand, PCL Holding, PTT Innovation Institute, and Theera Trading. The project is financially supported by PTT, PTTEP, Kasikorn Bank, Siam Commercial Bank, GPSC, VISTEC, Siriraj Hospital, Mahidol University, and Department of Medical Sciences, Ministry of Public Health. C.C. was funded by Wellcome International Intermediate Fellowship (216457/Z/19/Z) and Sanger International Fellowship. M.A.C and P.S.F. acknowledge funding from UKRI-EPSRC (EP/R014000/1, EP/S001859/1), UKRI-BBSRC (BB/M025632/1) and the UK Dementia Research Institute.

## Author contributions

Conceptualization: MP, AH, SS, KA, CU, DP, NH

Methodology: MP, AH, SS, KA, CU, DP, NH, SC, CC

Investigation: MP, AH, SS, KA, AP, KS, PM, TWo, AA, KJ, NAt, SV, TWa

Material support: NN, PC, RT, NW, SM, MAC, PSF, JJ, AL, OA, JG, FZ, SC

Clinical study support: AP, NAt, JB, RS, NAn, NH

Supervision: CU, DP, NH

Writing—original draft: MP, AH, SS, KA, CU, DP

Writing—reviewing and editing: all authors

## Competing interests

O.A., J.G., and F.Z. are co-founders of Sherlock Biosciences and Pine Trees Health. M.A.C. and P.S.F. have acted as consultants for Analytik Jena GmbH. M.P., A.H., K.A., S.S., D.P., and C.U. have filed a patent in Thailand on the formulations for multiplexed detection of SARS-CoV-2 RNA. The remaining authors declare no competing interests.

---

**Figure 1-figure supplement 1:**
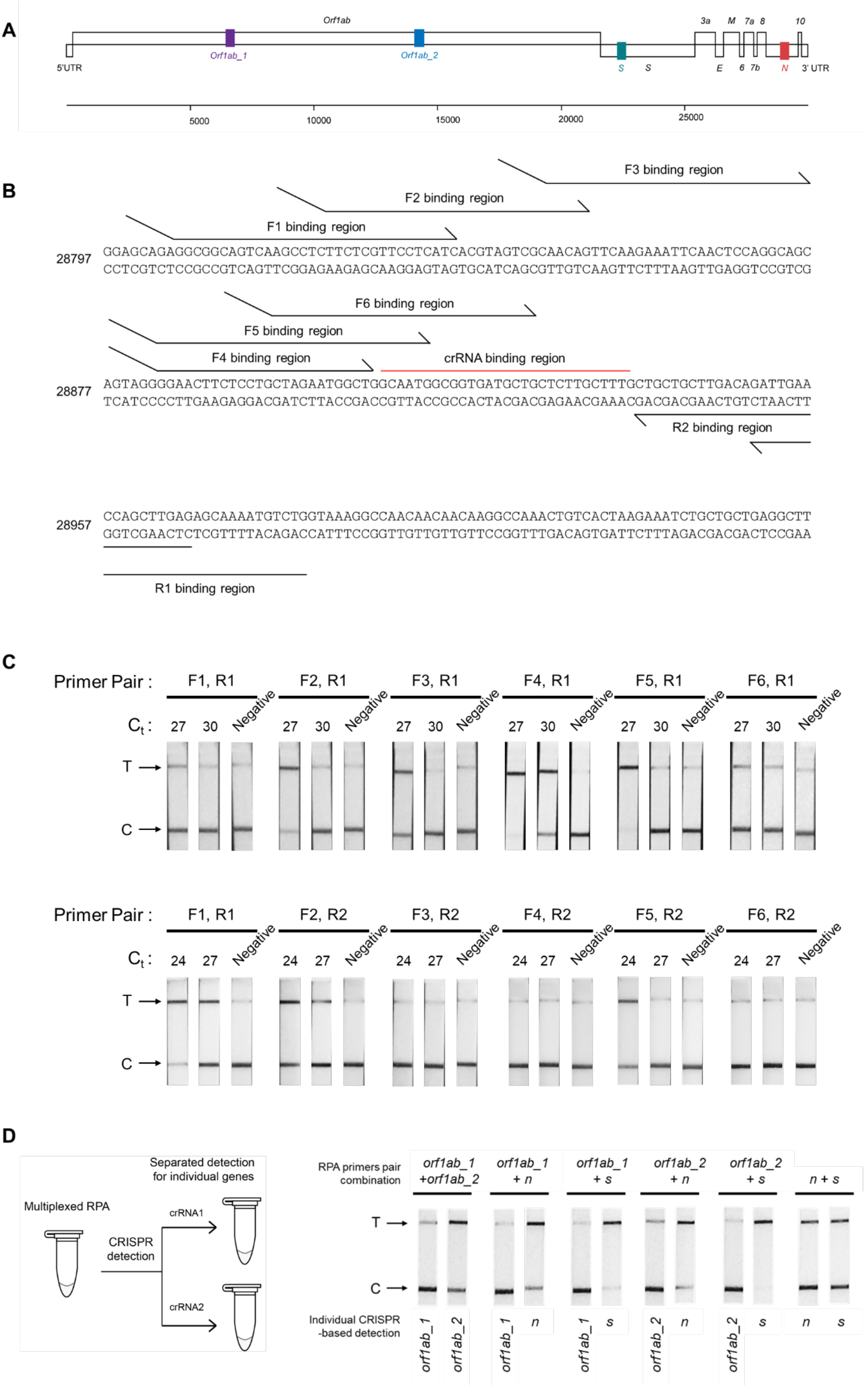
Optimizing the RT-RPA amplification of SARS-CoV-2 *n* gene. (**A**) Four regions within the *orf1ab*, *s*, and *n* genes of SARS-CoV-2 genome selected for detection are highlighted. (**B**) Binding positions of different primers to the SARS-CoV-2 *n* gene. (**C**) Testing efficiency of different primer pairs on SARS-CoV-2 RNA. RT-RPA using each primer pair was performed with SARS-CoV-2 RNA at two different concentrations as a template. Cas13a-based detection with lateral-flow readout was then used to assess successful amplification. Experiments with R1 reverse primers (Qian et al.) and R2 reverse primers (bottom) were performed using different SARS-CoV-2 RNA dilutions, but with an identical control primer pair (F1, R1) included in both sets of experiments, allowing us to compare relative performance of all primer pairs. Based on this screening, we selected the (F4, R1) primer pair for subsequent *n* gene amplifications. **D**) Multiplexed RPA reactions with pairwise combinations of RPA primers (4 different primer pairs; 6 pairwise combinations). A multiplexed RPA is individually assessed with a LwaCas13a-based detection reaction, each programmed with a specific crRNA for the SARS-CoV-2 gene, and lateral-flow readout. Successful amplification was marked by production of a strong-colored band at a test band (T), which indicates target-activated Cas13a activity and resulting in efficient cleavage of the FAM-biotin reporter.

**Figure 1-figure supplement 2:**
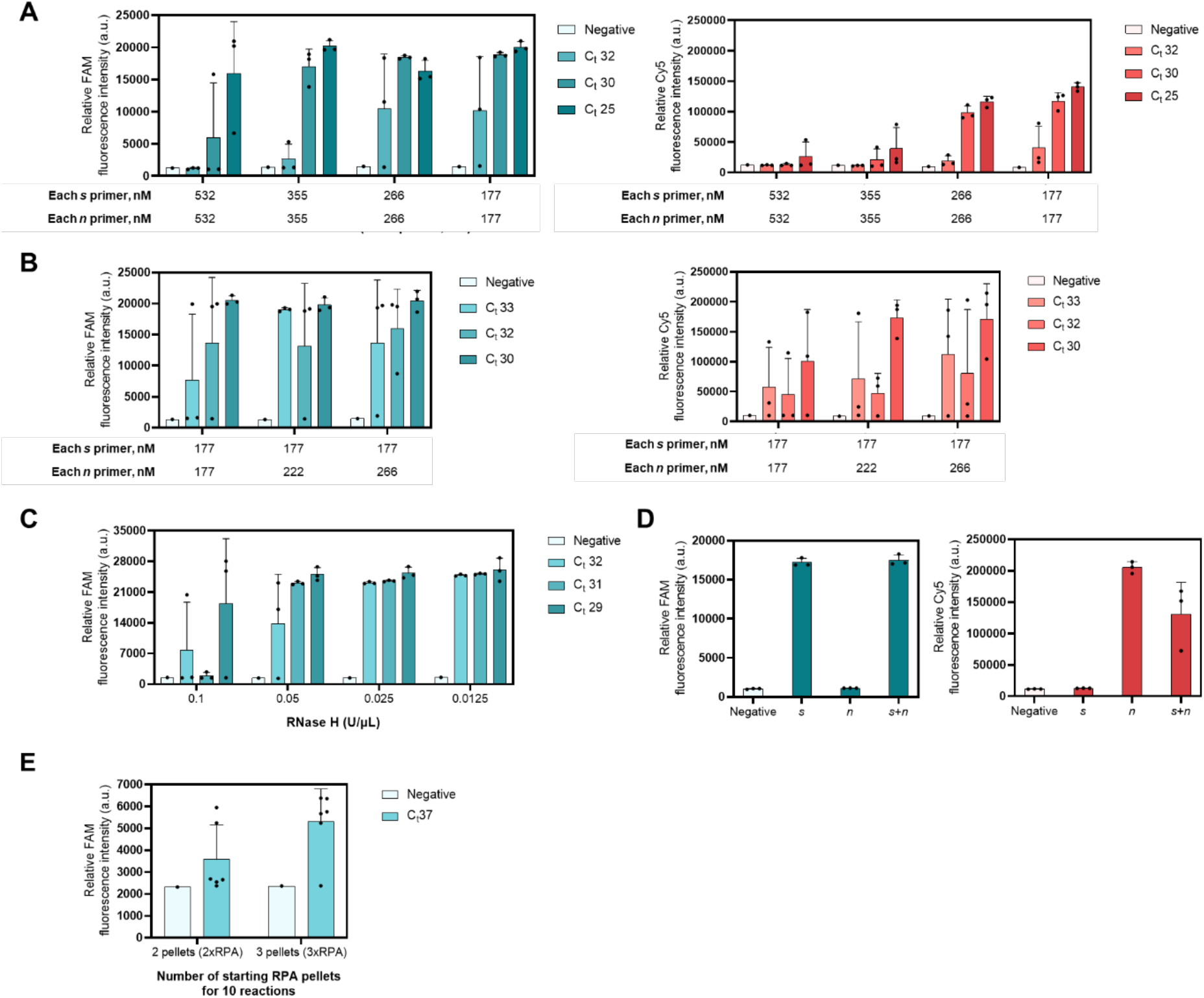
Evaluation of the sensitivity and specificity of the Multiplexed RT-RPA assay with real-time fluorescence detection. (**A**) Optimizing total primer concentrations in multiplexed RPA with 1:1 malar ratio of *n* to *s* primer. Multiplexed RPA for the *s* and *n* gene of SARS-CoV-2 was performed using indicated primer concentrations and serially diluted SARS-CoV-2 genomic RNA template. In the second step, generated *s* and *n* amplicons were detected using a multiplexed CRISPR-Cas reaction containing LwaCas13a and PsmCas13b enzymes. LwaCas13a is programmed with a crRNA targeting the *s* amplicon and cleaves a FAM/IABkFQ -functionalized polyU reporter once target-activated (left), while PsmCas13b is programmed with a crRNA targeting the *n* amplicon and cleaves a Cy5/IABkRQ-functionalized polyA reporter (right). (**B**) Fine-tuning primer concentrations in the multiplexed RPA reaction. Multiplexed RPA for the *s* and *n* gene of SARS-CoV-2 was performed using indicated primer concentrations, with serially diluted SARS-CoV- 2 genomic RNA as a template. Amplicons were detected in a multiplexed CRISPR-Cas reaction, via FAM fluorescence generated from *s*-targeted LwaCas13a (left) and Cy5 fluorescence from N-targeted PsmCas13b (right). (**C**) Optimizing RNase H concentrations in the RT-RPA reaction. RT-RPA for *s* gene amplification from serially diluted SARS-CoV-2 RNA, followed by LwaCas13a-based detection, was used. (**D**) Orthogonality of LwaCas13a and PsmCas13b in a multiplexed CRISPR-Cas detection. RPA reactions were performed with only *s* primers (*s*), only *n* primers (*n*), or combined *s* and *n* primers (*s*+*n*). Endpoint fluorescence intensities of the multiplexed detection reactions (90 min) containing all components for LwaCas13-based detection of the *s* gene and PsmCas13b-based detection of the *n* gene were shown. Generated FAM (left) and Cy5 (right) fluorescence indicated cleavage of the reporters by the *s*-targeted LwaCas13a and the *n*-targeted PsmCas13b, respectively. For C-F, negative controls have no SARS-CoV-2 RNA template input. Data are mean ± s.d. from 3 replicates. (**E**) Analytical sensitivity of increasing of RPA pellet. Multiplexed RT-RPA for the *s* and *n* gene of SARS-CoV-2 was performed using indicated number of RPA pellets, with diluted SARS-CoV-2 genomic RNA at C_t_ 37 as a template. Subsequently, the amplicons were detected via FAM fluorescence generated from LwaCas13a-based reaction programmed with *s*- and *n*-targeted crRNAs.

**Figure 1-figure supplement 3:**
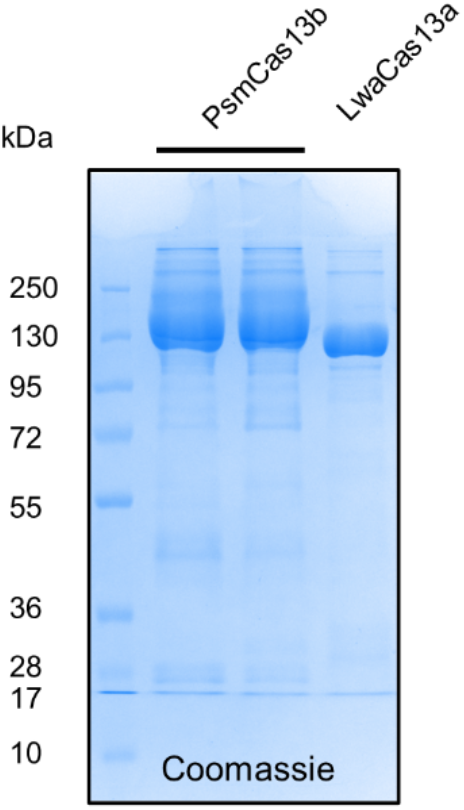
Protein purification of LwaCas13a and PsmCas13b. 12% SDS-PAGE gel of purified PsmCas13b and LwaCas13a. Each lane was loaded with 6 μg of protein. PsmCas13b was loaded twice.

**Figure 1-figure supplement 4:**
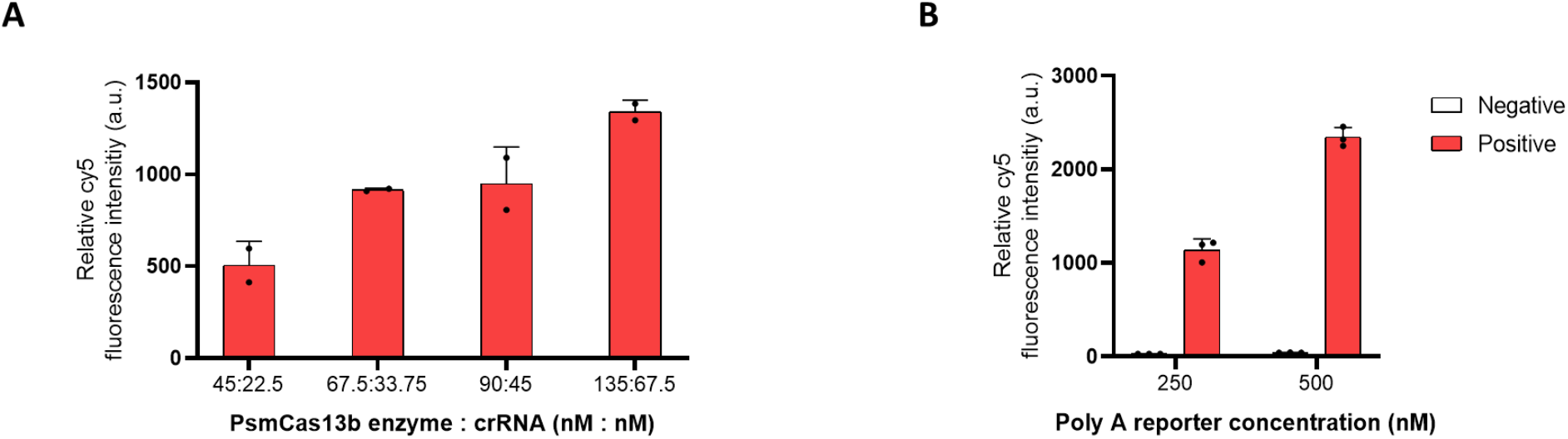
Optimizing PsmCas13b-based detection of the SARS-CoV-2 N gene. (**A**) Varying PsmCas13b enzyme and crRNA amount in the detection reaction. The concentration of Cy5-functionalized polyA reporter was 250 nM in all conditions in (A). (**B**) Varying the amount of Cy5-polyA reporter in the detection reaction. 135 nM PsmCas13b and 67.5 nM crRNA were used for all conditions in (B). The *n* amplicon from the same RPA reaction was used as a substrate for all PsmCas13b-based detection reactions, which were performed for 90 min at 37 °C, without any special additive in the RPA nor the CRISPR-Cas reactions. Endpoint fluorescence intensities were shown. Negative control reactions in (B) have no SARS-CoV-2 RNA template input. Error bars, ± s.d. from 2 replicates (A) and 3 replicates (B).

**Figure 1-figure supplement 5:**
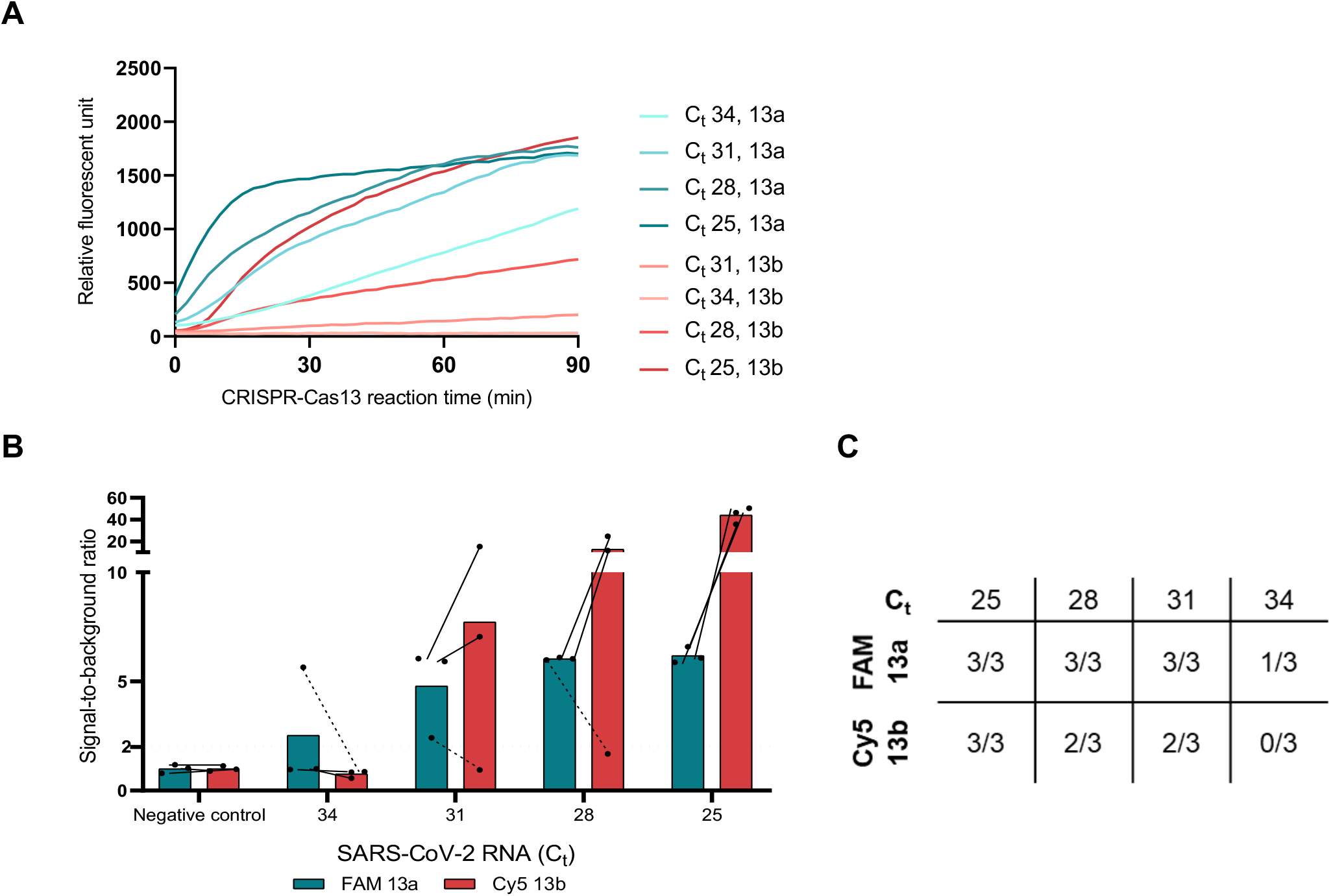
Comparison of LwaCas13a- and PsmCas13b-based detection. The *n* amplicons from the RPA reactions performed with serially diluted SARS-CoV-2 RNA were used as substrates for all Cas13-based detection reactions, which were performed for 90 min at 37°C. The concentrations of all other components in the CRISPR-Cas reactions were identical, and no special additive was added to the RPA nor the CRISPR-Cas reactions. (**A**) Kinetics of fluorescence signal generation from PsmCas13b-mediated detection is slower than that of LwaCas13a. (**B**) When we calculated signal-to-noise (S/N, with noise defined as fluorescence intensity obtained from a no input sample performed in parallel), PsmCas13b generates higher signal-to-noise than LwaCas13a, primarily due to lower background fluorescence in the negative control sample for PsmCas13b. However, LwaCas13a-mediated detection still has higher positive rates **(B, C)** than PsmCas13b. Error bars, ± s.d. from 3 replicates.

**Figure 1-figure supplement 6:**
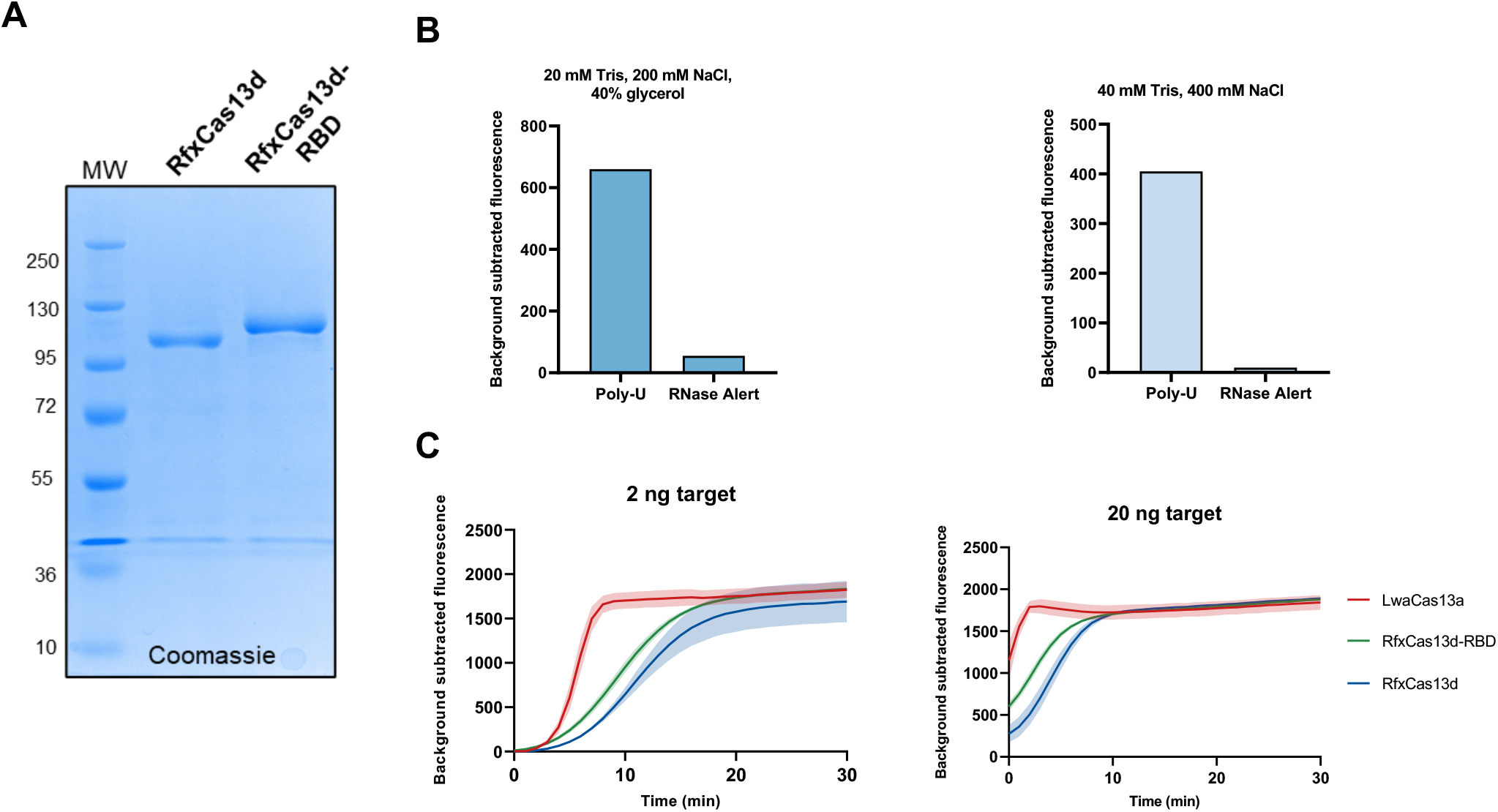
RfxCas13d-based detection of SARS-CoV-2 RNA. (**A**) SDS-PAGE gel of purified RfxCas13d (112 kDa) and RfxCas13d-RBD (120 kDa). Each lane was loaded with 2 ng of protein. (**B**) RfxCas13d exhibited collateral cleavage preference in degrading a polyU reporter over RNaseAlert®. An *n* gene amplicon of SARS-CoV-2 was used as a substrate in a Cas13d-mediated detection. Two reaction conditions as well as two potential collateral reporters of RfxCas13d were tested. Fluorescence signal generated after 60 min of the CRISPR-Cas reaction for each condition (performed once) is shown. (**C**) Comparison of LwaCas13a, RfxCas13d, and RfxCas13d-RBD in SHERLOCK detection of the SARS-CoV-2 *n* gene. RfxCas13d-RBD has improved collateral cleavage efficiency over RfxCas13d, but still does not match LwaCas13a. Fluorescence signal generated over time using the FAM-polyU reporter is shown, at 2 ng (left) and 20 ng (right) RNA input amount. Error bands in (C) ± s.d. from 3 replicates.

**Figure 1-figure supplement 7:**
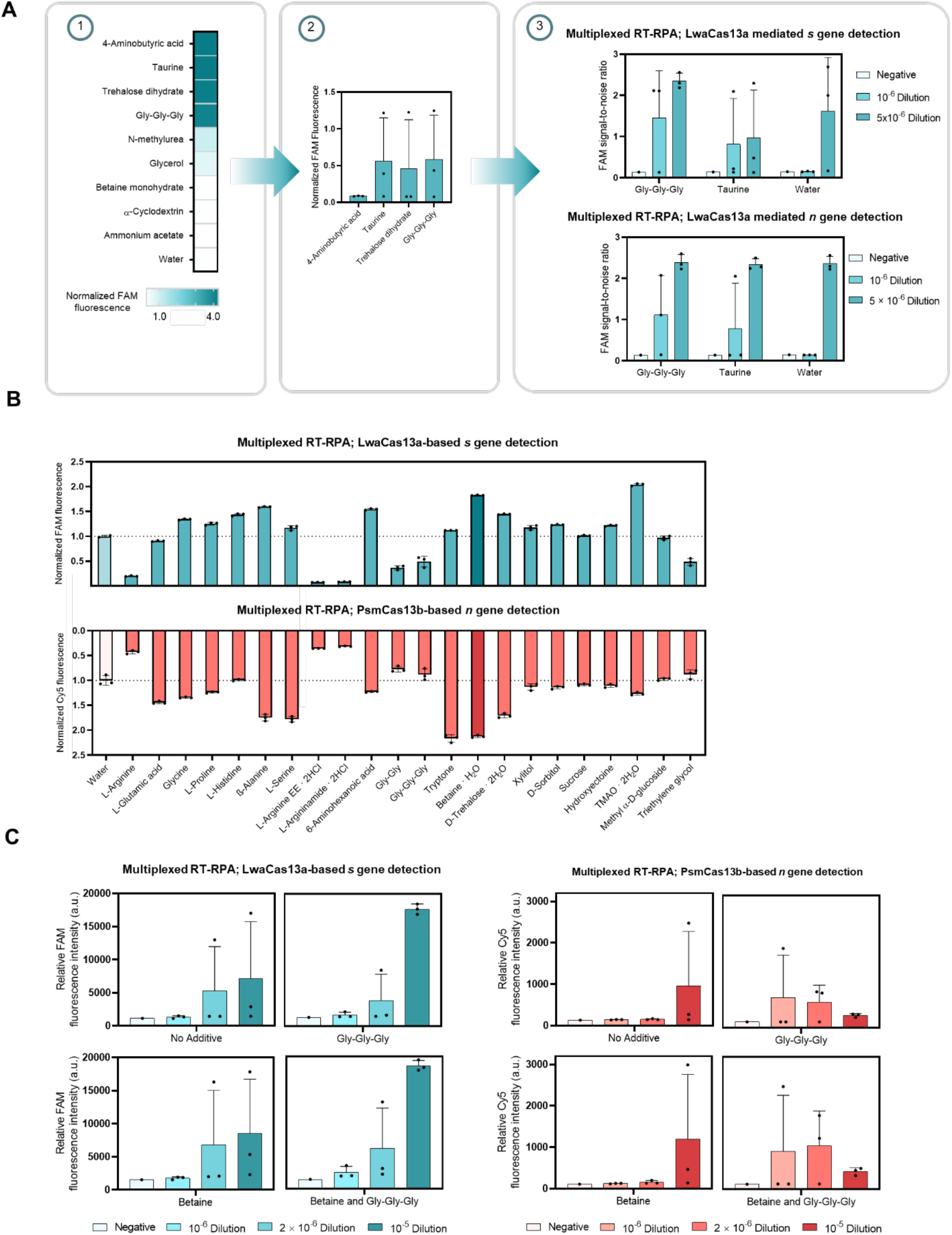
Screening of additives to enhance multiplexed RPA and CRISPR-Cas reactions. (**A**) Screening for additives that enhance multiplexed RT-RPA. Left: nine additives were assessed in a conventional two-step (RPA, then LwaCas13a-based CRISPR-Cas) SHERLOCK detection for the *s* gene of SARS-CoV-2. Middle: four active additives were evaluated further in triplicates, using less RNA input. Right: two best-performing additives—triglycine and taurine—were finally assessed in a multiplexed RPA reaction for the *s* and *n* gene of SARS-CoV-2. Two serial dilutions of SARS-CoV-2 RNA were used as a template; negative control has no SARS-CoV-2 RNA template input. Generated *s* and *n* amplicons were detected in separate LwaCas13a-based detection reactions, whose FAM fluorescence signal is shown. (**B**) Screening for additives that improve the multiplexed CRISPR reaction. Twenty-two additives were assessed in the multiplexed CRISPR-Cas detection, using an RPA product from a multiplexed. Endpoint FAM (Qian et al.) and Cy5 (bottom) fluorescence intensities were normalized against intensities obtained from the no-additive controls. Data are mean ± s.d. from 3 replicates. L-Arginine EE. 2HCl, L-Arginine ethyl ester dihydrochloride. TMAO, trimethylamine N-oxide. (**C**) Cumulative benefits of triglycine additive in the multiplexed RPA and betaine monohydrate additive in the multiplexed CRISPR-Cas reaction. Multiplexed RPA to amplify the *s* and *n* genes of SARS-CoV-2 was performed in the presence or absence of 40 mM triglycine, using serially diluted SARS-CoV-2 RNA as a template, and multiplexed RPA was allowed to proceed for 25 min at 42 °C. Multiplexed CRISPR-Cas reactions to detect the *s* and *n* amplicons were then performed in the presence or absence of 500 mM betaine. FAM and Cy5 fluorescence signal (indicative of Cas13a-mediated *s* gene and Cas13b-mediated *n* gene detection respectively) is shown. RNase-free water was used as input of all negative control reactions. Data are mean ± s.d. from 3 replicates.

**Figure 1-figure supplement 8:**
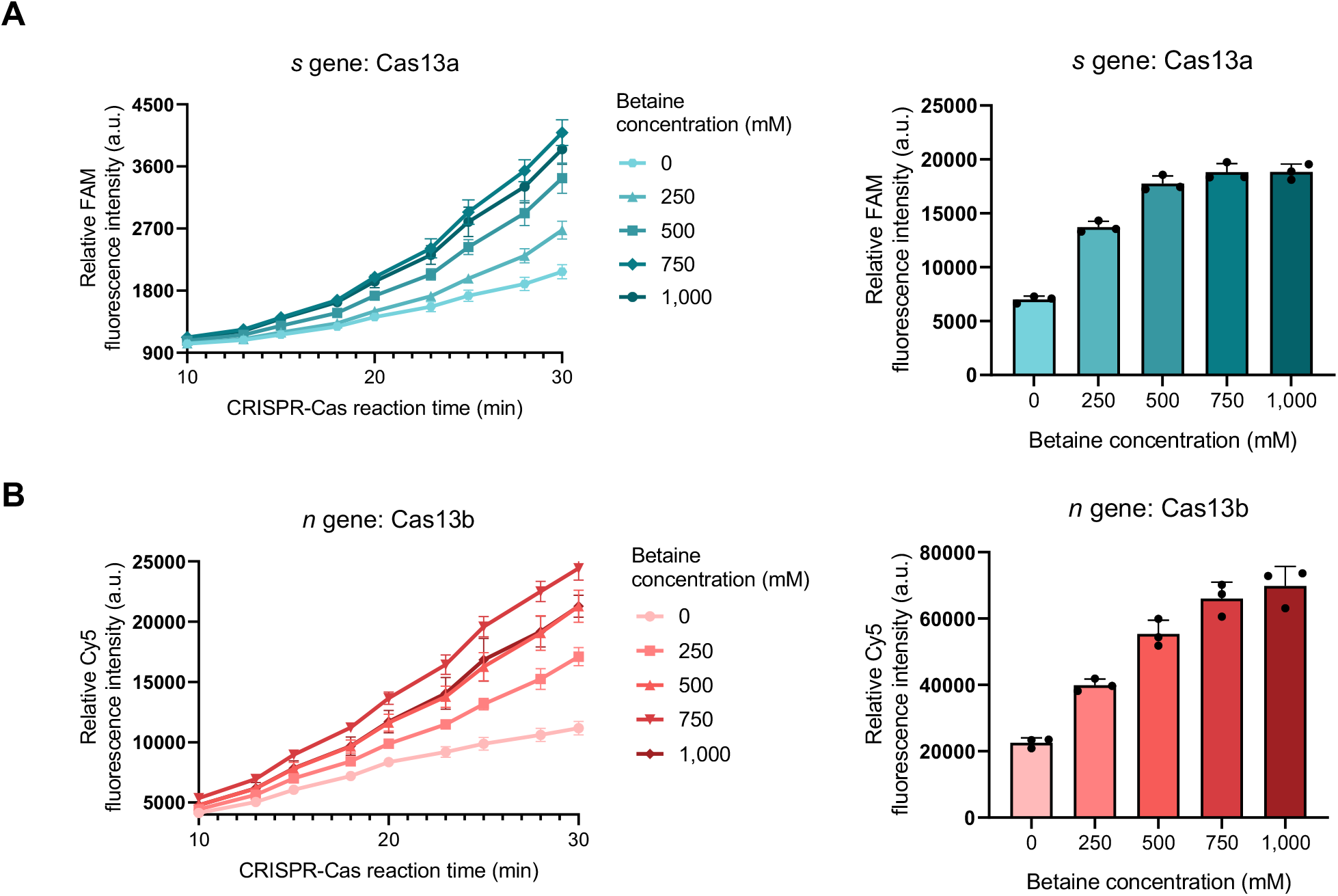
Optimizing betaine monohydrate concentration in multiplexed CRISPR-Cas detection. FAM fluorescence indicated cleavage of the FAM reporter by the *s*-targeted LwaCas13a, while Cy5 fluorescence was from cleavage of the Cy5 reporter by the *n*-targeted PsmCas13b. FAM (**A**) and Cy5 (**B**) fluorescence signal generated after 30 min of the multiplexed CRISPR-Cas reaction for each condition are shown. The target was a diluted RPA product at 1:300 dilution. Error bars, ± s.d. from 3 replicates.

**Figure 1-figure supplement 9:**
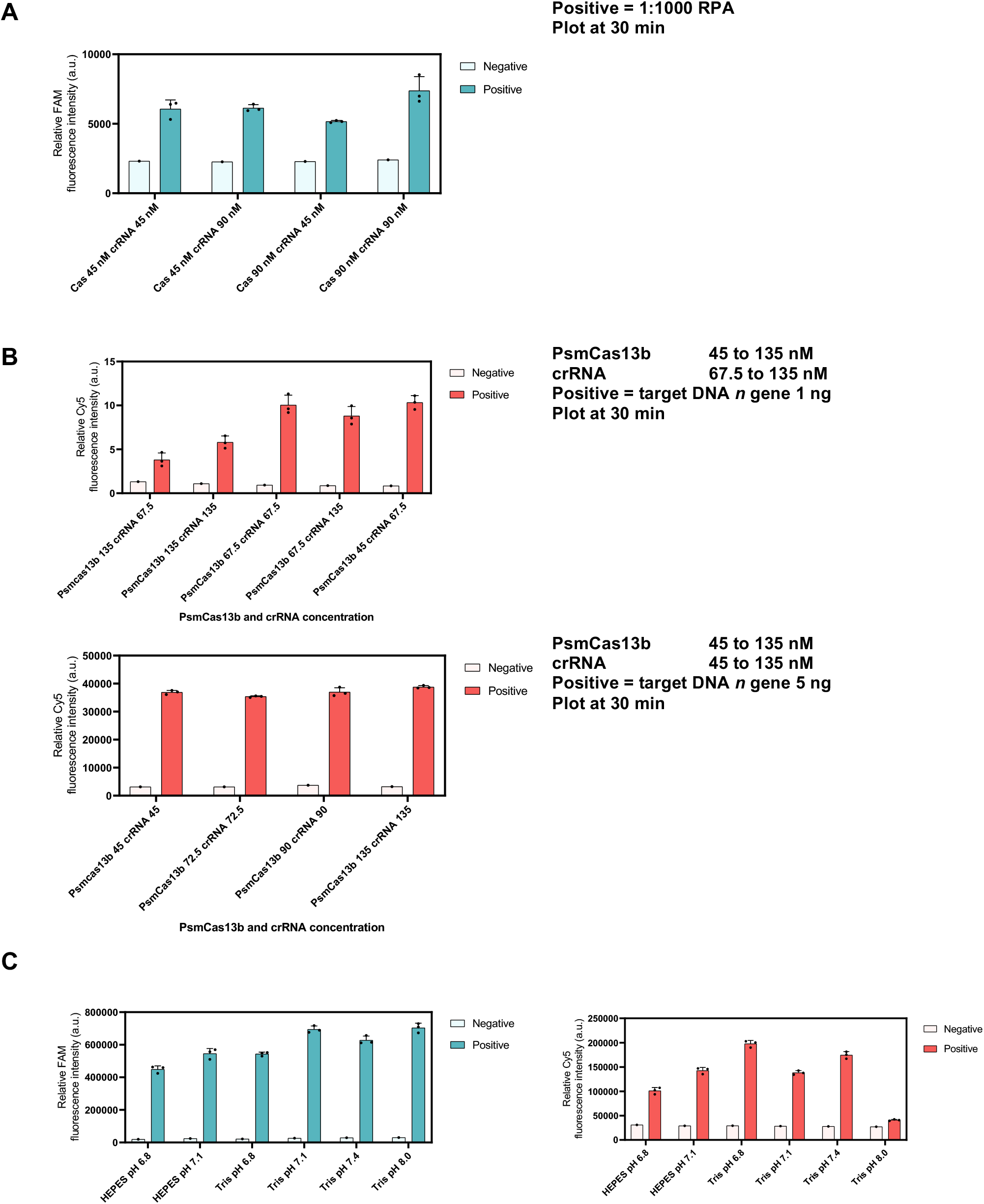
Effect of crRNA ratio and pH in multiplexed CRISPR-Cas 13 based detection. (A) LwaCas13a-based detection of a diluted RPA product at different concentration of LwaCas13a (45 – 90 nM) and LwaCas13a-crRNA (45 – 90 nM) for the *s* gene. (B) PsmCas13b-based detection of purified N amplicon at different concentration of PsmCas13b (45 – 135 nM) and PsmCas13b-crRNA (45 – 135 nM) for the *n* gene. (C) Identifying optimal buffer conditions for LwaCas13a and PsmCas13b. The multiplexed CRISPR-based detection reaction prepared in Tris-HCl or HEPES buffer (pH 6.8 – 8.0) were performed using a diluted multiplexed RPA product as input. Error bars, ± s.d. from 3 replicates.

**Figure 1-figure supplement 10:**
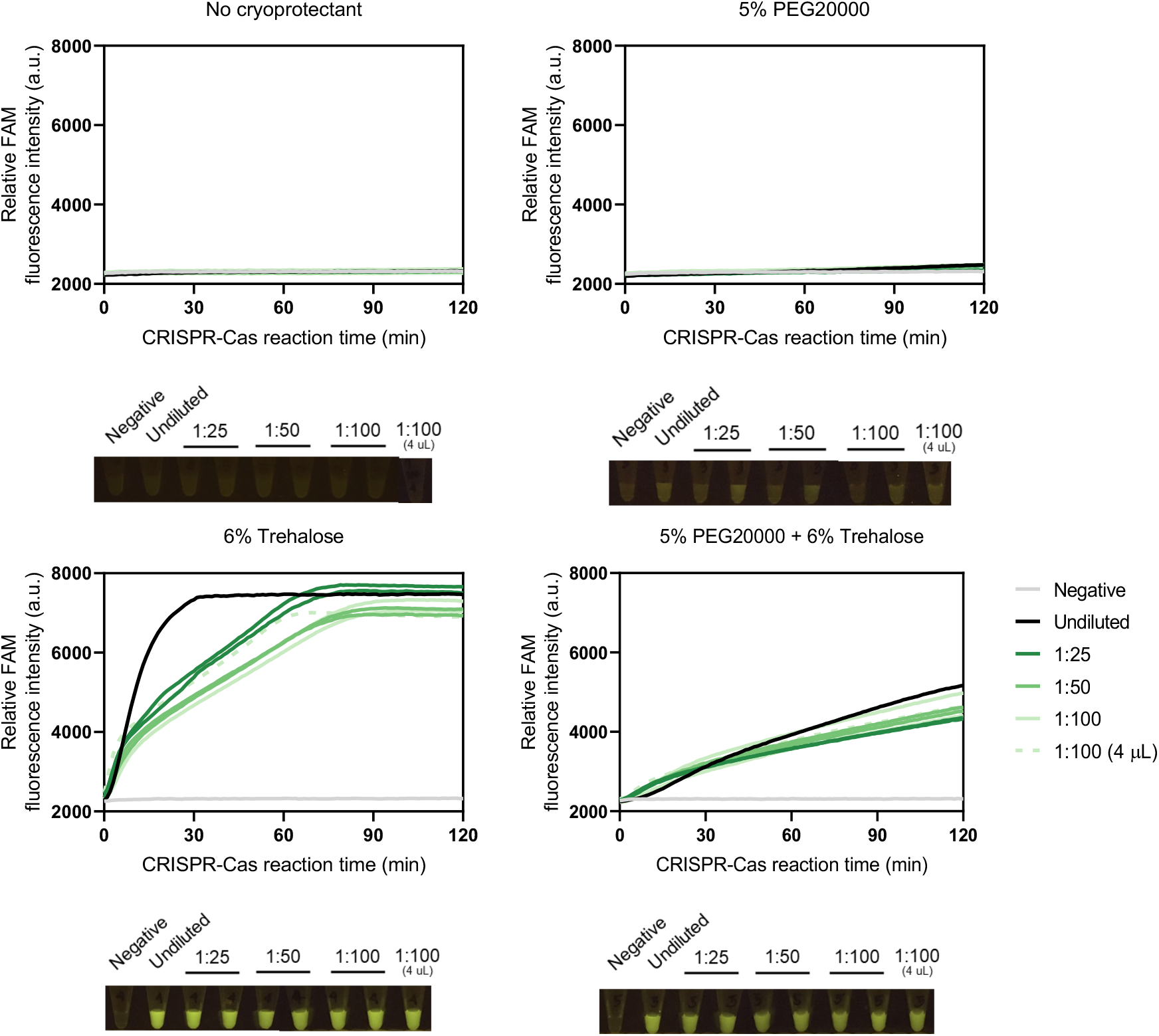
Effects of cryoprotectants on lyophilized CRISPR-Cas13a reactions. LwaCas13a-based detection reactions were prepared with or without 5% (w/v) PEG20000 and 6% (w/v) trehalose added as a cryoprotectant. MgCl_2_ was omitted from all reactions and was added at the rehydration step. The detection was performed at 37 °C using *n* gene RPA product at various dilution levels and monitored under FAM channel using a real-time thermal cycler. The input volume of the RPA dilutions was 2 μL except for the 1:100 dilution sample in which we used either 2-μL or 4-μL input volume. The end-point fluorescence in tubes was visualized using a BluPAD transilluminator.

**Figure 1-figure supplement 11:**
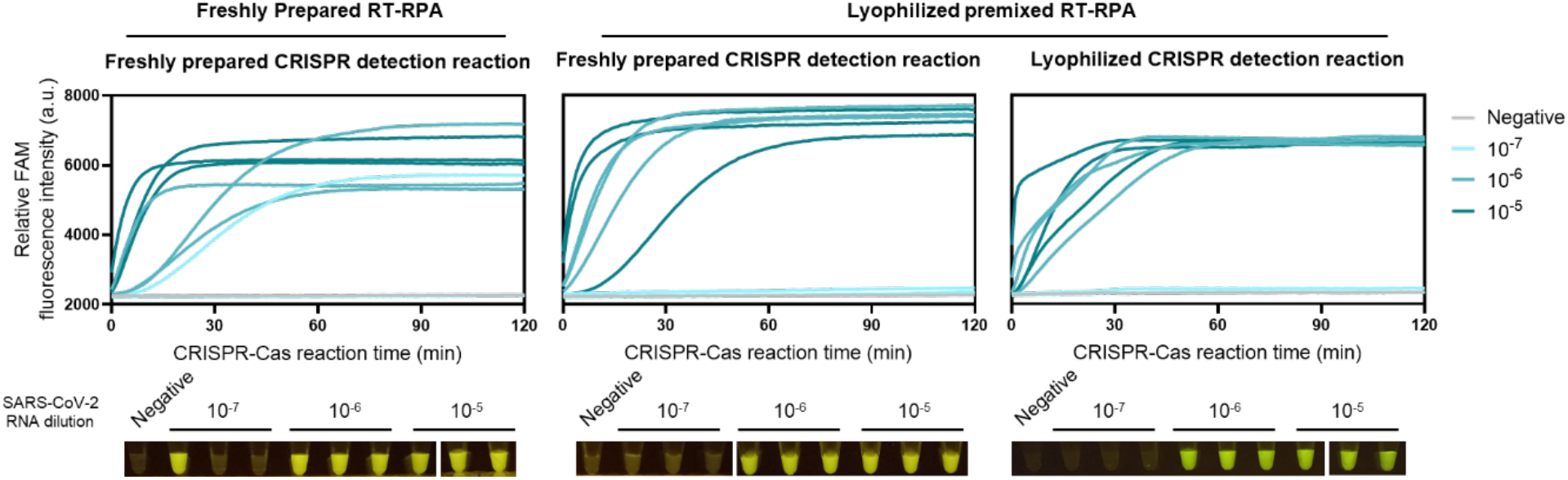
lyophilized RT-RPA for multiplexed amplification of the S and N genes. Left vs middle graphs: all-in-one lyophilized RT-RPA for multiplexed amplification of the *s* and *n* genes has similar sensitivity as freshly prepared RT-RPA. Non-volatile salt KOAc (the high concentration of which likely destabilizes protein components of RT-RPA upon lyophilization) and the typical reaction initiator Mg(OAc)_2_ were excluded, while all other components required for multiplexed RT-RPA including all protein components for RPA, reverse transcriptase, RNase H, primer pairs, and other substrates and buffering/crowding compounds can be lyophilized together. The lyophilized RT-RPA pellets were reconstituted with 12.7 µl of serially diluted SARS-CoV-2 RNA, Mg(OAc)_2_, and KOAc, and incubated at 42°C for 25 min. Thereafter, the multiplexed RPA products were used in a freshly prepared Cas13a-based detection with *s*- and *n*-targeted crRNAs. FAM fluorescence generated over 120 min for each condition is shown. Middle vs right graphs: lyophilized LwaCas13a-based detection reaction has similar sensitivity as freshly prepared reactions.

**Figure 2-figure supplement 1:**
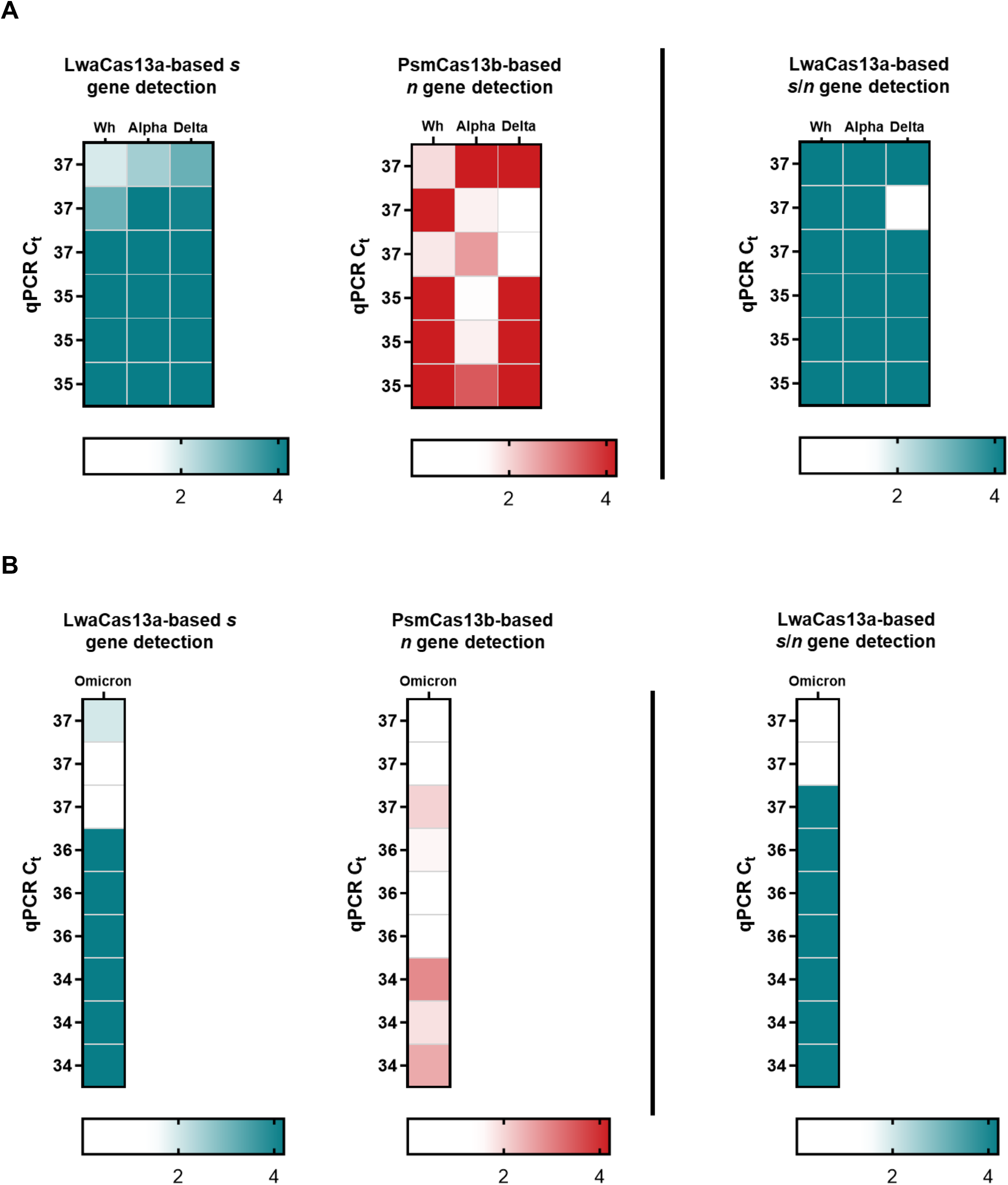
The lyophilized multiplexed CRISPR-based detection is robust across major SARS-CoV-2 variants. Performance of the multiplexed CRISPR-based detection for **(A)** ancestral Wuhan strain, Alpha variant, Delta variant, and **(B)** Omicron variant. FAM and Cy5 fluorescence intensities at 60 minutes were normalized against averaged intensities obtained from the no template control.

**Figure 3-figure supplement 1:**
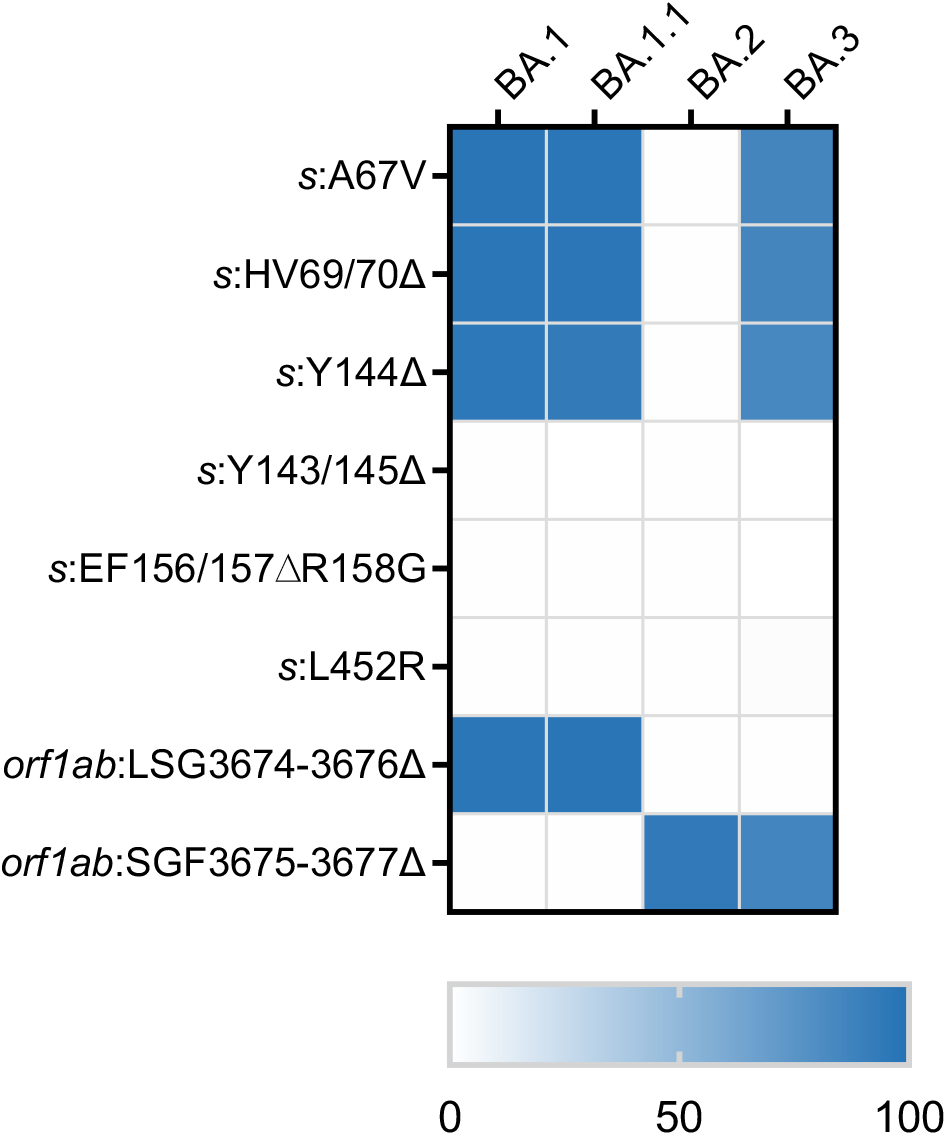
Mutation prevalence across Omicron sub-lineages. Percentages of mutations prevalence across four SARS-CoV-2 Omicron sub-lineages (BA.1, BA.1.1, BA.2, and BA.3) were obtained from Outbreak.info as of 15 March 2022; BA.1 (n = 1,005,355), BA.1.1 (n = 827,907), BA.2 (n = 316,708), and BA.3 (n = 694))

**Figure 3-figure supplement 2:**
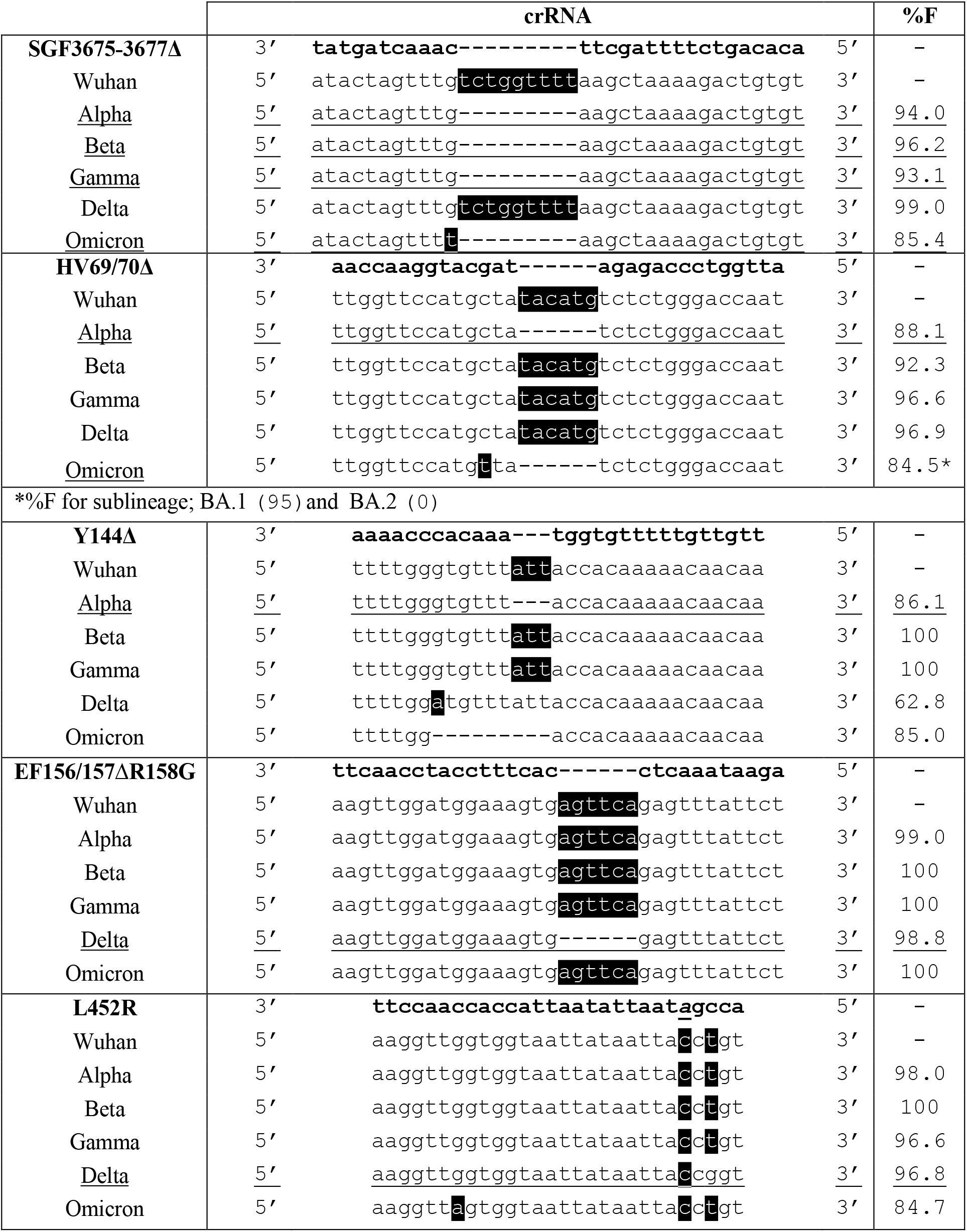
Multiple sequence alignments of the guide RNA spacer targeting a specific mutation representative of SARS-CoV-2 variant sequences. %F, frequency of the sequence among the variant genomes. The associated dataset was given in Figure 3‒source data 2. *Black shading highlights the mismatches to the corresponding crRNA. The underlines highlight the anticipated targeted variants and targeted sequences of each design*.

**Figure 3-figure supplement 3:**
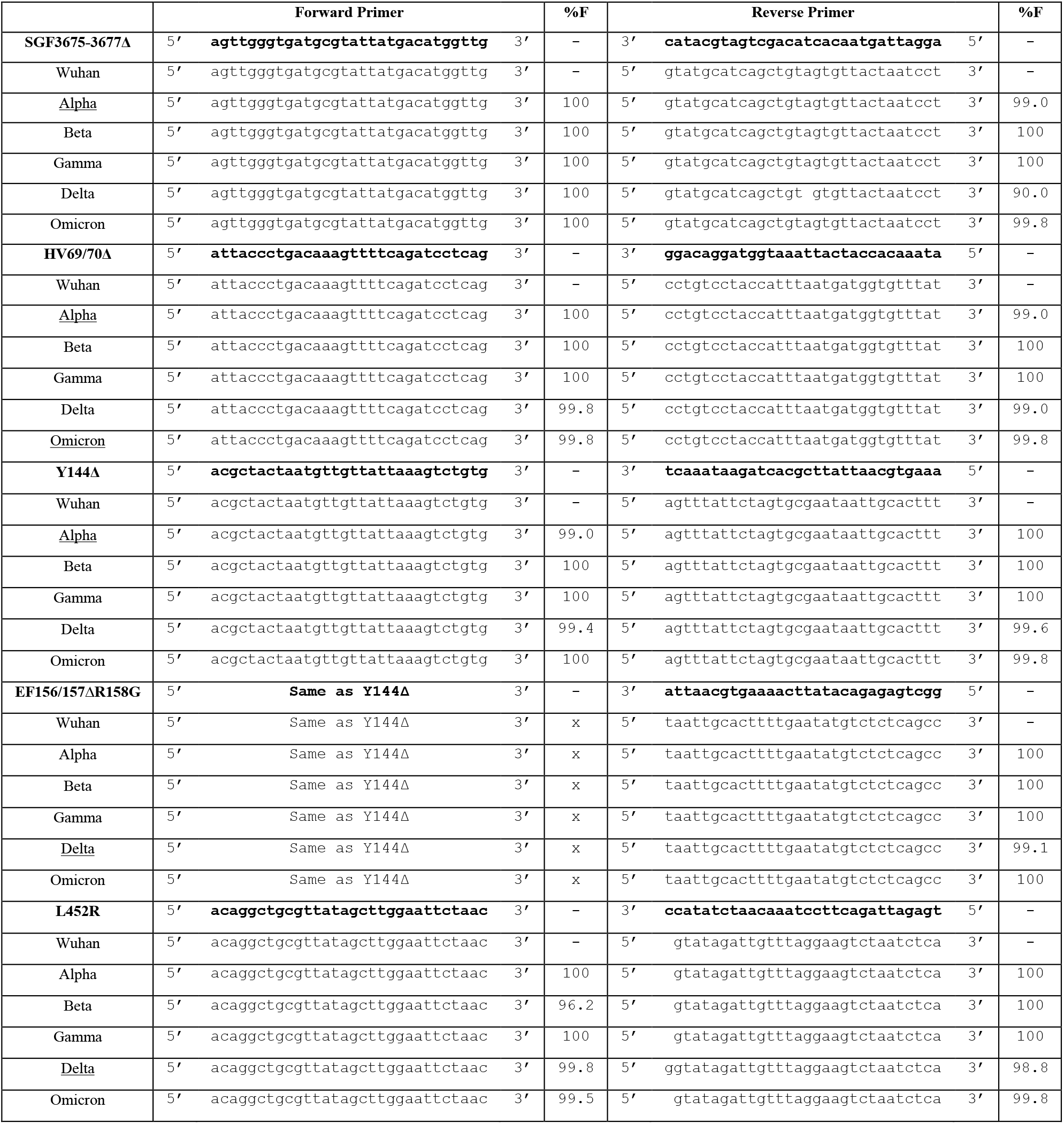
Multiple sequence alignments of primer pairs targeting a specific mutation representative of SARS-CoV-2 variant sequences. %F, frequency of the sequence among the variant isolates. The associated dataset was given in Figure 3‒source data 2. *Black shading highlights the mismatches to the corresponding crRNA. The underlines highlight the anticipated targeted variants and targeted sequences of each design*.

**Figure 3-figure supplement 4:**
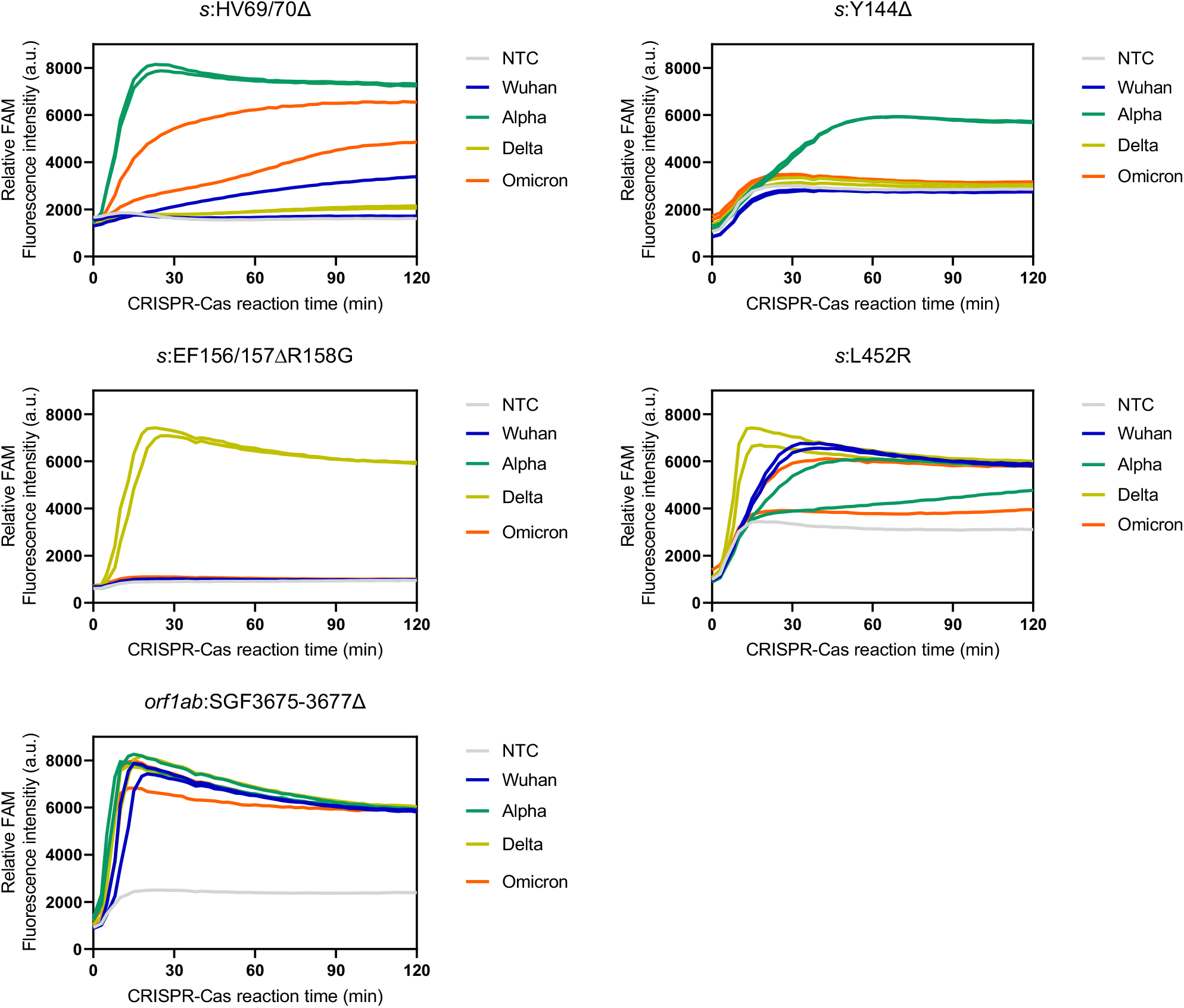
Kinetic tracing of CRISPR-Cas13a detection reaction underlying Figure 3B. Screening of primer sets and crRNA for cross-reactivity against the high viral RNA titers (C_t_ 28) of ancestral Wuhan, the alpha, and the delta strains, as well as the omicron strain. Kinetics of FAM fluorescence signal generation from the SARS-CoV-2 gene detection (as indicated) via a singleplexed CRISPR-Cas13a reaction, using cultured SARS-CoV-2 viral RNA extracts, except the omicron RNA extract from clinical sample, verified to be positive with different coronaviruses via RT-qPCR. Data are mean ± s.d. from 3 replicates. RNase-free water was used as input of all negative control reactions.

**Figure 3-figure supplement 5:**
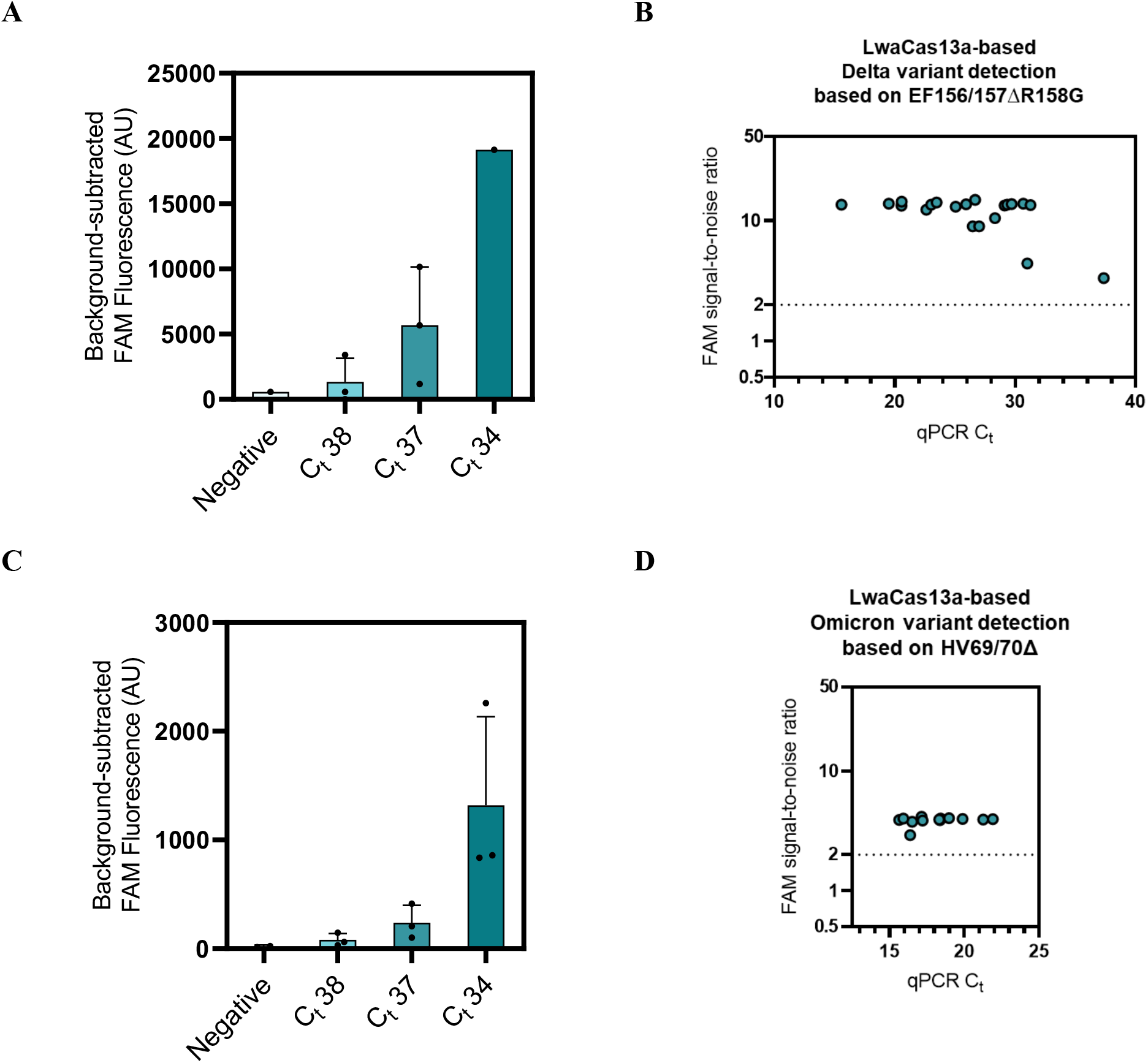
Singleplexed Detection of SARS-CoV-2 Delta and Omicron variants. **(A)** LwaCas13a-based Delta variant detection based on EF156/157ΔR158G performed on a dilution series of a SARS-CoV-2 Delta RNA extract with determined C_t_ value. FAM fluorescence at 60 minutes were subtracted with background signal generated in negative control whose input is RNase-free water. Data are mean ± s.d. from 3 replicates. **(B)** LwaCas13a-based Delta variant detection based on EF156/157ΔR158G performed on Clinical Delta samples with determined C_t_ value. FAM fluorescence at 60 minutes were normalized against intensities obtained from the no template control. Data are mean ± s.d. from 3 replicates. **(C)** LwaCas13a-based Omicron variant detection based on HV69/70Δ performed on a dilution series of a SARS-CoV-2 Omicron RNA extract with determined C_t_ value. FAM fluorescence at 60 minutes were subtracted with background signal generated in negative control whose input is RNase-free water. Data are mean ± s.d. from 3 replicates. **(D)** LwaCas13a-based Omicron variant detection based on HV69/70Δ performed on Clinical Delta samples with determined C_t_ value. FAM fluorescence at 60 minutes were normalized against intensities obtained from the no template control. Data are mean ± s.d. from 3 replicates.

**Figure 3-figure supplement 6:**
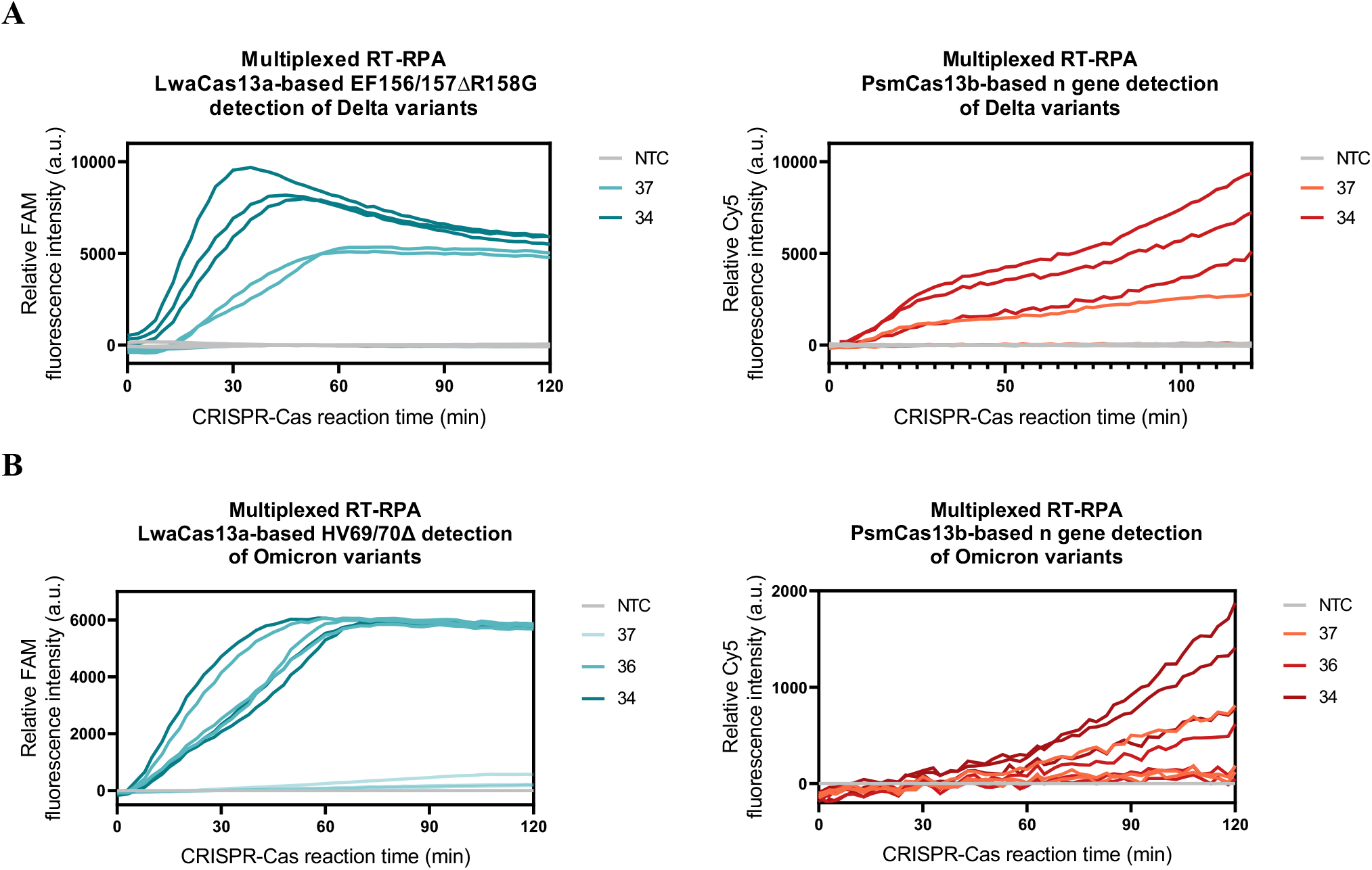
Kinetic traces of CRISPR-Cas13a/b detection reactions for Figure 3C and 3E. **(A)** Multiplexed LwaCas13a-mediated EF156/157ΔR158G (s gene) and PsmCas13b-mediated pan-SARS-CoV-2 (n gene) detection **(B)** Multiplexed LwaCas13a-mediated HV69/70Δ (s gene) and PsmCas13b-mediated pan-SARS-CoV-2 (n gene) detection. Kinetics of FAM (left) and Cy5 (right) fluorescence signal generation via a multiplexed CRISPR-Cas13a/b reaction performed on a dilution series with determined C_t_ value of Delta RNA extracts from laboratory culture **(A)** or Omicron RNA extracts from clinical samples **(B)** were shown. RNase-free water was used as input of all no template control (NTC) reactions.

**Figure 3-figure supplement 7:**
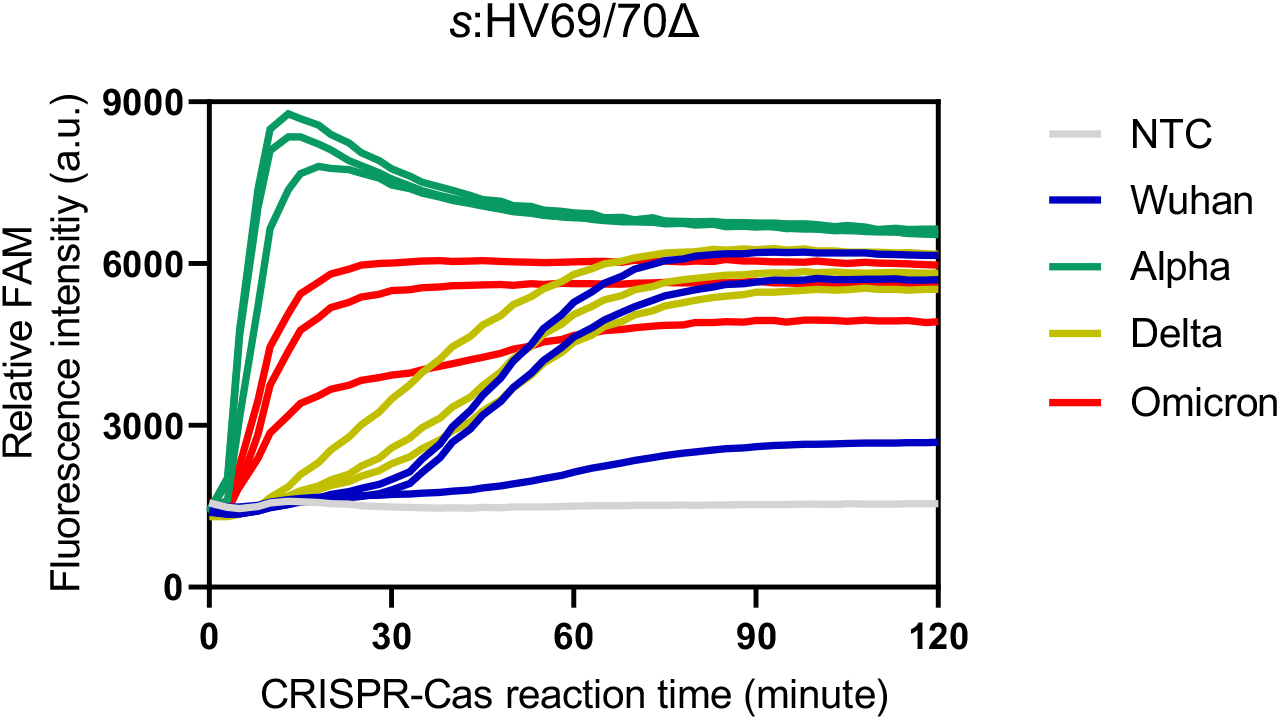
Exclusivity (cross-reactivity) testing of primer and crRNA targeting HV69/70Δ detection against the high viral RNA titers (C_t_ 25) of ancestral Wuhan, the alpha, and the delta strains, as well as the omicron strain. Kinetics of FAM fluorescence signal generation from the SARS-CoV-2 s gene harboring HV69/70Δ mutation detection via a singleplexed CRISPR-Cas13a reaction, using cultured SARS-CoV-2 viral RNA extracts, except the omicron RNA extract from clinical sample, verified to be positive with different coronaviruses via RT-qPCR. Data are mean ± s.d. from 3 replicates. RNase-free water was used as input for negative control reactions.

**Figure 3-figure supplement 8:**
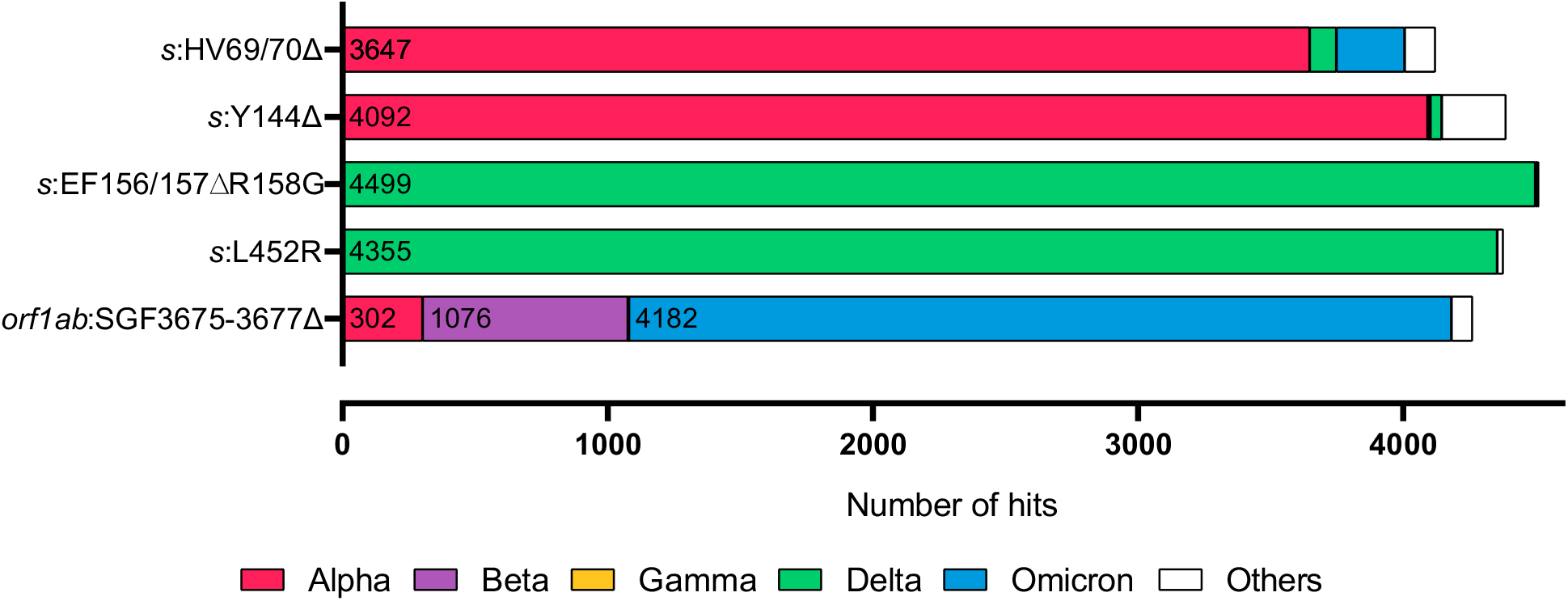
Exclusivity evaluation of designed crRNAs. Distribution of the SARS-COV-2 variant genome hits from BLAST search using crRNA spacer complementary sequences as queries. The associated dataset was given in Figure 3‒source data 1.

**Figure 4-figure supplement 1:**
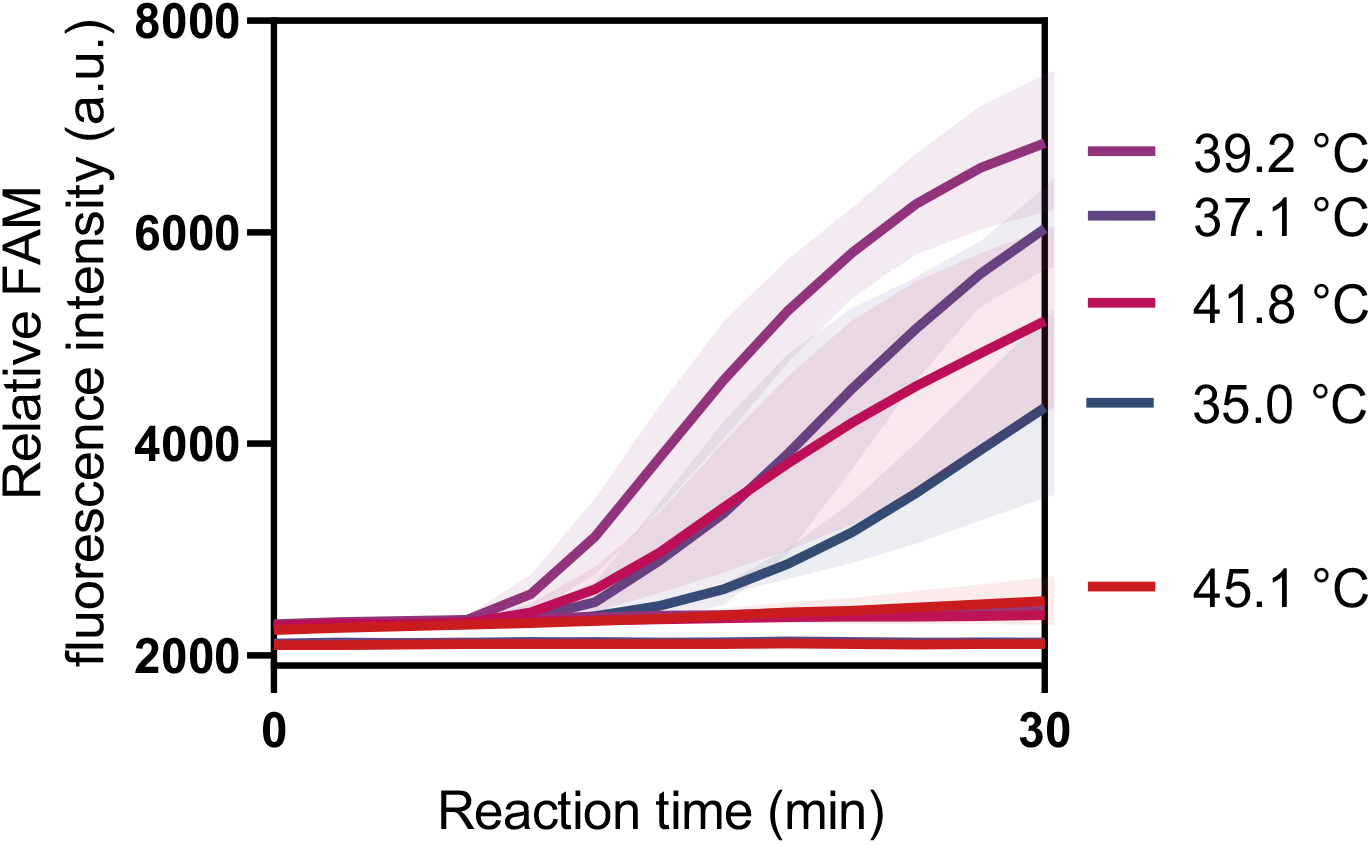
Effects of the reaction temperature on SHINE protocol. A one-pot SHINE detection was performed using *s* gene primers and LwaCas13a-crRNA targeting *s* gene at varying temperature ranging from 35.0 - 45.1 °C. FAM fluorescence was monitored using a real-time PCR (CFX Connect Real-Time PCR System, Bio-Rad). Error bars, ± s.d. from 3 replicates.

**Figure 4-figure supplement 2:**
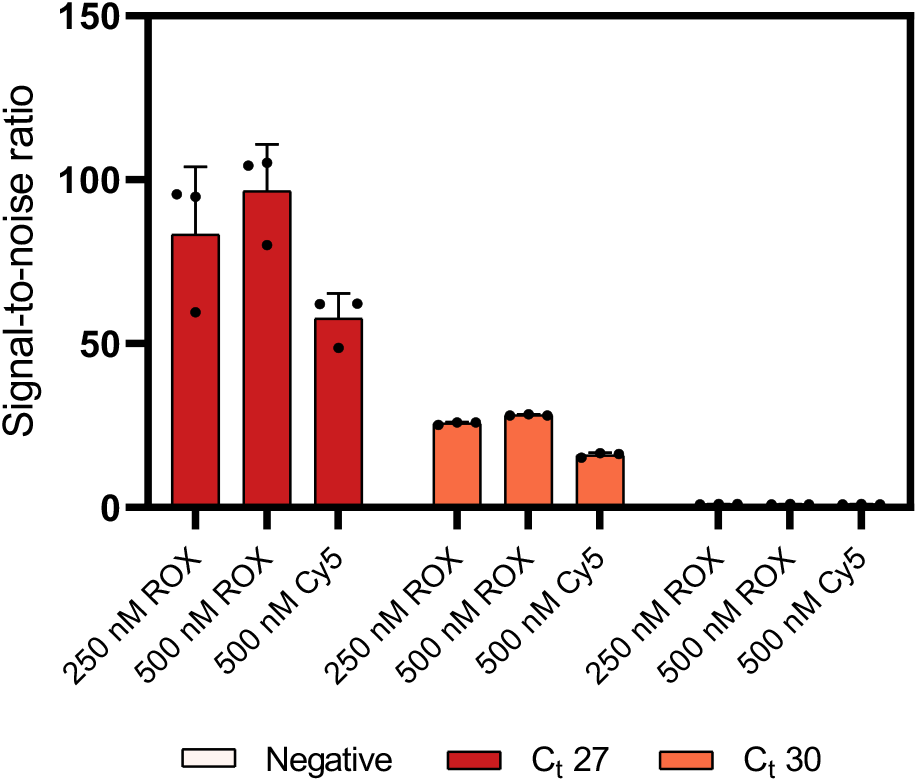
Comparing ROX-based vs Cy5-based polyA reporter for PsmCas13b-based detection. PsmCas13b-based detection of RT-RPA products with serially diluted SARS-CoV-2 RNA as input was performed with either 250 nM Rhodamine X (ROX)-PolyA reporter, 500 nM ROX- PolyA, or 500 nM Cy5-PolyA reporter. Signal-to-noise ratios (defined as fluorescence intensities of the samples divided by those from negative input samples) generated after 22.5 min of the CRISPR-Cas reaction for each condition are shown. Error bars, ± s.d. from 3 replicates.

**Figure 4-figure supplement 3:**
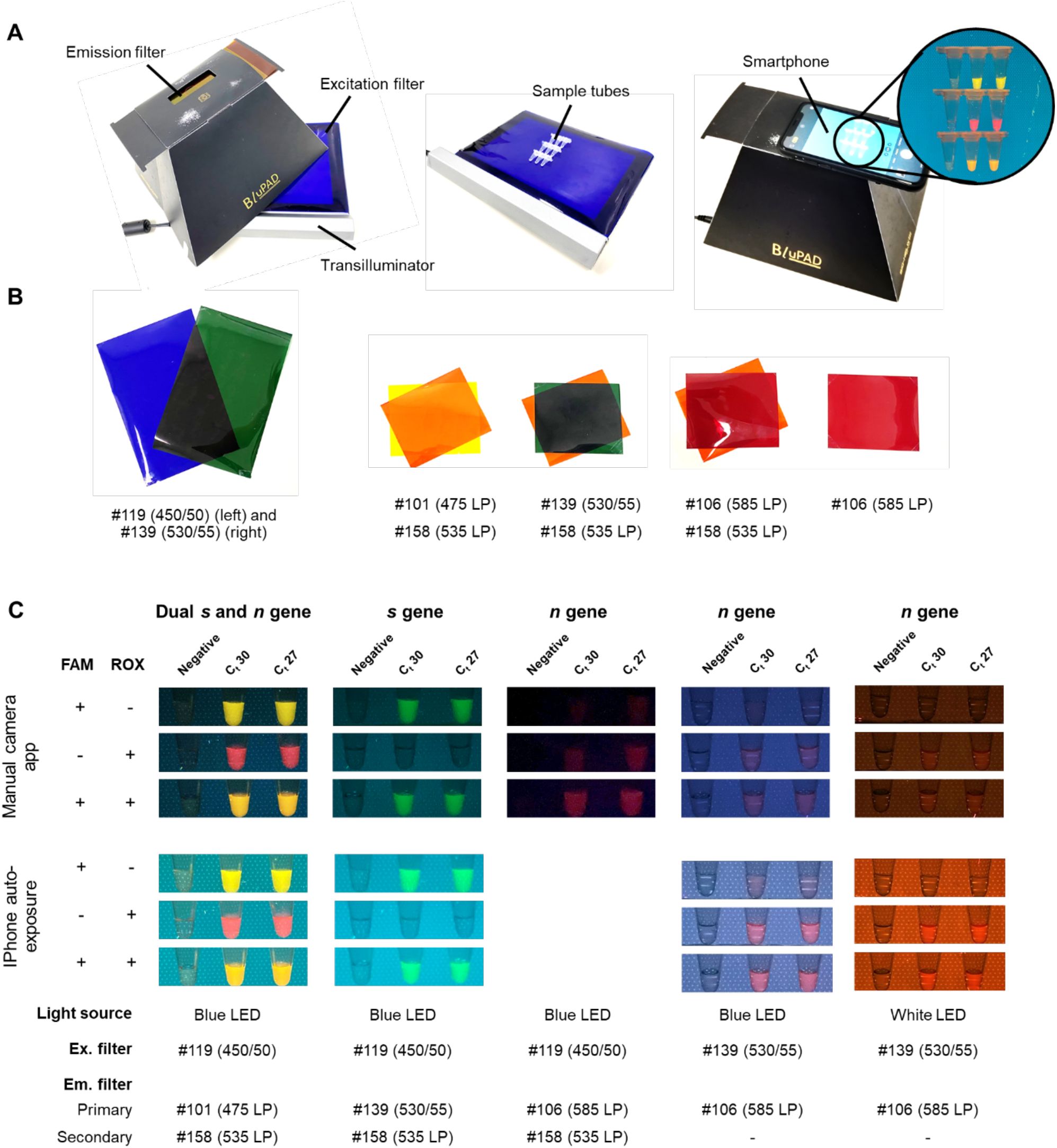
Direct-eye and smartphone-based visualization of multiplexed CRISPR-based detection. (**A**) An example of equipment setup. We used a dual blue/white LED transilluminator. (**B**) Lighting gels with appropriate light filtering for visualization of FAM and ROX. (**C**) Images taken by manual camera application (Qian et al.) and auto-exposure (bottom) using different combinations of LED light source, excitation filters, emission filters, and phone-based image acquisition for visualization of FAM and ROX signals generated from multiplexed CRISPR-based detection reactions.

**Figure 4-figure supplement 4:**
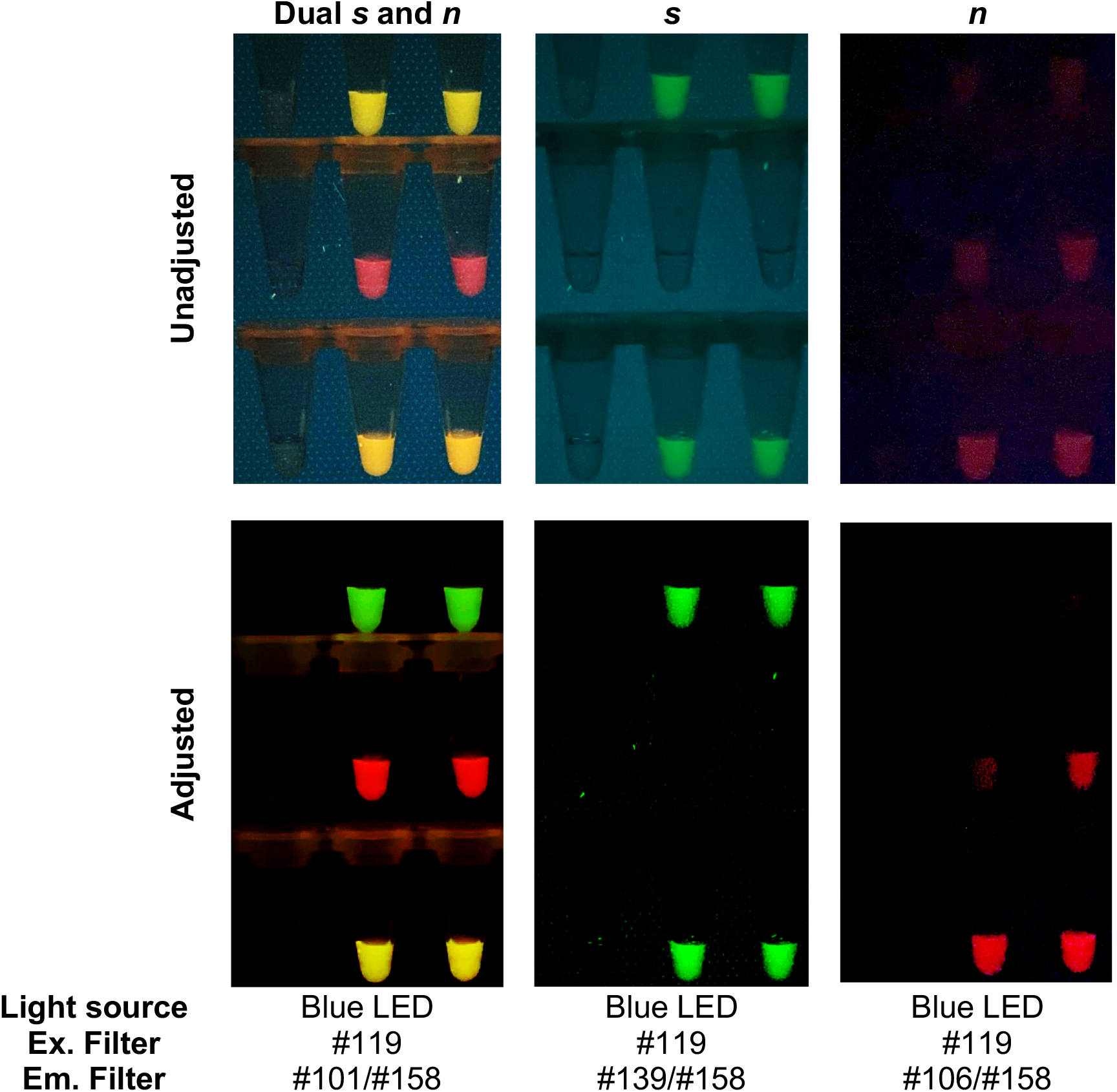
Raw smartphone images vs processed images of FAM/ROX-labeled multiplexed detection samples. The acquired images were processed using Curves adjustment menu and Channel mixers adjustment menu in Photoshop CC 2017 (Adobe Systems) to improve contrast and filter out blue color background, respectively.

**Figure 4-figure supplement 5:**
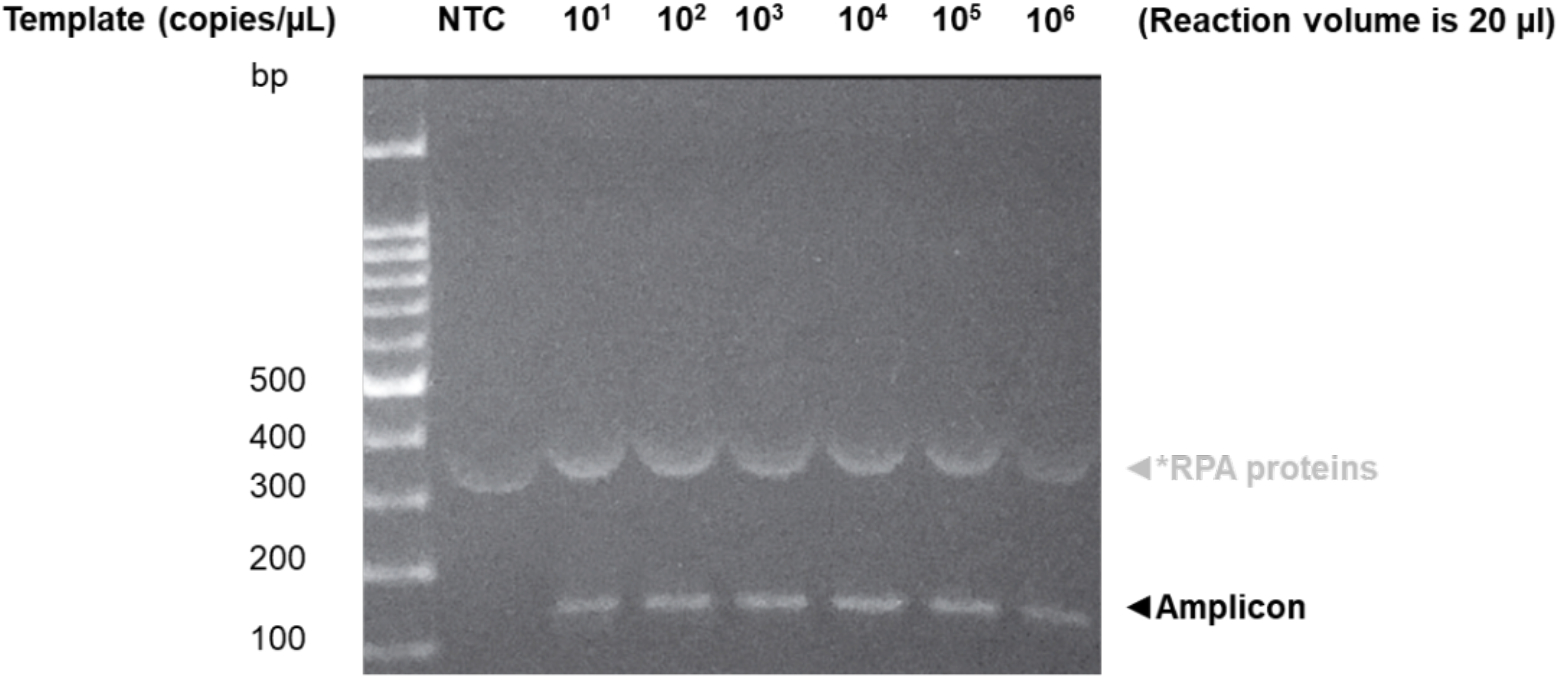
Activity of in-house RPA under standard reaction conditions. RPA amplification reactions using in-house enzyme components were performed based on the standard reaction condition as described in the methods section. The copy number of the DNA template (T7 promoter linked to the SARS-CoV-2 *s* gene) used as input was varied as indicated. NTC is a negative control with RNase-free water as input. After incubation at 37°C for 60 min, RPA products were mixed with Novel Juice DNA staining reagent (BIO-HELIX), separated on 2% (w/v) agarose gel and visualized under blue light (BluPAD, BIO-HELIX). Positions of DNA template (T7 promoter-*s* gene, 137 bp) and amplicons were indicated with black arrowheads. The Novel Juice DNA stain also weakly stains proteins present in the reaction (white arrowheads).

**Table S1.** Nomenclature of the SARS-CoV-2 lineages referenced in this study.

| **This study** | **WHO label** | **NextStrain clade** | **PANGO lineage** |
| --- | --- | --- | --- |
| Alpha | Alpha | 20I (Alpha, V1) | B.1.1.7 |
| Beta | Beta | 20H (Beta, V2) | B.1.351 |
| Gamma | Gamma | 20J (Gamma, V3) | P.1 |
| Delta | Delta | 21A (Delta)<br>21I (Delta)<br>20J (Delta) | B.1617.2 |
| Omicron | Omicron | 21K (Omicron)<br>21L (Omicron)<br>20M (Omicron) | BA.1<br>BA.1.1<br>BA.2<br>BA.3 |

**Table S2.**
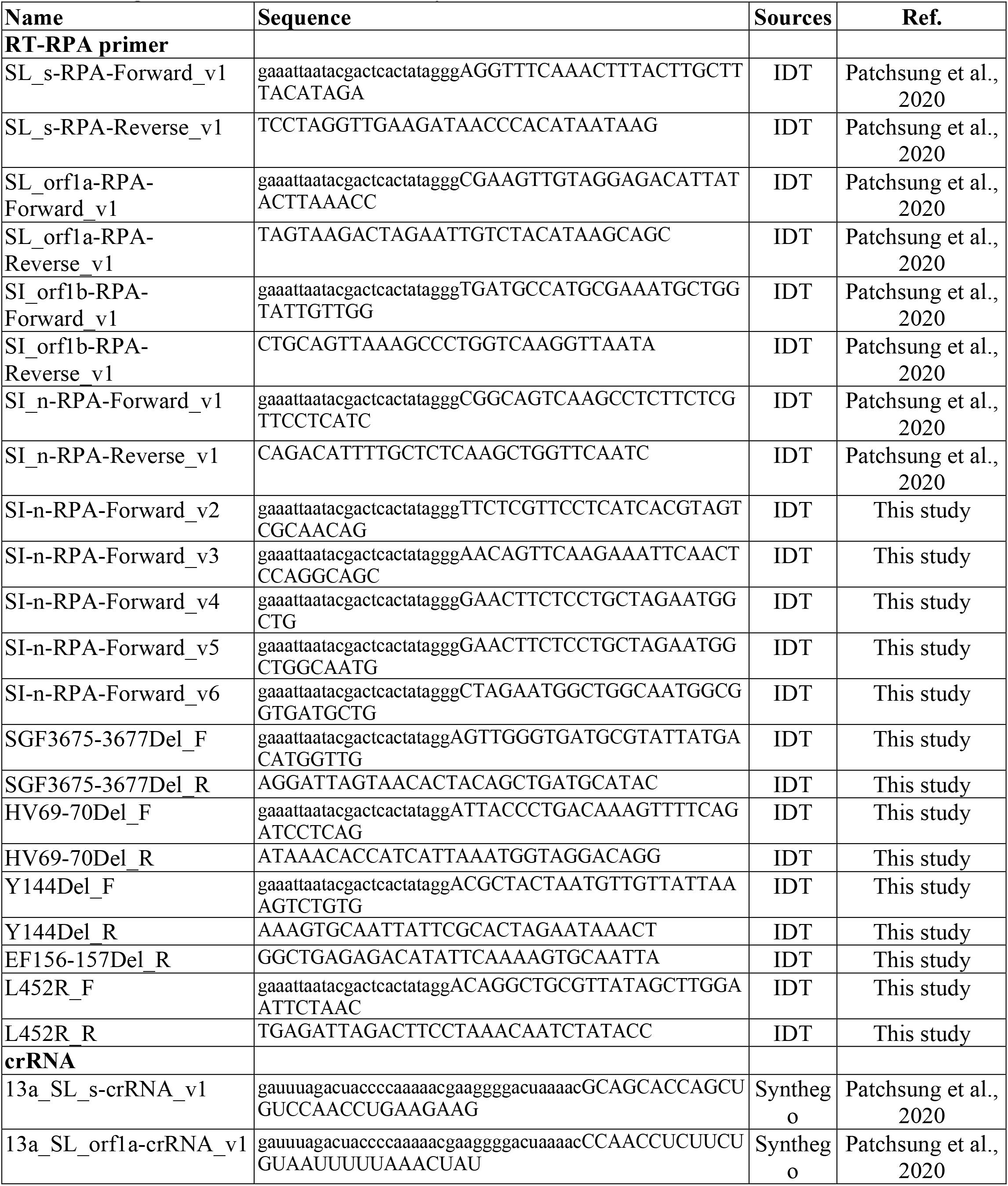
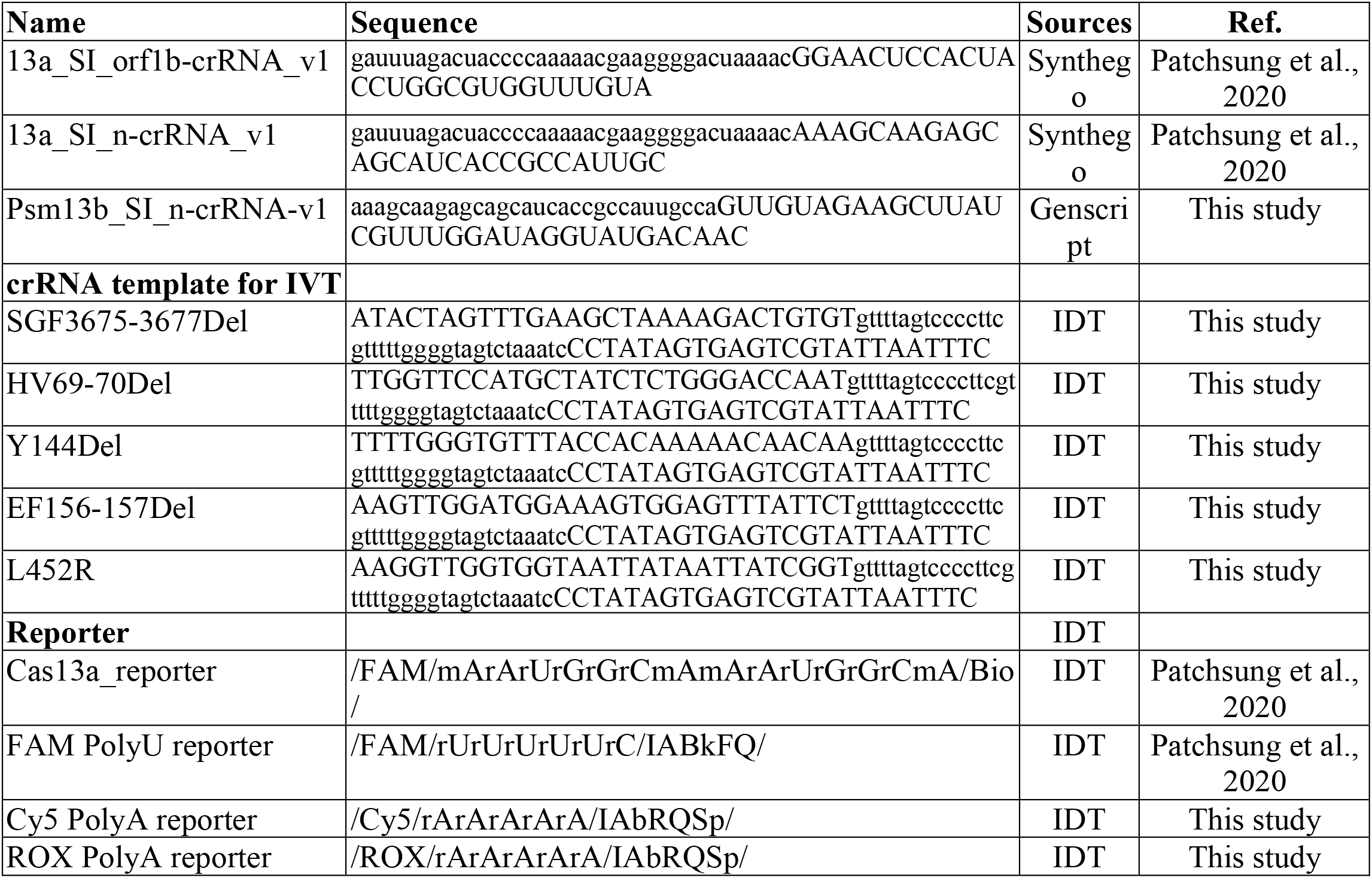
Oligonucleotides used in this study.

**Table S3.** Specific RT-RPA conditions for each figure. Aside from parameters shown in the table which were varied during optimizations, all other parameters are as given in the “Optimized multiplexed RT-RPA” methods section.

| Figure # | Primer identity and concentrations | RNase H Concentrations | Additive | RNA input |
| --- | --- | --- | --- | --- |
| Figure 2A | 177 nM each *s* primer and 222 nM *n* primer | 0.025 U/μL | 40 mM Triglycine | 5.3 |
| Figure 2B-2E | 177 nM each *s* primer and 222 nM *n* primer | 0.025 U/μL | 40 mM Triglycine | 12.4 |
| Figure 2F | 177 nM each *s* primer and 222 nM *n* primer | 0.025 U/μL | 40 mM Triglycine | 12.4 |
| Figure 3A-3B | 704 nM each primer for each variant | 0.025 U/μL | 40 mM Triglycine | 12.4 |
| Figure 3C | 177 nM each *s*(EF156/157ΔR158G) primer and 177 nM *n* primer | 0.025 U/μL | 40 mM Triglycine | 12.4 |
| Figure 3D | 177 nM each *s*(EF156/157ΔR158G) primer and 177 nM *n* primer | 0.025 U/μL | 40 mM Triglycine | 12.4 |
| Figure 3E | 177 nM each HV69/70Δ primer and 177 nM *n* primer | 0.025 U/μL | 40 mM Triglycine | 12.4 |
| Figure 3F | 177 nM each HV69/70Δ primer and 177 nM *n* primer | 0.025 U/μL | 40 mM Triglycine | 12.4 |
| Figure 4A | 177 nM each *s* primer and 222 nM *n* primer | 0.025 U/μL | 40 mM Triglycine | 5.3 |
| Figure 1–figure supplement 1C | 709 nM each *n* primer | - | - | 6.3 μL |
| Figure 1–figure supplement 1D | 355 nM each primer for each gene | - | - | 6.3 μL |
| Figure 1–figure supplement 2A, 2B | varying concentration of *s* and *N* primer pair | 0.025 U/μL | - | 6.3 μL |
| Figure 1–figure supplement 2C | 709 nM each *s* primer | Varying RNase H concentrations | - | 6.3 μL |
| Figure 1–figure supplement 2D | 704 nM each *n* primer, 704 each *s* primer, and 177 nM each *s* primer and 222 nM *n* primer for *s*, *n* and *s+n* respectively | 0.025 U/μL | 40 mM Triglycine | 5.3 μL |
| Figure 1–figure supplement 2E | 704 nM each *n* primer | 0.025 U/μL | 40 mM Triglycine | 5.3 μL |
| Figure 1–figure supplement 4 | 709 nM each *n* primer | - | - | 6.3 μL |
| Figure 1–figure supplement 5 | 709 nM each *n* primer | 1 U/μL | - | 6.3 μL |
| Figure 1–figure supplement 6 | 709 nM each *n* primer | - | - | 6.3 μL |
| Figure 1–figure supplement 7A | 177 nM each *s* and *n* primer | 0.025 U/μL | Varying additives | 3.2 μL |
| Figure 1–figure supplement 7B | 177 nM each *s* and *n* primer | 0.025 U/μL | - | 6.3 μL |
| Figure 1–figure supplement 7C | 177 nM each *s* and *n* primer | 0.025 U/μL | With and without 40 mM Triglycine | 3.2 μL |
| Figure 1–figure supplement 8 | 177 nM each *s* primer and 222 nM *n* primer | 0.025 U/μL | 40 mM Triglycine | 5.3 μL |
| Figure 1–figure supplement 10 | 704 nM each *n* primer | 0.025 U/μL | 40 mM Triglycine | 5.3 μL |
| Figure 1–figure supplement 11 | 177 nM each *s* primer and 222 nM *n* primer | 0.025 U/μL | 40 mM Triglycine | 5.3 μL |
| Figure 2–figure supplement 1 | 177 nM each *s* primer and 222 nM *n* primer | 0.025 U/μL | 40 mM Triglycine | 12.4 μL |
| Figure 3–figure supplement 4 | 704 nM each primer for each variant | 0.025 U/μL | 40 mM Triglycine | 12.4 μL |
| Figure 3–figure supplement 5 | 704 nM each *s*(EF156/157ΔR158 G) primer | 0.025 U/μL | 40 mM Triglycine | 12.4 μL |
| Figure 3–figure supplement 6 | 177 nM each *s*(EF156/157ΔR158 G or HV69/70Δ) primer and 177 nM *n* primer | 0.025 U/μL | 40 mM Triglycine | 12.4 μL |
| Figure 3–figure supplement 7 | 704 nM each *s*(HV69/70Δ) primer | 0.025 U/μL | 40 mM Triglycine | 12.4 μL |

**Table S4.** Specific Cas13-based reaction conditions for each figure. Aside from parameters shown in the table which were varied during optimizations, all other parameters are as given in the “Optimized multiplexed Cas13-based detection with fluorescence readout” methods section.

| Figure | Buffer identity and concentration | Cas enzyme identity and concentration | crRNA identity and concentration | RNA reporter identity and concentration | additive | MgCl<sub>2</sub> concentration | Equipment used to monitor Cas-based reactions |
| --- | --- | --- | --- | --- | --- | --- | --- |
| Figure 2<br>Multiplexed LwaCas13a-PsmCas13b detection | 20 mM HEPES pH 6.8 | 45 nM LwaCas13a, 135 nM PsmCas13b, | 22.5 nM LwaCas13a crRNA targeting *s* gene, 67.5 nM PsmCas13b crRNA targeting *n* gene | 250 nM PolyU-FAM, 500 nM PolyA-ROX | 750 mM betaine | 24 mM | Tecan microplate reader |
| Figure 2<br>LwaCas13a Dual S/N detection | 20 mM HEPES pH 6.8 | 45 nM LwaCas13a | 22.5 nM LwaCas13a crRNA targeting *s* gene, 22.5 nM LwaCas13a crRNA targeting *n* gene | 250 nM PolyU-FAM | 750 mM betaine | 24 mM | Tecan microplate reader |
| Figure 3A-3B | 40 mM Tris pH 7.4 | 45 nM LwaCas13a | 22.5 nM LwaCas13a crRNA targeting each variant | 250 nM PolyU-FAM | - | 6 mM | Tecan microplate reader |
| Figure 3C | 40 mM Tris pH 7.4 | 45 nM LwaCas13a, 135 nM PsmCas13b, | 22.5 nM LwaCas13a crRNA targeting each variant, 67.5 nM PsmCas13b crRNA targeting *n* gene | 250 nM PolyU-FAM, 500 nM PolyA-ROX | 750 mM betaine | 24 mM | Tecan microplate reader |
| Figure 3D | 40 mM Tris pH 7.4 | 45 nM LwaCas13a, | 22.5 nM LwaCas13a crRNA targeting | 250 nM PolyU-FAM, | 750 mM betaine | 24 mM | Tecan microplate reader |
|  |  | 135 nM PsmCas13b | each variant, 67.5 nM PsmCas13b crRNA targeting *n* gene | 500 nM PolyA-ROX |  |  |  |
| Figure 3E | 40 mM Tris pH 6.8 | 45 nM LwaCas13a, 135 nM PsmCas13b | 22.5 nM LwaCas13a crRNA targeting HV69/70Δ, 67.5 nM PsmCas13b crRNA targeting *n* gene | 250 nM PolyU-FAM, 500 nM PolyA-ROX | - | 24 mM | Tecan microplate reader |
| Figure 3F | 40 mM Tris pH 6.8 | 45 nM LwaCas13a, 135 nM PsmCas13b | 22.5 nM LwaCas13a crRNA targeting HV69/70Δ, 67.5 nM PsmCas13b crRNA targeting *n* gene | 250 nM PolyU-FAM, 500 nM PolyA-ROX | - | 24 mM | Tecan microplate reader |
| Figure 4A | 40 mM Tris pH 7.4 | 45 nM LwaCas13a | 22.5 nM LwaCas13a crRNA targeting *s* gene, 22.5 nM LwaCas13a crRNA targeting *n* gene | 250 nM PolyU-FAM | - | 6 mM | Real-time PCR |
| Figure 4B | 20 mM HEPES pH 6.8 | 45 nM LwaCas13a, 135 nM PsmCas13b, | 22.5 nM LwaCas13a crRNA targeting *s* gene, 67.5 nM PsmCas13b crRNA targeting *n* gene | Only 250 nM PolyU-FAM, only 500 nM PolyA-ROX, or both | 750 mM betaine | 24 mM | Naked eye visualization |
| Figure 1–<br>figure supplement 1C, 1D | 40 mM Tris pH 7.4 | 45 nM LwaCas13a | 22.5 nM LwaCas13a crRNA for each gene | 1 μM FAM-Bio | - | 6 mM | Lateral flow strip |
| Figure 1–<br>figure supplement 2A, 2B, 2D | 20 mM HEPES pH 6.8 | 45 nM LwaCas13a, 135 nM PsmCas13b, | 22.5 nM LwaCas13a crRNA targeting *s* gene, 67.5 nM PsmCas13b crRNA targeting *n* gene | 250 nM PolyU-FAM, 500 nM PolyA-Cy5 | - | 24 mM | Tecan microplate reader |
| Figure 1–<br>figure supplement 2C | 40 mM Tris pH 7.4 | 45 nM LwaCas13a | 22.5 nM LwaCas13a crRNA for *s* gene | 250 nM PolyU-FAM | - | 6 mM | Tecan microplate reader |
| Figure 4B | 20 mM HEPES pH 6.8 | 45 nM LwaCas13a, 135 nM PsmCas13b, | 22.5 nM LwaCas13a crRNA targeting *s* gene, 67.5 nM PsmCas13b crRNA targeting *n* gene | Only 250 nM PolyU-FAM, only 500 nM PolyA-ROX, or both | 750 mM betaine | 24 mM | Naked eye visualization |
| Figure 4D, 4E, and 4F | 40 mM Tris pH 7.4 | 45 nM LwaCas13a | 22.5 nM LwaCas13a crRNA for *n* gene | 250 nM PolyU-FAM | - | 6 mM | Tecan microplate reader |
| Figure 4G | 40 mM Tris pH 7.4 | 45 nM LwaCas13a | 22.5 nM LwaCas13a crRNA targeting *S* gene, 22.5 nM LwaCas13a crRNA targeting *n* gene | 250 nM PolyU-FAM | - | 6 mM | Real-time PCR |
| Figure 4H | 40 mM Tris pH 7.4 | 45 nM LwaCas13a | 22.5 nM LwaCas13a crRNA targeting *S* gene, 22.5 nM LwaCas13a<br>crRNA targeting *n* gene | 250 nM PolyU-FAM | - | 6 mM | Real-time PCR |
| Figure 1–<br>figure supplement 1 | 40 mM Tris pH 7.4 | 45 nM LwaCas13a | 22.5 nM LwaCas13a crRNA for each gene | 1 µM FAM-Bio | - | 6 mM | Lateral flow strip |
| Figure 1–<br>figure supplement 1D | 40 mM Tris pH 7.4 | 45 nM LwaCas13a | 22.5 nM LwaCas13a crRNA for each gene | 1 µM FAM-Bio | - | 6 mM | Lateral flow strip |
| Figure 1–<br>figure supplement 2A, 2B, 2D | 20 mM HEPES pH 6.8 | 45 nM LwaCas13a, 135 nM PsmCas13b, | 22.5 nM LwaCas13a crRNA targeting *s* gene, 67.5 nM PsmCas13b crRNA targeting *n* gene | 250 nM PolyU-FAM, 500 nM PolyA-Cy5 | - | 24 mM | Tecan microplate reader |
| Figure 1–<br>figure supplement 2C | 40 mM Tris pH 7.4 | 45 nM LwaCas13a | 22.5 nM LwaCas13a crRNA for *s* gene | 250 nM PolyU-FAM | - | 6 mM | Tacan microplate reader |
| Figure 1–<br>figure supplement 2D | 40 mM Tris pH 7.4 | 45 nM LwaCas13a | 22.5 nM LwaCas13a crRNA for *s* gene or *n* gene | 250 nM PolyU-FAM | - | 6 mM | Tacan microplate reader |
| Figure 1–<br>figure supplement 2E | 40 mM Tris pH 7.4 | 45 nM LwaCas13a | 22.5 nM LwaCas13a crRNA for *n* gene | 250 nM PolyU-FAM | - | 6 mM | Tacan microplate reader |
| Figure 1–<br>figure supplement 4A | 20 mM HEPES pH 6.8 | Varying PsmCas13b amount | Varying PsmCas13b crRNA | 250 nM PolyA-Cy5 | - | 6 mM | Varioskan microplate reader |
| Figure 1–<br>figure supplement 4B | 20 mM HEPES pH 6.8 | 135 nM PsmCas13b | 67.5 nM PsmCas13b crRNA | varying PolyA-Cy5 amount | - | 6 mM | Varioskan microplate reader |
| Figure 1–<br>figure supplement 5<br>LwaCas13a | 20 mM HEPES pH 6.8 | 45 nM LwaCas13a | 22.5 nM LwaCas13a crRNA for *n* gene | 250 nM PolyU-FAM | - | 6 mM | Varioskan microplate reader |
| Figure 1–<br>figure supplement 5<br>PsmCas13b | 20 mM HEPES pH 6.8 | 135 nM PsmCas13b | 67.5 nM PsmCas13b crRNA | 500 nM PolyA-Cy5 | - | 6 mM | Varioskan microplate reader |
| Figure 1–<br>figure supplement 6B | 40 mM Tris pH 7.4 | 45 nM RfxCas13d | 28.4 nM RfxCas13d crRNA | 250 nM PolyU-FAM reporter or 125 nM RNase Alert | - | 6 mM | Varioskan microplate reader |
| Figure 1–<br>figure supplement 6C | 40 mM Tris pH 7.4 | 45 nM RfxCas13d or RfxCas13d-RBD or LwaCas13a | 22.5 nM LwaCas13a crRNA or 28.4 nM RfxCas13d crRNA | 500 nM PolyU/FAM reporter | - | 6 mM | Tecan microplate reader |
| Figure 1–<br>figure supplement 7A | 40 mM Tris pH 7.4 | 45 nM LwaCas13a | 22.5 nM LwaCas13a crRNA for target gene | 250 μM PolyU-FAM | - | 6 mM | Tecan microplate reader |
| Figure 1–<br>figure supplement 7B | 20 mM HEPES pH 6.8 | 45 nM LwaCas13a, 135 nM PsmCas13b, | 22.5 nM LwaCas13a crRNA targeting *s* gene, 67.5 nM PsmCas13b crRNA targeting *n* gene | 250 nM PolyU-FAM, 500 nM PolyA-Cy5 | Varying additives | 24 mM | Tecan microplate reader |
| Figure 1–<br>figure supplement 7C | 20 mM HEPES pH 6.8 | 45 nM LwaCas13a, 135 nM PsmCas13b, | 22.5 nM LwaCas13a crRNA targeting *s* gene, 67.5 nM PsmCas13b crRNA targeting *n* gene | 250 nM PolyU-FAM, 500 nM PolyA-Cy5 | With and without 500 mM betaine | 24 mM | Tecan microplate reader |
| Figure 1–<br>figure supplement 8 | 20 mM HEPES pH 6.8 | 45 nM LwaCas13a, 135 nM PsmCas13b, | 22.5 nM LwaCas13a crRNA targeting *s* gene, 67.5 nM PsmCas13b crRNA targeting *n* gene | 250 nM PolyU-FAM, 500 nM PolyA-Cy5 | Varying betaine amount | 24 mM | Tecan microplate reader |
| Figure 1–<br>figure supplement 9A | 40 mM Tris pH 7.4 | Varying amount of LwaCas13a | Varying amount of LwaCas13a crRNA targeting *s* gene | 250 nM PolyU-FAM | - | 6 mM | RT-PCR machine |
| Figure 1–<br>figure supplement 9B | 20 mM HEPES pH 6.8 | Varying amount of PsmCas13b | Varying amount of PsmCas13b crRNA targeting *n* gene | 500 nM PolyA-Cy5 | - | 6 mM | Varioskan microplate reader |
| Figure 1–<br>figure supplement 9C | Varying type of buffers | 45 nM LwaCas13a, 45 nM PsmCas13b | 45 nM LwaCas13a crRNA targeting *s* gene, 45 nM PsmCas13b crRNA targeting *n* gene | 250 nM PolyU-FAM, 500 nM PolyA-Cy5 | - | 6 mM | Tecan microplate reader |
| Figure 1–<br>figure supplement 10 | 40 mM Tris pH 7.4 | 45 nM LwaCas13a | 22.5 nM LwaCas13a crRNA targeting *n* gene |  | - | 6 mM | Real-time PCR |
| Figure 1–<br>figure supplement 11 | 40 mM Tris pH 7.4 | 45 nM LwaCas13a | 22.5 nM LwaCas13a crRNA targeting *S* gene, 22.5 nM LwaCas13a crRNA targeting *n* gene (s/n gene detection) | 250 nM PolyU-FAM | 750 mM betaine | 24 mM | Tecan microplate reader |
| Figure 4–<br>figure supplement 2 | 20 mM HEPES pH 6.8 | 45 nM LwaCas13a, 135 nM PsmCas13b, | 22.5 nM LwaCas13a crRNA targeting *s* gene, 67.5 nM PsmCas13b crRNA targeting *n* gene | 250 nM PolyU-FAM with 500 nM PolyA-Cy5, 250 nM or 500 nM PolyA-ROX | 750 mM betaine | 24 mM | Tecan microplate reader |

**Table S5.** The limit of detection (LoD) of the multiplexed CRISPR-based detection of SARS-CoV-2 RNA. Estimated LoDs at 95% positive rate (95% LoD) from LwaCas13a-mediated *s* gene detection, PsmCas13b-mediated *n* gene detection, LwaCas13a-mediated *s* and *n* gene detection, or combined LwaCas13a-mediated *s* gene and PsmCas13b-mediated *n* gene detection determined using Probit regression. Calculation was performed using MedCalc (MedCalc software Ltd).

| **95% LoD qPCR C<sub>t</sub> (95% CI)** | **At 60 minutes**<br>**CRISPR reaction time** |
| --- | --- |
| **Detecting the *s* gene via LwaCas13a** | 36.83* |
| **Detecting the *n* gene via PsmCas13b** | 36.78* |
| **Detecting either the *s* or *n* gene via LwaCas13a** | 36.48 (35.99 – 36.97) |
| **Detecting either the *s* or *n* gene via**<br>**LwaCas13a/PsmCas13b** | 37.01* |

\*The margins of error were negligible.

## Appendix 1

### The linear correlation between qPCR Ct value and RNA copy number obtained from ddPCR

RT-qPCR or ddPCR analyses were performed on paired samples from a dilution series of RNA extract from the cultured SARS-CoV-2 Wuhan strain. The experiment was carried out in triplicate (Appendix Table 1). Associated instructions were described in the *Materials and Methods* section. Linear regression analysis was performed. (Appendix Figure 1)

**Appendix Figure 1.**
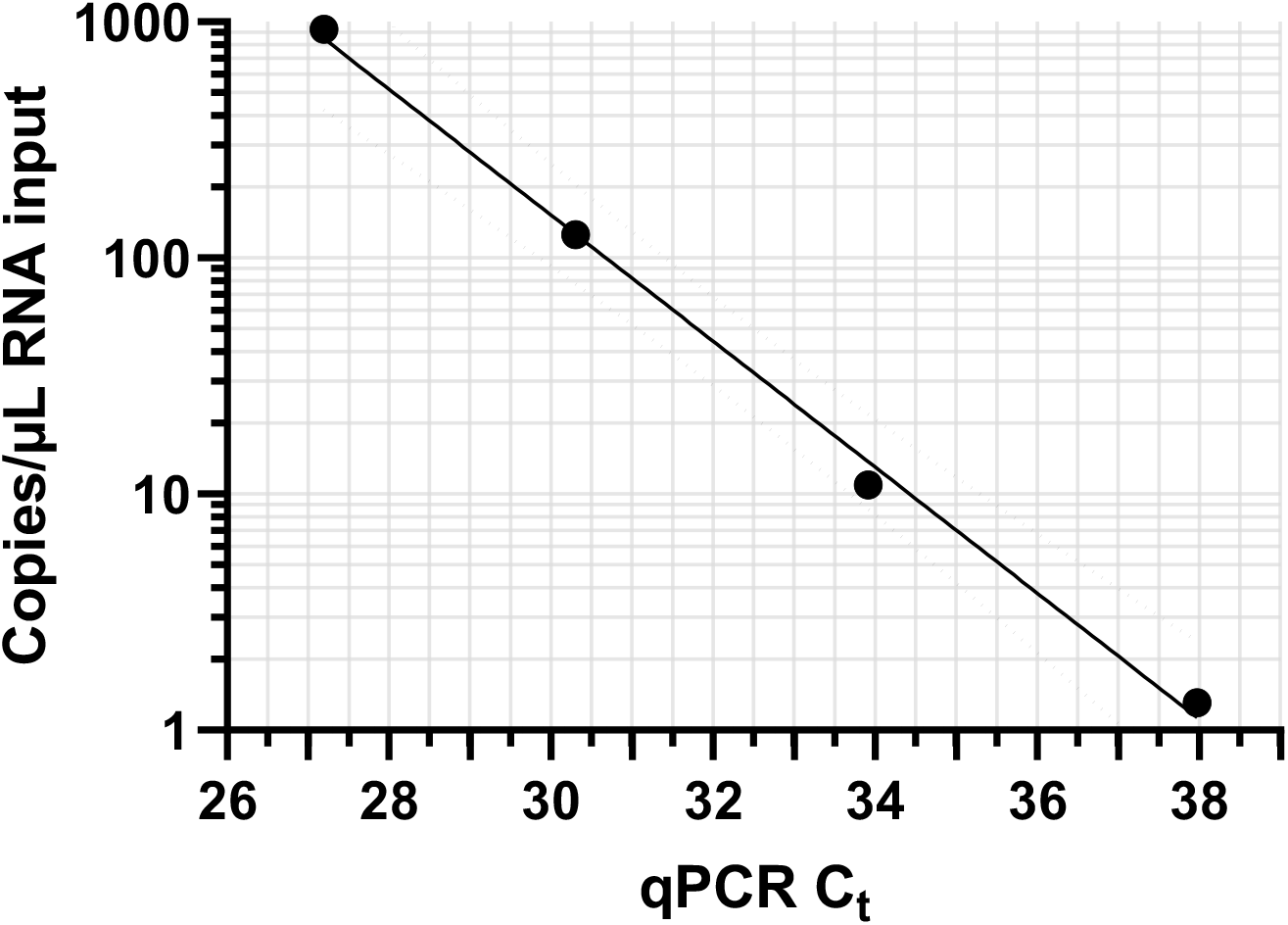
A standard plot of qPCR Ct value versus copies per μL obtained from ddPCR. Data are mean from three replicates. Linear regression equation: log_10_ (Copies per μL input) = -0.2669 (qPCR C_t_) + 10.187, R^2^ = 0.9967*. Dashed lines represent the 95% confidence intervals*

**Appendix Table 1.** Raw data underlying Appendix Figure 1

| Dilution | qPCR Ct | | | | | Copies per $\mu\text{L}$ input | | | | |
| --- | --- | --- | --- | --- | --- | --- | --- | --- | --- | --- |
|  | Triplicates |  |  | Mean | SD | Triplicates |  |  | Mean | SD |
| 1x | 27.15 | 27.20 | 27.23 | 27.19 | 0.04 | 912 | 896 | 984 | 930.67 | 46.88 |
| 10x | 30.31 | 30.30 | 30.30 | 30.30 | 0.01 | 114.8 | 137.2 | 123.6 | 125.20 | 11.29 |
| 100x | 34.12 | 33.75 | 33.86 | 33.91 | 0.19 | 8.4 | 12.4 | 12 | 10.93 | 2.20 |
| 1000x | 38.30 | 38.25 | 37.36 | 37.97 | 0.53 | 1.12 | 0.4 | 2.4 | 1.31 | 1.01 |

